# Deep Agentic Variant Prioritisation for Expert Level Genetic Diagnosis

**DOI:** 10.64898/2026.02.17.26346421

**Authors:** Muti Kara, Ahmet Furkan Gungor, Safa Eren Kuday, Oguzhan Ozcelik, Serdar Ceylaner, Fatma Dereli, Furkan Ozden

**Affiliations:** Lidya Genomics, Turkey; Department of Computer Science, Bilkent University, Turkey; Department of Computer Science, ETH Zurich, Switzerland; Intergen Genetic and Rare Diseases Diagnosis and Research Center, Turkey; Department of Computer Science, University of Oxford, United Kingdom

**Keywords:** In-context learning, Variant prioritization, Rare disease, Genetic diagnosis

## Abstract

Diagnosing rare genetic disease hinges on identifying the causal variant among thousands of candidates, a task that leaves most of the 300 million affected patients undiagnosed despite the availability of genome sequencing. Here we present DAVP (Deep Agentic Variant Prioritisation), a hierarchical agentic-LLM pipeline combining gene pre-filtering, knowledge-graph-grounded variant reports, and iterative LLM tournaments to rank variants against each patient’s phenotype. We benchmark DAVP on 709 diagnosis cases across four datasets, including LIRDB-47—a new cohort of 47 Turkish rare-disease patients with 63 clinically diagnosed causal variants. DAVP achieves 85.5% variant-level top-3 recall on the full benchmark; on LIRDB-47, it places the causal variant in the top-3 in 76.2% of patients (top-20 in 88.9%), a 1.6-fold improvement over the best phenotype-driven baseline (Exomiser, 47.6%). Our work suggests that agentic LLM systems combining evidence synthesis with patient-specific reasoning have the potential to reshape clinical genomic workflows.

## Introduction

Rare diseases affect approximately 300 million people worldwide in more than 7,000 different disorders, yet patients endure an average diagnostic odyssey of 4.7 years, with 73% experiencing at least one misdiagnosis [1, 2]. This delay carries devastating consequences: 30% of children with rare diseases die before age 5, while delayed diagnosis incurs avoidable costs ranging from $86,000 to $517,000 per patient [3, 4]. As approximately 80% of rare diseases have genetic origins [5], genomic sequencing has emerged as a critical diagnostic tool, yet the massive data volume has created analytical bottlenecks exceeding human cognitive capacity. Whole exome sequencing identifies 20,000 coding variants per individual, with 97.8-99.6% classified as rare variants requiring evaluation [6–8]. Although computational filtering reduces this to hundreds of candidates, configuration requires substantial bioinformatics expertise across thousands of parameter combinations, creating an error-prone process susceptible to missing critical variants [9]. Variant interpretation remains the critical bottleneck: 41% of patients receive variants of uncertain significance (VUS), with only 7.3% eventually reclassified after > 30 months [10, 11]. Pooled diagnostic yields remain 38.6% for whole genome and 37.8% for whole exome sequencing, leaving most rare disease patients undiagnosed despite comprehensive testing [12, 13]. While ultra-rapid sequencing achieves 36-hour technical turnaround [14], the diagnostic odyssey averages 4.7 years [2], compounded by 61% of new disease-gene associations receiving only “Limited” validity classification [15, 16]. Traditional ACMG/AMP guidelines showed only 34% initial inter-laboratory concordance [17, 18]. Computational predictors (SIFT, PolyPhen-2, CADD, REVEL) misclassify up to one-third of benign variants as pathogenic and operate without patient context [19–22]. Platforms like Exomiser integrate phenotype data but fail with novel genes or atypical presentations [23, 24]. Recent AI advances show promise, GPT-4 achieved 90.5% on USMLE Step 3 [25], PhenoBrain outperformed specialists [26], AI-MARRVEL doubled diagnostic rates [27], and AlphaMissense predicted pathogenicity for 71 million missense variants [28], yet these systems lack multi-step reasoning and dynamic evidence gathering [29]. Agentic AI systems offer a promising direction through autonomous tool use, self-correction, and goal-directed behavior [30], with recent medical applications demonstrating enhanced diagnostic capabilities [31, 32]. We present Deep Agentic Variant Prioritisation (DAVP), an agentic AI system performing patient-specific variant evaluation through dynamic evidence synthesis. DAVP employs three components: (i) `prelimin8` for intelligent gene pre-screening matching phenotypes to gene-disease associations; (ii) `inGeneTopMatch` leveraging semantic knowledge graphs for multi-hop reasoning over gene-phenotype-disease relationships; and (iii) `elimin8` implementing iterative pairwise comparisons incorporating population frequency, predictions, functional evidence, and patient factors. The system adapts evidence-gathering to each patient’s presentation, accesses current literature through retrieval-augmented generation, and maintains transparent reasoning chains documenting prioritization logic. DAVP has the potential to transform variant prioritization from a computationally intractable bottleneck into a transparent, patient-centered diagnostic process.

## Results

### Overview of Deep Agentic Variant Prioritization (DAVP)

DAVP is implemented as a hierarchical multi-stage pipeline that progressively filters and ranks variants using both deterministic and LLM-based components (Fig. 1). Starting from the full variant set in the input VCF, an initial gene-level ranking is used to select the top 256 genes for downstream analysis. DAVP is agnostic to the choice of pre-filter; we adopt Exomiser [23] for the experiments reported here because it dominates the cost-vs-recall Pareto frontier among the eight rankers we evaluated (Supplementary Section S6), but any score-based gene-level ranker can drop in at a quantified recall cost. The *Prelimin8* stage then applies an octant-based elimination procedure (Supplementary Algorithm S1) over these candidates, using LLM-guided tournaments to reduce the gene set to 64 genes. For these 64 genes and their associated variants, *inGeneTopMatch* constructs detailed variant reports by querying a large biomedical knowledge graph and aggregating ontology links, ClinVar and GWAS evidence, and gene-level cache entries. The *Elimin8* stage applies the same octant-based elimination strategy at the variant level to obtain a reduced set of 20 variants. Finally, a *Final Tournament* applies a single-elimination tournament sort (Supplementary Algorithm S2) over these 20 variants, using LLM pairwise comparisons conditioned on variant reports and clinical context to produce a ranked list of the top three candidate variants. The overall computational characteristics of these stages, including LLM call counts and sequential rounds, are summarised in Supplementary Table S1.

**Fig. 1.**
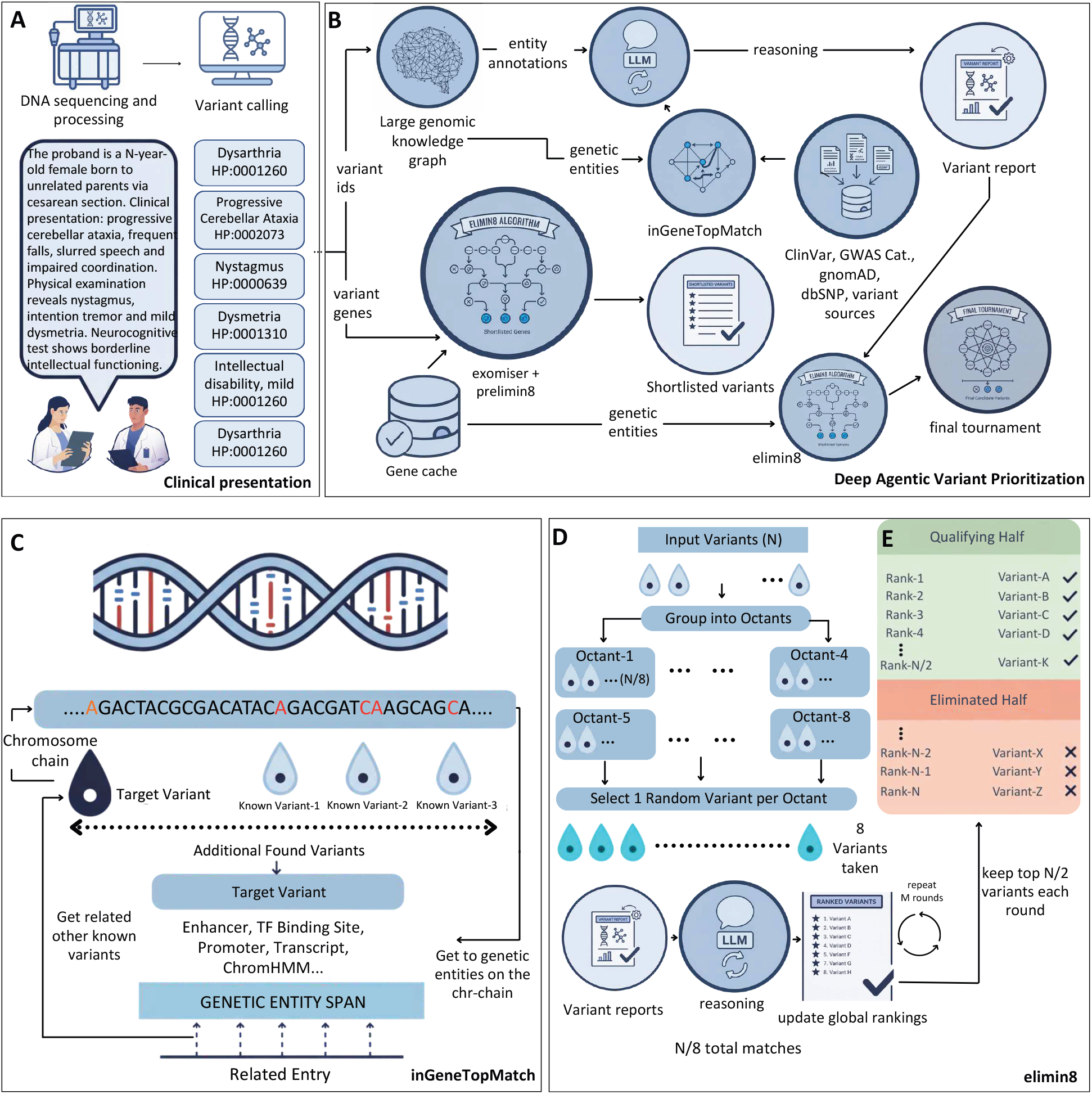
Deep Agentic Variant Prioritization (DAVP) system architecture and workflow. **A)** Clinical data processing pipeline showing DNA sequencing, variant calling, and phenotype extraction from a representative case of an N-year-old female with progressive cerebellar ataxia. Clinical features are standardized to Human Phenotype Ontology (HPO) terms. **B)** Core DAVP architecture integrating multiple components: a large genomic knowledge graph containing genetic entities, LLM-powered reasoning module, inGeneTopMatch algorithm for variant-gene association, external databases (ClinVar, GWAS Catalog, gnomAD, dbSNP), and the elimin8 tournament-style elimination algorithm with final single-elimination tournament selection to generate prioritized variant reports. **C)** Detailed view of the elimin8 algorithm showing iterative variant selection across *M* rounds, with octant-based grouping and progressive refinement to *N/*8 matches while updating global rankings. **D)** Tournament elimination process demonstrating the binary selection mechanism where *N/*2 variants are eliminated in each round, progressively refining the candidate list from initial qualifying variants to final prioritized results.

### DAVP Enables Expert-Level Diagnosis

We evaluated DAVP and three baseline phenotype-driven methods (Exomiser, LIRICAL, Xrare) across four datasets comprising 709 diagnosis cases in total: 63 diagnosis cases from the real-world LIRDB-47 cohort and 646 simulated cases from three independent benchmarks (Simulation, MyGene2-based, and UDN-based, which is characterised by novel and limited-evidence genes; Table S2). For each case, we recorded the rank of the known causal gene and causal variant and computed cumulative distribution functions (CDFs) and top-*k* recall values.

Figure 2A shows the aggregated CDFs of causal variant (top) and causal gene (bottom) ranks over all 709 diagnosis cases. At the gene level, DAVP attains a top-1 recall of 69.39% and a top-3 recall of 85.61%, increasing to 91.11% and 93.09% at *k* = 10 and *k* = 20, respectively (Table S19). The corresponding variant-level recalls are 69.25% (top-1), 85.47% (top-3), 90.83% (top-10), and 92.67% (top-20) (Table S20). The DAVP CDF curves in Fig. 2A saturate by approximately *k* = 20 at both the gene and variant levels, with further increases beyond *k* = 20 mainly affecting the tail (top-50 recall 96.19% at the gene level and 95.63% at the variant level). Across the same 709 cases, the baseline methods exhibit lower aggregated recall values; for example, at the gene level, Exomiser, LIRICAL, and Xrare achieve top-3 recalls of 65.44%, 72.36%, and 49.51%, respectively (Table S19), and at the variant level they achieve 64.32%, 63.61%, and 49.22% top-3 recall, respectively (Table S20). Figure 2B summarises these aggregated differences using per-case head-to-head comparisons: over all cases, DAVP ranks the causal variant above Exomiser in 37.38% of cases and below in 13.40%, with 49.22% ties; analogous win/tie/loss rates versus LIRICAL and Xrare are 32.44/51.48/16.08% and 66.29/25.25/8.46%, respectively (Table S21).

**Fig. 2.**
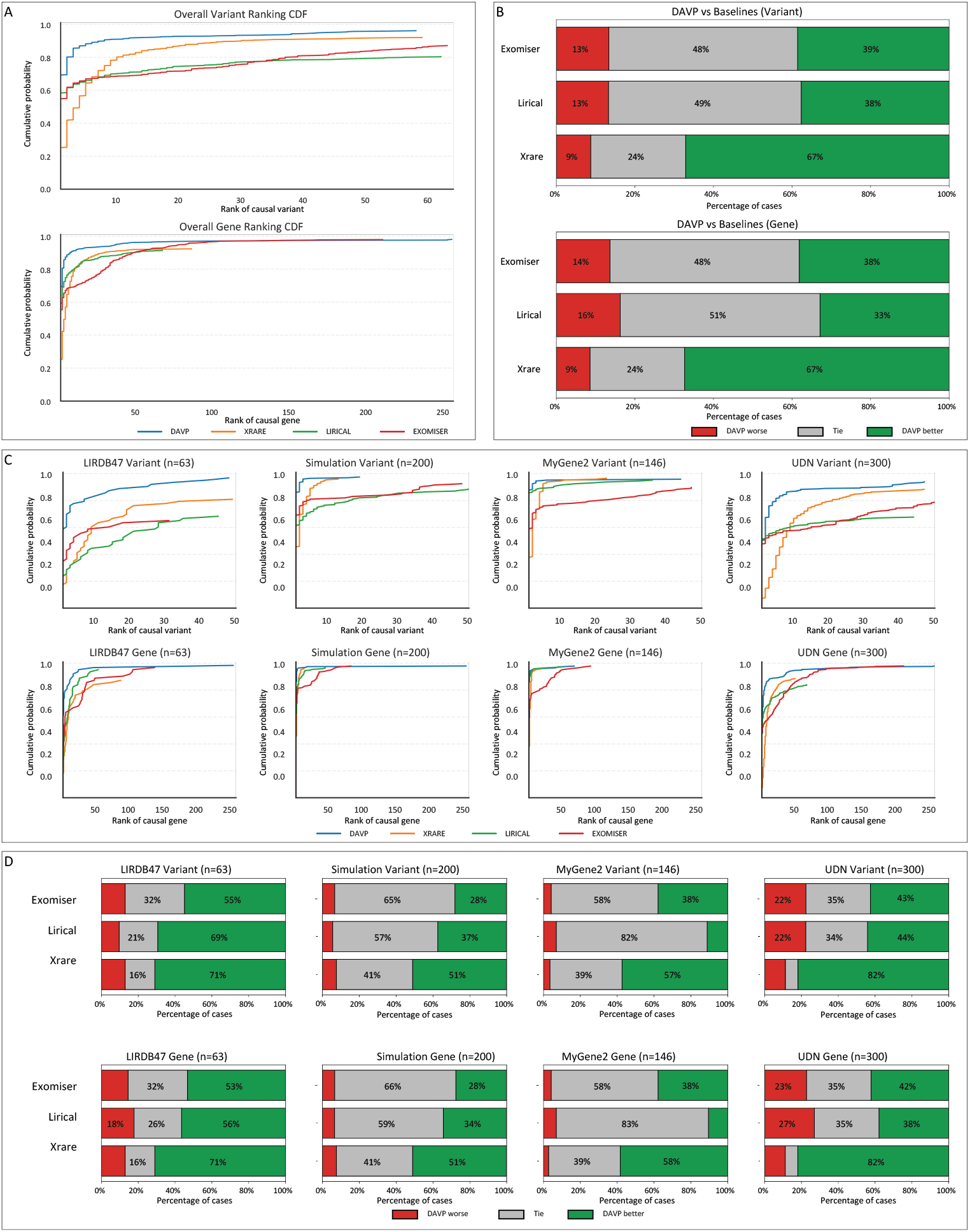
Ranking comparison of DAVP and baseline methods. **(A)** Cumulative distribution functions (CDFs) of causal variant (top) and causal gene (bottom) ranks aggregated across all four datasets (*n* = 709 diagnosis cases). **(B)** Per-case pairwise comparisons between DAVP and Exomiser, LIRICAL, and Xrare at the variant (top) and gene (bottom) levels. **(C)** Variant- and gene-level ranking CDFs stratified by dataset (LIRDB-47: *n* = 63; Simulation: *n* = 200; MyGene2: *n* = 146; UDN: *n* = 300). **(D)** Per-dataset pairwise comparisons between DAVP and each baseline method.

We next examined method performance by dataset (Fig. 2C,D; Tables S19, S20, S21). On the Simulation dataset (200 cases), DAVP achieves gene-level top-3 and top-20 recalls of 96.00% and 97.50%, and variant-level top-3 and top-20 recalls of 96.00% and 97.50% (Tables S19, S20). On the MyGene2-based dataset (146 cases), DAVP reaches gene-level top-3 and top-20 recalls of 94.52% and 95.89%, and variant-level top-3 and top-20 recalls of 94.52% and 95.21%. On LIRDB-47 (63 diagnosis cases), DAVP yields gene-level top-3 and top-20 recalls of 77.78% and 92.06%, and variant-level top-3 and top-20 recalls of 76.19% and 88.89%. On the UDN-based dataset (300 cases), DAVP attains gene-level top-3 and top-20 recalls of 76.00% and 89.00%, and variant-level top-3 and top-20 recalls of 76.00% and 89.00%. In all four datasets, the DAVP CDF curves in Fig. 2C show a rapid rise within the top-20 ranks, with differences across datasets mainly reflected in the fraction of cases solved within the top-3 and top-10 positions. The UDN-based dataset exhibits lower absolute recall values for all methods compared to the Simulation and MyGene2 datasets (Fig. 2C; Tables S19, S20).

Per-dataset head-to-head comparisons (Fig. 2D; Table S21) further characterise ranking behaviour. For example, in LIRDB-47, DAVP ranks the causal variant above Exomiser in 52.38% of cases (33.33% ties, 14.29% losses) and above Xrare in 69.84% of cases (17.46% ties, 12.70% losses). In the UDN-based dataset, DAVP ranks the causal variant above Xrare in 80.33% of cases (8.67% ties, 11.00% losses), and above Exomiser and LIRICAL in 41.33% and 37.33% of cases, respectively (Table S21). Across all datasets, DAVP exhibits a higher fraction of wins than losses against each baseline method in these pairwise comparisons, while the fraction of ties reflects the many cases where methods assign the same ranks to the causal variant.

We also quantified the computational characteristics of the DAVP pipeline. As summarised in Supplementary Table S1, both the Prelimin8 and Elimin8 stages issue (*N* − *K*)/4 eight-way ranking calls for an input of *N* items and target size *K*, over ⌈log_*2*_(*N/K*)⌉ sequential rounds; *inGeneTopMatch* issues *N* report-generation calls in a single parallel round; and the Final Tournament issues 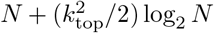 pairwise-comparison calls over *k*_*top*_ · ⌈log_*2*_ *N*⌉ rounds. For the default configuration (top-256 Exomiser genes, Prelimin8 reduction to 64 genes, Elimin8 reduction to 20 variants, top-3 final tournament), the measured per-sample mean over the 709-case benchmark is 42.9 Prelimin8, 81.3 inGeneTopMatch, 33.6 Elimin8 and 27.5 Final Tournament calls, totalling 181.1 LLM calls per sample (Supplementary Table S12).

### DAVP Performance on Uncertain Variants and Across Clinical Subspecialties

To characterise performance on variants with different levels of prior evidence, we stratified the simulated cases (Simulation, MyGene2-based, UDN-based) by ClinVar label of the causal variant into three groups: pathogenic, likely pathogenic, and variants of uncertain significance (VUS). In total, 238 cases had pathogenic causal variants, 246 had likely pathogenic variants, and 160 had variants of uncertain significance (comprising 121 uncertain and 39 conflicting interpretations) (Table S27). Figure 3A,B show the CDFs of causal variant and gene ranks for the VUS subset (panel A) and the pathogenic/likely pathogenic subset (panel B). For VUS causal variants (Table S26), DAVP attains gene-level top-1, top-3, and top-20 recalls of 41.25%, 69.38%, and 85.62%, respectively, and variant-level recalls of 41.25%, 69.38%, and 85.62% at the same *k* values (Tables S26, S27). For pathogenic causal variants (*n* = 238), DAVP reaches gene-level top-1, top-3, and top-20 recalls of 80.7%, 93.3%, and 96.6%; for likely pathogenic causal variants (*n* = 246), the corresponding recalls are 79.7%, 91.5%, and 95.5% (Table S27). Across review-status strata, DAVP gene-level top-3 recall is 86.4% for variants with 0–1 ClinVar stars (*n* = 147), 82.7% for 2-star variants (*n* = 306), and 92.2% for variants with 3–5 stars (*n* = 193) (Table S27). The VUS and low-review-status subsets thus exhibit lower absolute recall values and slower CDF growth than the pathogenic and high-review subsets (Fig. 3A,B; Tables S26, S27), while maintaining the same qualitative behaviour.

**Fig. 3.**
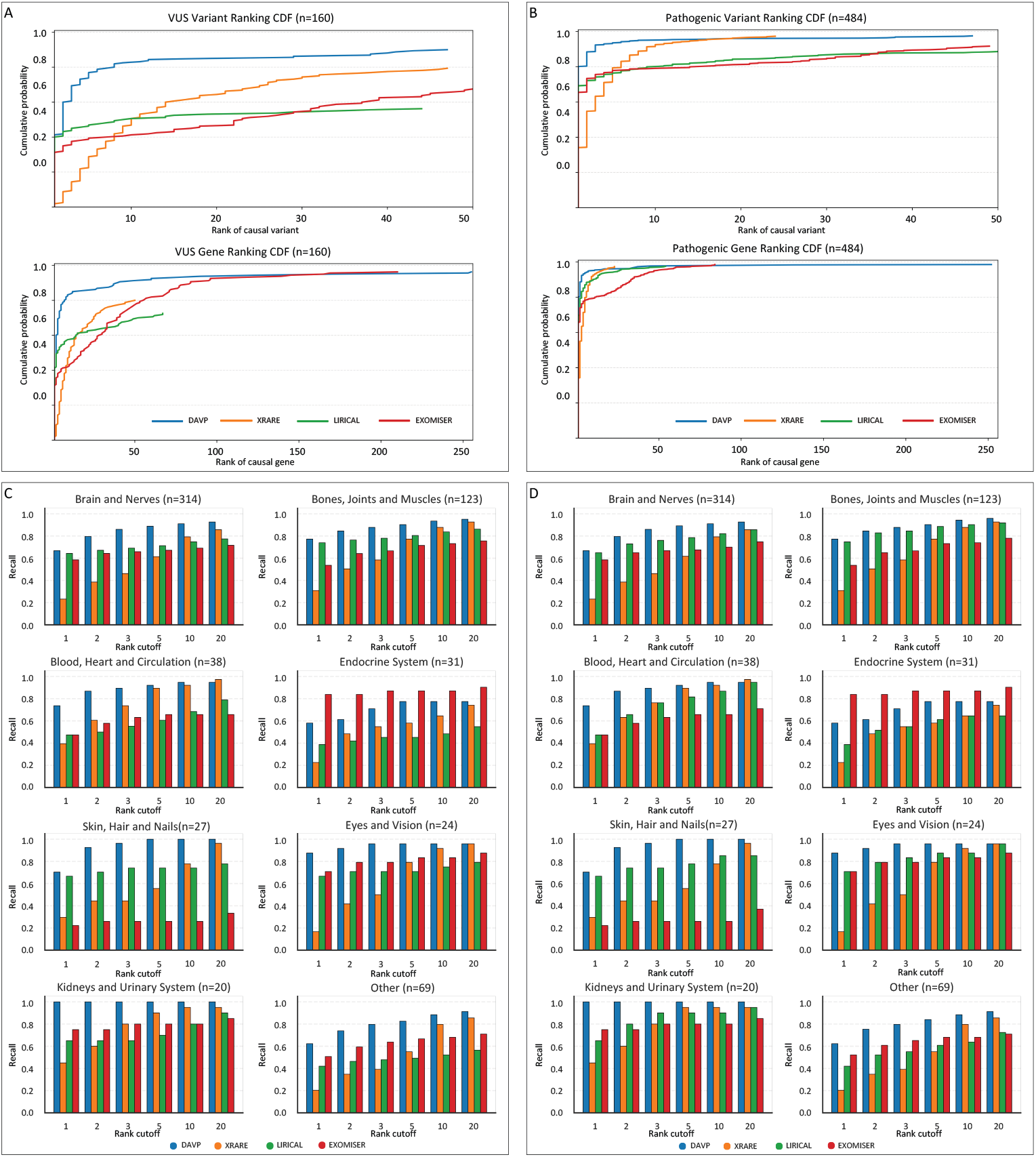
Stratified analysis by ClinVar classification and organ system. **(A)** CDFs of causal variant (top) and causal gene (bottom) ranks for cases with ClinVar VUS causal variants (*n* = 160). (B) CDFs for cases with pathogenic or likely pathogenic causal variants (*n* = 484). **(C)** Variant-level top-*k* recall stratified by organ system. **(D)** Gene-level top-*k* recall stratified by organ system.

We next examined performance across clinical subspecialties using the primary organ system labels assigned to each case. Fourteen categories are represented, with sample sizes ranging from 3 (Female Reproductive System) to 314 (Brain and Nerves) diagnosis cases (Table S7). Figure 3C,D summarise the top-*k* variant-level and gene-level recalls by organ system, and Table S7 reports top-3 and top-20 diagnosis recall values. For example, in the Brain and Nerves category (*n* = 314), DAVP attains top-3 and top-20 diagnosis recall of 85.99% and 92.68%, respectively; in Bones, Joints and Muscles (*n* = 123), these recalls are 87.80% and 95.93%; in Skin, Hair and Nails (*n* = 27), 96.30% and 100.00%; and in Digestive System (*n* = 19), 63.16% and 84.21%. Smaller categories show more variability due to sample size; for instance, Female Reproductive System (*n* = 3) has 100.00% recall at both top-3 and top-20, while Mouth and Teeth (*n* = 9) has 66.67% and 77.78% (Table S7). Across all 14 organ systems, DAVP achieves top-3 diagnosis recall above 63% and top-20 recall above 75% (Table S7).

We also analysed the relationship between phenotype richness and gene-level ranking. Supplementary Figure S7 shows top-10 gene recall grouped by the number of HPO terms per patient (bins: 1–5, 6–10, 11–20, *>*20), aggregated across all datasets. For DAVP, top-10 gene recall in these bins is 89.6%, 90.6%, 93.7%, and 92.7%, respectively (from Supplementary Fig. S7). Baseline methods exhibit lower recalls in the same bins; for example, Xrare’s top-10 gene recall ranges from 65.9% (1–5 HPO terms) to 84.0% (>20 terms), LIRICAL from 74.2% to 84.0%, and Exomiser from 67.1% to 74.7%. The dataset-level HPO distributions underlying these analyses are shown in Supplementary Figure S2 (top left), where the UDN-based dataset displays the highest median HPO term count and the LIRDB-47 cohort the lowest. Supplementary Figure S2 (top right) also reports the distribution of candidate variants per patient, with LIRDB-47 having the largest median variant count among the four datasets.

### DAVP Rankings Are Biologically Grounded and Not Driven by Any Single Annotation Source

To examine how DAVP rankings relate to biological and annotation-based features, we analysed several properties of genes and variants as a function of their DAVP rank.

First, we quantified gene-level HPO ontology-based similarity for each case by computing the similarity between the patient’s HPO profile and the set of HPO terms associated with each gene. Figure 4A shows the distribution of these similarity scores grouped by DAVP gene rank bins (1–3, 4–10, 11–50, >50). The similarity distributions for ranks 1–3 and 4–10 are shifted towards higher values relative to the 11–50 and >50 bins, with the highest-density regions for the 1–3 bin lying in the upper portion of the plot. In contrast, genes ranked >50 exhibit a broader and lower similarity distribution, with a larger fraction of low-similarity values. These distributions indicate that the genes placed in the top positions by DAVP tend to have higher annotation-based HPO similarity to the patient phenotype profile than genes placed at lower ranks, as directly measured by the ontology-based similarity metric used in Fig. 4A.

**Fig. 4.**
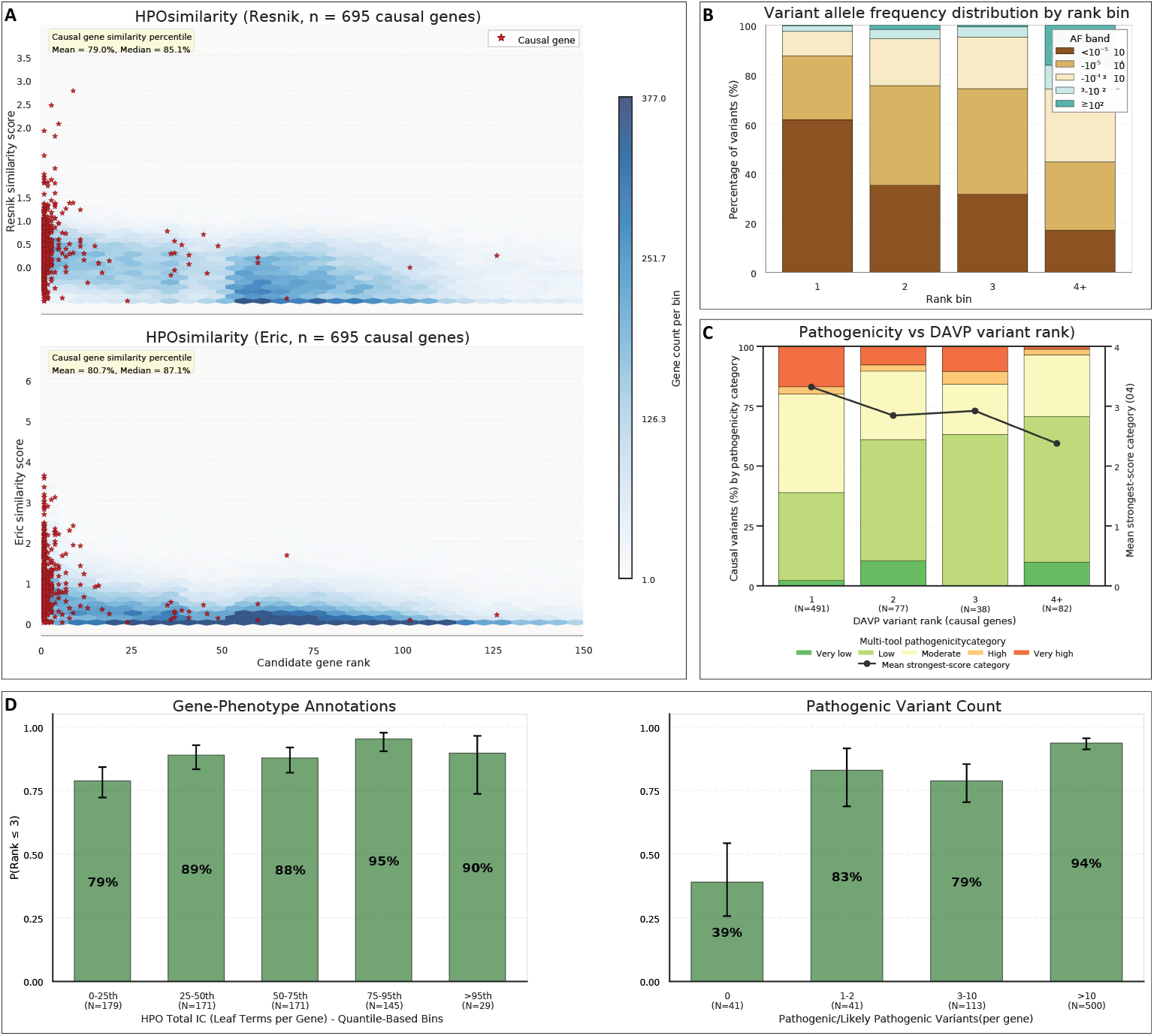
Relationship between DAVP rankings and biological features. **(A)** Distribution of HPO semantic similarity scores across DAVP gene rank bins. **(B)** Population allele frequency distributions stratified by DAVP variant rank. **(C)** Aggregate pathogenicity scores across DAVP rank categories from eight independent scoring systems: CADD, AlphaMissense, SIFT, PolyPhen-2, REVEL, DANN, MetaLR, and ACMG severity criteria. Scores from each tool were binned into categories 1–5 (1=benign, 5=pathogenic). Colored bars show the mean binned score for each tool across samples at each rank. The black line shows the mean of the maximum pathogenic prediction across all eight tools for each sample. **(D)** Top-3 recall stratified by gene annotation density (HPO information content and ClinVar pathogenic variant count).

Second, we examined population allele frequencies of DAVP-ranked variants using gnomAD-derived frequencies. Figure 4B and Supplementary Table S28 show the allele frequency distributions for variants at rank 1, rank 2, rank 3, and ranks 4+. Among rank-1 variants, 80.36% have allele frequency < 10^−5^, 13.24% fall between 10^−5^ and 10^−4^, 5.06% between 10^−4^ and 10^−3^, 1.19% between 10^−3^ and 10^−2^, and only 0.15% have allele frequency > 0.01 (Table S28). For rank-2 variants, the corresponding proportions are 50.89% (< 10^−5^), 30.51% (10^−5^–10^−4^), 14.43% (10^−4^–10^−3^), 2.83% (10^−3^–10^−2^), and 1.34% (> 0.01). Rank-3 variants show 45.39%, 34.08%, 16.67%, 3.27%, and 0.60% across the same bins. For variants ranked 4 or lower, the distribution shifts towards higher frequencies: 23.31% (< 10^−5^), 27.42% (10^−5^–10^−4^), 28.06% (10^−4^–10^−3^), 8.80% (10^−3^–10^−2^), and 12.42% (> 0.01). Thus, the top-ranked variants are enriched for very rare alleles (frequency < 10^−5^), whereas variants at lower ranks include a larger fraction of more common alleles (Fig. 4B; Table S28).

Third, we evaluated how DAVP ranks relate to multiple independent pathogenicity prediction frameworks. For each variant, we collected categorical pathogenicity scores from eight tools (CADD, AlphaMissense, SIFT, PolyPhen-2, REVEL, DANN, MetaLR, and ACMG-based severity) and binned them into five categories (1=benign, 5=pathogenic). Figure 4C shows, for each DAVP rank bin, the mean binned score for each tool (coloured bars) and the mean of the maximum score across all tools (black line). The maximum-score curve is highest for the 1–3 rank bin and gradually decreases across the 4–10 and 11–50 bins, with the lowest values in the >50 bin. Individual tools display heterogeneous behaviour; for instance, some tools show less separation between ranks than others, and the coloured bars for any given bin span a range of mean values. This indicates that no single predictor correlates well with the DAVP ranking, although the aggregate maximum score across tools shows a slightly better correlation (Fig. 4C).

Fourth, we analysed DAVP performance as a function of gene annotation density. We used two proxies for annotation density: (i) the HPO information content of each gene (sum of information content of all associated HPO terms) and (ii) the number of ClinVar pathogenic variants previously reported for that gene (Methods). Figure 4D shows top-3 gene-level recall across bins of these measures. For example, when stratifying by ClinVar pathogenic variant count into three groups (0, 1–5, > 5), DAVP achieves similar top-3 recalls across bins (Fig. 4D), with differences of only a few percentage points. When stratifying by ClinVar review status of the causal variant (0–1, 2, 3–5 stars), gene-level top-3 recall ranges from 82.7% to 92.2% (Table S27). When stratified by HPO information content, top-3 gene-level recall falls within a narrow range, and when stratified by ClinVar pathogenic-variant count, it likewise falls within a mostly narrow range.

Finally, we examined dataset-level structure in the clinical representations used by DAVP. Supplementary Figure S2 (bottom row) displays a two-dimensional UMAP projection of patient embeddings derived from the clinical epicrises. When coloured by dataset (bottom left), MyGene2-based cases form a visually compact group, while Simulation, UDN-based, and LIRDB-47 cases occupy overlapping but partially separated regions. When coloured by disease category (bottom right), cases with similar organ-system labels tend to appear near each other, producing visible clusters corresponding to major categories such as neurological and musculoskeletal disorders. Supplementary Figure S2 (top row) complements these views with distributions of HPO term counts and candidate variant counts per patient, which differ across datasets as noted above and help contextualise the ranking behaviour observed in Figures 2 and 3.

## Discussion

Systems like DAVP require continued development to address three critical challenges in clinical genomics: context-dependent variant causality, knowledge accumulation quality, and polygenic trait interpretation. First, context-dependent causality remains fundamentally challenging despite advances in variant pathogenicity prediction. Recent population-based studies reveal that ClinVar pathogenic variants exhibit only 7% mean penetrance in biobanks compared to 90%+ in family studies [33, 34], while the same genetic variants can produce dramatically different phenotypes depending on genetic background, sex, ancestry, tissue context, and environmental factors [35]. Current ACMG/AMP guidelines [17], designed for highly penetrant Mendelian disorders, fail to capture this complexity through their dichotomous classification framework, and even state-of-the-art AI systems like AlphaMissense [28] and EVE [36] provide disease-agnostic pathogenicity scores without accounting for the tissue-specific, developmental, and genetic background effects that determine actual clinical outcomes. The inherent biological reality that penetrance is not a fixed parameter but varies across contexts, exemplified by HbS variants protective in malaria-endemic regions yet harmful elsewhere, or X-chromosome variants with female-to-male ratios exceeding 11:1 [35], necessitates moving beyond static variant classifications toward context-aware probabilistic frameworks that integrate modifier genes, epigenetic factors, and phenotype-specific effects.

Secondly, knowledge accumulation through literature and databases suffers from substantial quality issues that hinder reliable interpretation. Current data reveals 41% of individuals undergoing genetic testing receive at least one variant of uncertain significance (VUS) [10], with only 7.3% of VUSs reclassified over an 8-year period (mean time to reclassification: 2.5 years), while 5.7% of ClinVar variants harbour conflicting interpretations between submitters [37]. Error propagation through literature remains pervasive: studies demonstrate 33–43% annotation error rates in curated databases [38], and when common variants initially classified as pathogenic are re-evaluated, 40% are downgraded [39], with older classifications (pre-2014) showing 67% downgrade rates and literature-only sources reaching 63%. These errors cascade through citation networks and database-to-database transfers, creating persistent “zombie” variants that maintain incorrect classifications despite contradictory evidence. Ancestry bias further compounds the problem, with VUS rates varying inversely with genomic database representation (Pacific Islander individuals showing highest rates) and historical false-positive rates 5.8-fold higher for African ancestry in HGMD data (2014) [40]. The fundamental issue is that causality relations formed through scientific literature lack systematic validation mechanisms, and while database quality has improved dramatically since 2015 (ClinVar achieving 97% reduction in false positives) [40], the slow reclassification timelines and low resolution rates mean knowledge accumulation lags far behind discovery, with clinical evidence showing 18–55× enrichment in successful reclassifications yet remaining scarce for most variants [41]. It is precisely in this challenging landscape that dynamic, agentic approaches like DAVP may add value: by synthesizing evidence across multiple sources rather than relying on any single pre-computed score, such systems can provide meaningful prioritization even when curated annotations are incomplete or conflicting. Future systems must therefore couple agentic reasoning with active learning frameworks that prioritize functional validation of high-impact uncertain variants, develop automated literature curation with provenance tracking to identify error propagation chains, and establish continuous evidence aggregation pipelines that update variant classifications as new data emerges rather than relying on sporadic manual re-evaluation.

Finally, extending these approaches to polygenic trait fine-mapping and disease mechanism hypothesis formation presents distinct challenges and opportunities. Complex traits fundamentally differ from Mendelian diseases through extreme polygenicity (10,000–100,000+ causal variants per trait), small effect sizes (typical OR 1.01–1.1 versus OR > 10 for Mendelian variants), regulatory complexity (90%+ signals in noncoding regions), and tissue-specific context-dependency that current methods struggle to capture. The causality gap between statistical association and biological mechanism remains vast: linkage disequilibrium creates irreducible uncertainty (median credible set size: 5 SNPs), functional validation is resource-intensive, and the regulatory context determining which genes are affected by non-coding variants remains largely opaque. Emerging AI and computational systems offer complementary capabilities: knowledge graphs like BioKDE [42] and GenomicKB [43] demonstrate automated hypothesis generation that could have predicted 40% of lung cancer drug targets ∼15 years early through probabilistic semantic reasoning over causal relationships; deep learning systems like SHEPHERD [44] enable individualized rare disease diagnosis by learning from simulated patients; and causal machine learning frameworks are beginning to distinguish direct regulatory effects from confounded correlations in single-cell perturbation data. However, these systems exhibit critical limitations: variant pathogenicity predictors like AlphaMissense [28] provide population-scale coverage but lack disease and tissue specificity; knowledge graphs aggregate data without true causal learning and require extensive manual curation; causal inference methods struggle with unmeasured confounders and scalability to genome-wide analyses [45]. Our ablation studies suggest one productive direction: prolonged reasoning depth appears more effective than simply increasing the volume of iterative comparisons, with enabling the agentic thinking budget yielding the largest performance gains (55% relative improvement) in our experiments. The path forward requires integrating these disparate approaches into unified platforms that combine variant effect prediction with phenotype-driven prioritization, knowledge graph reasoning for mechanism hypotheses, causal inference for regulatory relationships (leveraging single-cell perturbation screens), and continuous learning from clinical outcomes to refine predictions. Specifically, systems should move from static variant classifications to dynamic probabilistic models that update with accumulating evidence, from correlation-based associations to causal mechanistic models validated through perturbation data, and from single-scale analysis to multiscale integration spanning molecular effects through cellular phenotypes to organismal disease.

## Materials & Methods

### Datasets and Benchmarks

We evaluated DAVP on 709 diagnosis cases across four benchmarks: three simulated cohorts (Custom Simulation, MyGene2-based, and UDN-based; 646 cases total) and one real-world cohort (LIRDB-47; 63 cases from 47 Turkish rare-disease patients). A separate 100-case validation set was used for prompt construction and configuration selection only and is excluded from all reported evaluations. Per-cohort summary statistics are provided in Supplementary Table S2.

#### Validation set

We sampled 100 individuals from the 1000 Genomes Project [46] and generated simulated cases over them using the same pipeline described under *Cohort construction* below. This set was used solely to refine in-context prompts, select algorithmic configurations, and choose base models; all adjustments were made through in-context specification rather than gradient-based learning. These cases are excluded from every reported evaluation.

#### Simulated benchmarks

The *Custom Simulation* cohort (*n* = 200) draws background genomes from the 1000 Genomes Project [46] and spikes in pathogenic or likely pathogenic causal variants from three consecutive ClinVar [47] VCF releases (2023-12-30, 2024-12-30, 2025-09-28), with 80% of causal variants drawn from the 2024–2025 window and 20% from the 2023–2024 window; this distribution ensures the majority post-date the 2023 knowledge-graph snapshot consulted by *inGeneTopMatch*. Variants are further stratified 50/50 by ClinVar review status to limit reliance on heavily reviewed entries that may be over-represented in LLM pretraining. The *MyGene2-based* cohort (*n* = 146) is built from the MyGene2 benchmark [48] released with the SHEPHERD framework [44], with each gene-level diagnosis mapped to a ClinVar variant via the gene-to-variant conversion described under *Cohort construction*. The *UDN-based* cohort (*n* = 300) is derived from the benchmark of Alsentzer et al. [49] and is deliberately split into 150 *UDN-Pathogenic* cases (catalogued causal gene with a prior ClinVar pathogenic or likely pathogenic annotation) and 150 *UDN-VUS* cases (novel causal gene with only a VUS or conflicting ClinVar interpretation), stress-testing the evidence-poor end of the rare-disease spectrum. Each case additionally inherits the false-positive candidate variants from the original UDN diagnostic workup as realistic distractors that survived expert curation. A per-sub-cohort breakdown and a distractor-stratified analysis of DAVP’s ranking behaviour on this cohort are provided in Supplementary Section S11.

#### Real-world benchmark: LIRDB-47

LIRDB-47 (*Lidya InterGen Rare Disease Benchmark*) comprises 47 clinically confirmed rare-disease patients from the Intergen Genetic and Rare Diseases Diagnosis and Research Center in Turkey; 12 patients carry two or more independently diagnosed causal variants, yielding 63 evaluation cases that are scored separately throughout. Each case includes a clinically validated causal variant, a de-identified clinician-curated narrative, and HPO terms assigned by the treating clinician, making LIRDB-47 the only benchmark in this study whose phenotype labels are unfiltered human-expert output. The cohort uses Intergen’s production exome-sequencing pipeline and the same variant-annotation schema as the simulated benchmarks, so the prioritiser input format is identical across all four datasets. Ethics approval and informed consent for de-identified release were obtained through Intergen.

#### Cohort construction

For the three simulated cohorts we generate synthetic clinical narratives in three stages: a grounded-search language model is prompted with the causal gene–disease pair to retrieve common and rare symptoms and demographic predilections from indexed literature; a reasoning model then composes a patient vignette under a structured-output schema (age, sex, optional ethnicity, free-form symptom list); and the rendered epicrisis is processed by a retrieval-augmented HPO-extraction pipeline that embeds atomic clinical observations against a ChromaDB index of HPO term descriptions. For MyGene2 and UDN the original benchmarks supply HPO annotations directly, restricted to “observed” terms; for LIRDB-47 the clinician-authored epicrisis and HPO list are retained verbatim. Gene-to-variant mapping for the gene-based cohorts (MyGene2, UDN) uses canonicalised disease-label fuzzy matching against ClinVar with progressive relaxation of significance and match-score thresholds. The full procedure—variant selection windows, perturbation strategy, the three-stage narrative pipeline, and the HPO-RAG configuration—is described in Supplementary Section S2.

### DAVP Pipeline

#### Problem Definition

For each patient we receive a 5-tuple (*V, P, c*, **X**, *G*), where *V* = {*v*_*1*_, &, *v*_*m*_} is the set of observed variants, *P* = {*p*_*1*_, &, *p*_*k*_} a set of Human Phenotype Ontology (HPO) terms, *c* a free-text clinical epicrisis, **X** = [**x**_*1*_, &, **x**_*m*_] a stacked annotation matrix in which each **x**_*i*_ ∈ ℝ^*d*^ encodes population allele frequencies, ACMG criteria, in silico pathogenicity predictors, molecular consequence, and gene-level context, and *G* = {*g*_*1*_, &, *g*_*n*_} the gene set induced by *V* (with *V*_*j*_ = {*v*_*i*_ ∈ *V*: gene(*v*_*i*_) = *g*_*j*_}). The goal is to identify a causative subset *V* ^⋆^ ⊆ *V* explaining *P* and to produce an ordering *v*_*(1)*_ ≻ *v*_*(2)*_ ≻ · · · ≻ *v*_(|*V*_ ⋆_|)_. The pipeline does not assume a single numeric pathogenicity score; instead it integrates heterogeneous evidence—annotations, phenotype similarity, knowledge-graph context, and LLM-based reasoning—to derive the ranked subset.

#### Filtering

The LLM-driven stages of DAVP can in principle operate over the full candidate set, so the gene-level pre-filter is not a method dependency but an interchangeable cost-control component: the per-sample LLM call budget grows linearly with input size (Supplementary Table S13), and any deterministic ranker that retains the answer gene with high probability suffices to cap inference cost. We characterise this cost-vs-recall trade-off systematically in Supplementary Section S6, evaluating nine gene-level rankers—three phenotype-aware (Exomiser with HPO, LIRICAL, Xrare) and six phenotype-agnostic (Exomiser without HPO, ACMG severity, CADD, REVEL, AlphaMissense, DANN)—across the cut-off grid *K* ∈ {8, 16, 32, 64, 128, 256, 512, 1024}. Exomiser [23] dominates the resulting Pareto frontier (Supplementary Table S15, Supplementary Figure S4), so we adopt it as the operational default for all reported experiments; any of the alternatives drops in at a quantified recall cost. For each gene *g*_*j*_ ∈ *G* we retrieve its Exomiser score and retain the top *N*_*g*_ = 256 genes, 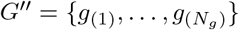, with corresponding variant set *V* ^*′′*^ = {*v*_*i*_ ∈ *V*: gene(*v*_*i*_) ∈ *G*^*′′*^}. The cutoff *N*_*g*_ = 256 sits at the knee of the Exomiser-with-HPO recall-vs-cost Pareto curve (Supplementary Section S6) and retains the causal gene in 97.7% of cases (Supplementary Table S15).

#### Gene Cache

A patient-agnostic *gene cache* stores, for every protein-coding gene *g*, a structured ten-section dossier *c*(*g*) = (*a*_*1*_(*g*), &, *a*_*10*_(*g*)) compiled once via grounded LLM web retrieval over OMIM, HPO, ClinVar, gnomAD, and GTEx; the ten sections cover gene identity and clinical context, constraint and intolerance metrics, phenotype spectrum, genotype–phenotype correlations, representative pathogenic variants, tissue expression, molecular mechanism, diagnostic yield, key clinical literature, and a per-run HPO-matching summary. Because |*G*_*all*_| ≪ |*V* |, the cache is built once and indexed by gene at runtime: if gene(*v*_*i*_) = *g*, then *c*(*v*_*i*_) ≡ *c*(*g*). Across the 709-case benchmark the cache spans 16,311 unique genes with a mean dossier length of approximately 9,200 characters and 94.7% of dossiers containing all ten sections. The schema, per-section presence rates, and aggregate cache statistics are tabulated in Supplementary Tables S8 and S9; a worked example dossier is provided in Supplementary Section S4.

#### Prelimin8

Prelimin8 reduces the candidate gene set from *N*_*g*_ = 256 to *K*_*p8*_ = 64 via an iterated octant-elimination procedure. We initialise a working set *S*^*(0)*^ = *V* ^*′′*^ with scores 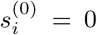. At each iteration *t*, items are sorted by current score and partitioned into 8 ordered groups 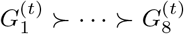; one representative per group is sampled to form a tournament set 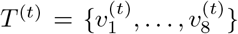; an LLM ranks the eight items and scores are updated by the fixed weight vector **w** = [10, 7, 5, 3, 2, 1, 0, −1], 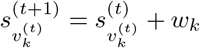, while all other items retain their previous scores; the bottom half of *S*^*(t)*^ is then discarded. The procedure repeats until |*S*^*(t)*^| ≤ *K*_*p8*_. Each LLM call sees the patient context (*P, c*) alongside a compact per-item report *r*_*i*_ = (**x**_*i*_, *c*(*g*_*i*_)) that pairs the variant’s annotations with its gene-cache entry. Pseudocode is given in Supplementary Algorithm S1.

#### inGeneTopMatch

For each variant surviving Prelimin8, inGeneTopMatch builds a Detailed Variant Report by traversing GenomicKB [43], a biomedical knowledge graph 𝒢_*KB*_ = (𝒱,ℰ) with approximately 3.5 × 10^8^ typed nodes and 1.4 × 10^9^ edges spanning genes, transcripts, regulatory elements, ontology terms, GWAS entries, ClinVar records, and 200 bp *chr-chain* segments tiling the genome. We retrieve the genomic entities *E*_*i*_ = {*e* ∈ 𝒱: pos(*v*_*i*_) ∈ span(*e*)} overlapping the variant position—exons, regulatory elements, splice sites, promoters—and for each *e* ∈ *E*_*i*_ collect its ontology annotations together with the ClinVar and GWAS evidence from other variants mapped to the same entity (formal set-builder definitions in Supplementary Section S2). The assembled per-variant document 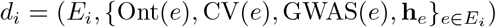) is passed to the LLM together with the patient HPO set *P* and gene-cache entry *c*(*g*_*i*_), producing a Detailed Variant Report *R*_*i*_ = ⟨*v*_*i*_, **x**_*i*_, *c*(*g*_*i*_), *d*_*i*_, LLM(*d*_*i*_, *P*)⟩ that integrates annotation evidence, gene-level context, knowledge-graph signals, and an LLM-derived mechanistic summary. The stage issues exactly one LLM call per surviving variant.

#### Elimin8

Elimin8 re-applies the octant-elimination procedure of Prelimin8 (Supplementary Algorithm S1) to the post-inGeneTopMatch survivors, with one substantive change: each LLM ranking call is conditioned on the full Detailed Variant Reports *R*_*i*_ rather than the compact *r*_*i*_ used by Prelimin8. The richer context lets the LLM weigh knowledge-graph-derived mechanism evidence that is too expensive to materialise across the full Prelimin8 candidate pool. Elimin8 reduces the working set to *K*_*e8*_ = 20 variants.

#### Final Tournament

The Final Tournament resolves the remaining 20 variants into a top-3 ranking via a single-elimination pairwise tournament with iterative top-*k* extraction (Supplementary Algorithm S2)—an algorithm distinct from the octant procedure used by Prelimin8 and Elimin8. Bracket pairings are randomised so that strong variants are spread across the tree rather than meeting in early rounds; within each pair, however, the two variants are presented to the LLM in Elimin8 score order, so the higher Elimin8-ranked variant always appears as item 1 in the comparison prompt. This aligns the well-documented LLM positional preference toward the first option with the upstream Elimin8 signal, turning the bias into a regulariser that breaks ties in favour of the variant the previous stage already preferred. In each round, variants are paired and an LLM compares each pair (*v*_*i*_, *v*_*j*_) conditioned on the patient context (*P, c*) and the two Detailed Variant Reports (*R*_*i*_, *R*_*j*_), selecting a winner; an unpaired variant in an odd round advances automatically. The single winner is rank 1. To extract subsequent ranks *r* ∈ {2, 3} we collect every variant that lost directly to a current top-(*r* − 1) candidate but is not yet ranked and run a fresh sub-tournament over that set; this guarantees that each rank is the strongest variant among those not yet eliminated by a higher-ranked candidate. Each comparison is logged for downstream interpretability, and a clinician-facing summary aggregates the Detailed Variant Reports for the top-3 variants, returning the prioritised list together with the LLM-generated supporting evidence for clinical review.

#### Evaluation

We compare DAVP against three established phenotype-driven prioritisers: Exomiser [23], LIRICAL [50], and Xrare [51]. Performance is reported as top-*k* recall at *k* ∈ {1, 3, 5, 10, 20, 50}, evaluated at both gene level and variant level; per-case head-to-head performance is summarised as win/tie/loss percentages with two-sided Wilcoxon signed-rank significance tests. To isolate the contribution of phenotype-aware reasoning, we additionally compare against five phenotype-agnostic single-score baselines (CADD, REVEL, AlphaMissense, DANN, and an ACMG-severity ranking) in Supplementary Section S7.

#### Computational characteristics

All four LLM stages run Gemini 2.5 Flash with sampling temperature 0.7, output cap 65,536 tokens, and an 8,192-token extended-reasoning (“thinking”) budget; inGeneTopMatch is the sole exception, running with the thinking budget disabled because its task is summarisation rather than ranking. Prelimin8 and Elimin8 each issue (*N* − *K*)/4 eight-way ranking calls over ⌈log_*2*_(*N/K*) ⌉ sequential rounds; inGeneTopMatch issues exactly *N* report-generation calls in a single parallel round; the Final Tournament issues 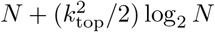 pairwise comparisons over *k*_*top*_ · ⌈log_*2*_ *N*⌉ rounds (Supplementary Table S1). Under the paper configuration this works out to a measured mean of approximately 181 LLM calls per case end-to-end, with per-cohort breakdowns in Supplementary Table S12 and full per-stage hyperparameters in Supplementary Table S10. The independent contribution of each stage is evaluated in the stage-level ablation (Supplementary Figure S6, Table S17); a hyperparameter sweep over base model, rounds-before-elimination, and thinking-budget settings is reported in Supplementary Table S18.

## Data Availability

The source code and data to reproduce this work are available at https://github.com/Muti-Kara/davp

## Data Availability

All four benchmark datasets used in this study are released together with the DAVP source code at https://github.com/Muti-Kara/davp. The Custom Simulation, MyGene2-based, and UDN-based simulated benchmarks are distributed with scripts to regenerate them from the 1000 Genomes Project and ClinVar. For LIRDB-47 we release the causative variants together with the reported HPO terms in de-identified form. Ethics approval for LIRDB-47 was obtained from the Lokman Hekim University Scientific Research Ethics Committee.

## Code Availability

The DAVP source code, including all pipeline stages, prompt templates, gene-cache construction scripts, and analysis notebooks, is available at https://github.com/Muti-Kara/davp under an open-source licence. Specific versions used to generate the results in this paper are tagged in the repository.

## Acknowledgements

We acknowledge the contributions of the employees of Lidya Genomics and specifically thank Can Alkan for fruitful discussions and support with data acquisition. We thank Furkan Karademir for assistance with HPO-RAG support. We also thank MD Ismail Berk Unlu for clinical comments and guidance.

## Author Contributions

FO designed the study. SC and FD curated and provided the data. MK, AFG, SEK, OO and FO designed and implemented the models.

## Rights Retention Statement

This work is supported by Lidya Genomics. For the purpose of Open Access, the author has applied a CC BY-NC-ND public copyright licence to any Author Accepted Manuscript (AAM) version arising from this submission.

## Supplementary Material

### Algorithm 1

Elimin8: Octant-Based Iterative Elimination

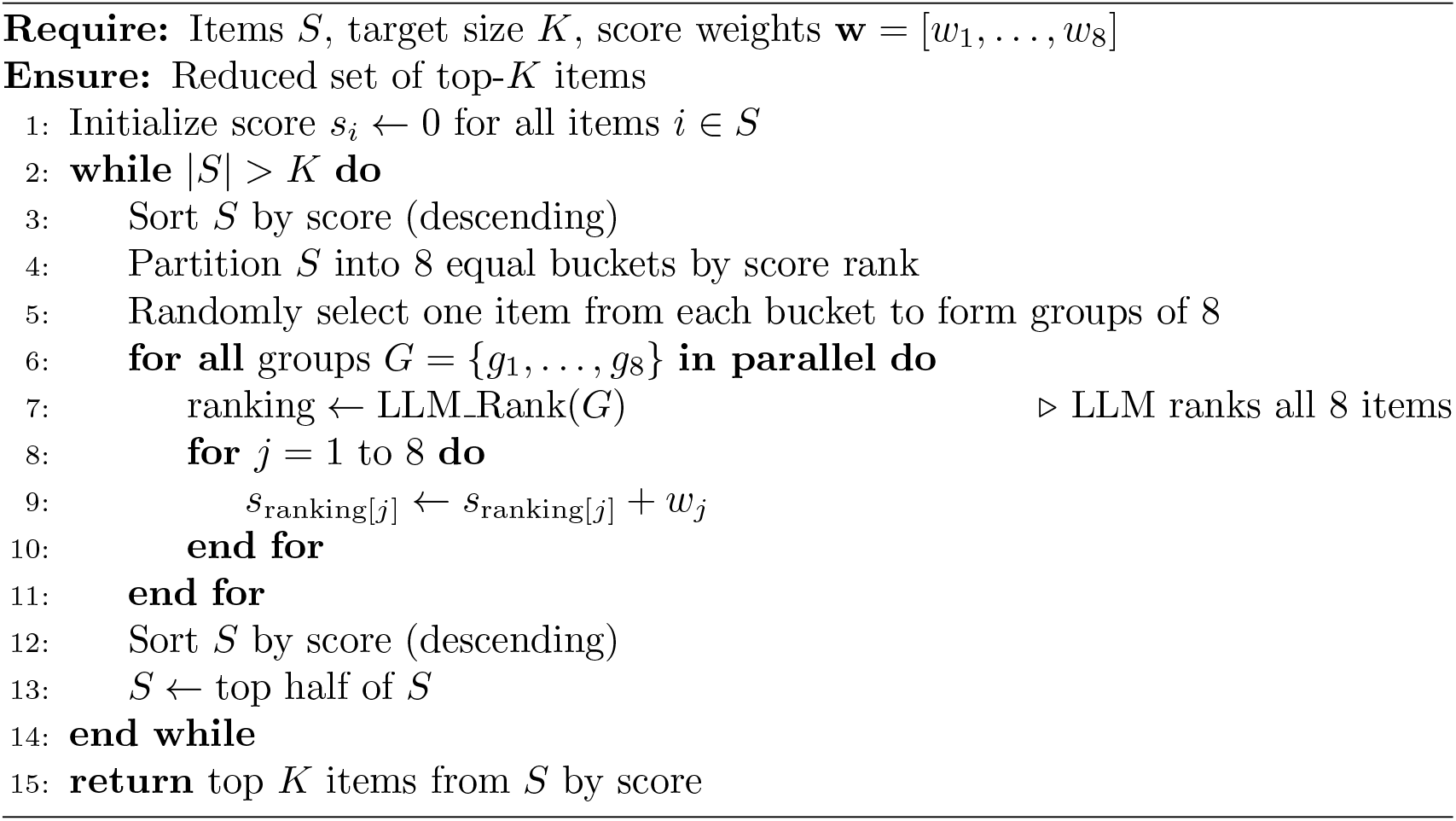

**Table S1:**
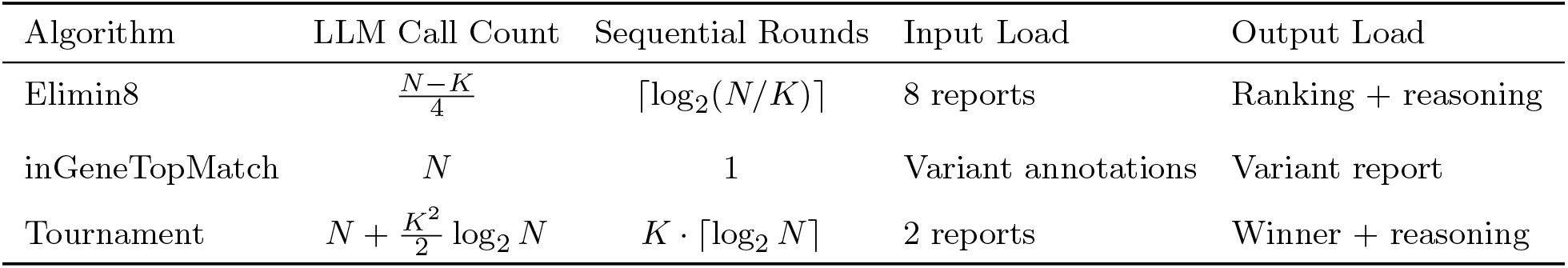
Computational complexity of DAVP algorithms. *N*: input item count, *K*: output item count.

| Algorithm | LLM Call Count | Sequential Rounds | Input Load | Output Load |
| --- | --- | --- | --- | --- |
| Elimin8 | $\frac{N-K}{4}$ | $\lceil \log_2(N/K) \rceil$ | 8 reports | Ranking + reasoning |
| inGeneTopMatch | $N$ | 1 | Variant annotations | Variant report |
| Tournament | $N + \frac{K^2}{2} \log_2 N$ | $K \cdot \lceil \log_2 N \rceil$ | 2 reports | Winner + reasoning |

## S1. Algorithm pseudocode and computational complexity

This section gives the verbatim pseudocode for the two iterative algorithms invoked across the four DAVP stages and tabulates their per-call complexity. The cost-vs-cutoff implications (per-sample LLM call count as a function of the gene-level pre-filter cutoff *N*_*g*_) are reported in Supplementary Section S6, alongside the filter-recall trade-off.

### Algorithm 2

Tournament Sort: Single-Elimination with Top-*k* Extraction

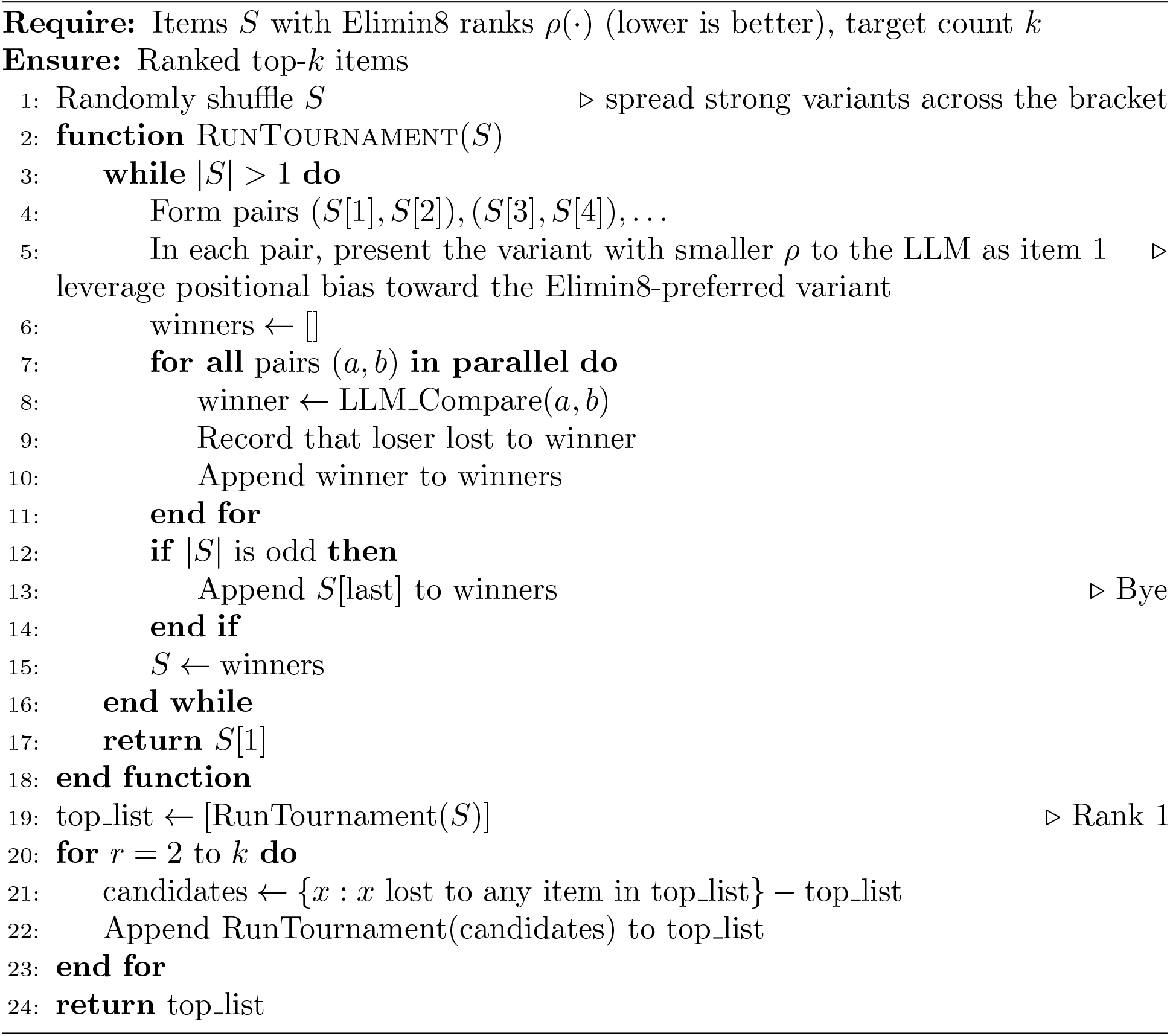

## S2. Extended benchmark methodology

This section expands the Datasets subsection with the cohort-construction details omitted from the main text for space.

### Custom Simulation (*n* = 200)

For each case a single 1000 Genomes Project individual [46] contributes the background VCF and a single ClinVar [47] variant is spiked in as the causal allele. No individual is reused. Causal variants are drawn from three consecutive ClinVar releases (2023-12-30, 2024-12-30, 2025-09-28); 80% from variants first appearing between the 2024 and 2025 releases and 20% from those appearing between 2023 and 2024, ensuring the majority post-date the 2023 knowledgegraph snapshot consulted by *inGeneTopMatch*. To counter LLM pretraining bias toward heavily reviewed entries, we stratify 50/50 by ClinVar review status (0–2 vs. 3–4 stars). Only pathogenic and likely pathogenic variants are selected as causal. Every non-causal variant in the candidate list is a real call from the background individual; no read-level simulation is performed.

### MyGene2-based simulation (*n* = 146)

We start from the MyGene2 benchmark released with the SHEPHERD framework [44]. MyGene2 supplies gene-level diagnoses without causal variants, so we apply the gene-to-variant conversion described below to assign a ClinVar variant to each case.

### UDN-based simulation (*n* = 300)

We derive the UDN simulation from the benchmark of Alsentzer et al. [49], slicing the cohort to stress-test the hardest end of the rare-disease spectrum. Half of the cases (150) have causal genes that lack any ClinVar pathogenic or likely pathogenic variant, forcing the pipeline to rely on phenotype and gene-context reasoning; the ground-truth variants are ClinVar VUS or conflicting interpretations (*UDN-VUS*). The other 150 have catalogued causal genes with prior ClinVar pathogenic annotations, resolved through the same gene-to-variant conversion (*UDN-Pathogenic*).

### LIRDB-47 (*n* = 63)

LIRDB-47 comprises 47 Turkish rare-disease patients from the Intergen Genetic and Rare Diseases Diagnosis and Research Center, of whom 12 carry two or more independently diagnosed causal variants (9 patients with 2 causal variants, 2 with 3, and 1 with 4); each causal variant is evaluated as a separate case, yielding 63 cases. Causal variants were curated from routine clinical diagnostic reports, and HPO annotations were assigned by the treating clinician, making LIRDB-47 the only benchmark whose phenotype labels are unfiltered human-expert output. The cohort uses Intergen’s production exome-sequencing pipeline and the same variant-annotation schema as the simulated benchmarks.

### Gene-to-variant conversion

For both MyGene2 and UDN, each gene-level diagnosis is mapped to a ClinVar variant. Benchmark diagnosis strings and ClinVar disease labels are first canonicalised: lowercased, punctuation stripped, generic suffixes such as “syndrome” or “disease” removed, and Roman numerals normalised to Arabic. For every gene we compute a fuzzy similarity score between the benchmark diagnosis and the disease labels of ClinVar variants in that gene using `rapidfuzz` token-set ratio. The mapping is then resolved by a six-stage cascade designed to balance the two error modes of fuzzy matching: a strict similarity floor of 0.85 together with a pathogenicity-first preference controls Type I error (false matches to the wrong disease entry), and progressive relaxation along the (significance, similarity) axes controls Type II error (excluding cases where no entry survives the strictest criteria). The cascade tries, in order: (i) pathogenic plus disease-label match, (ii) VUS plus disease-label match, (iii) pathogenic without a disease-label constraint, (iv) VUS without a disease-label constraint, (v) benign plus disease-label match, and (vi) benign without a disease-label constraint. The first stage that returns at least one candidate selects one variant uniformly at random, which becomes the case’s ground-truth causal allele. Cases for which no variant passes under any of the six stages are excluded before evaluation. All resulting matches were additionally inspected manually to catch fuzzy-match collisions between closely related but clinically distinct disease subtypes (for example, diabetes type 1 versus type 2), and miscategorised entries were corrected before the cohorts were finalised.

### Epicrisis generation

For every case not in LIRDB-47 we generate a synthetic clinical narrative in three stages. First, a grounded-search language model (Perplexity Sonar, temperature 0.3) is prompted to retrieve common symptoms, rare symptoms, and demographic predilections from indexed literature; cases with a catalogued genedisease label (Custom Simulation, MyGene2, UDN-Pathogenic) use a disease-grounded prompt, and the UDN-VUS half (novel genes lacking a usable disease label) uses a symptom-grounded prompt that takes the HPO term list as input instead. Second, a reasoning model (Gemini 2.5 Flash, temperature 0.3, no extended-reasoning channel) composes a patient vignette under a prompt that instructs it to act as a medical educator preparing a mock clinical examination: include a subset of common symptoms, substitute some rare symptoms for common ones, and insert one or two off-topic distractor symptoms. The vignette is emitted under a structured output schema constraining age, sex, optional ethnicity, and a free-form symptom list. Third, the rendered epicrisis is passed to the HPO extraction pipeline. For LIRDB-47 we retain the clinician-authored epicrisis verbatim.

### HPO extraction via retrieval-augmented generation

For the simulated benchmarks, HPO terms are derived from each case’s epicrisis through a two-call pipeline. A chunking call (Gemini 2.5 Flash) splits the epicrisis into atomic clinical observations, excluding statements about demographics, family history, or negative findings. Each chunk is embedded with a biomedical sentence encoder (BioBERT-MNLI, `pritamdeka/BioBERT-mnli-snli-scinli-scitail-mednli-stsb`) and queried against a ChromaDB collection of pre-embedded HPO term descriptions to retrieve the ten nearest neighbours. A second LLM call ranks the retrieved terms against the chunk and selects the one or two most relevant, producing the case’s final HPO list as the union over chunks. The pipeline is permitted to reject all retrieved terms for a chunk. For MyGene2 and UDN the original HPO lists are used directly, restricted to terms flagged as “observed”. LIRDB-47 uses the clinician-assigned HPO annotations.

### Methodology prompts

The following verbatim prompts drive the epicrisisgeneration and HPO-RAG stages described above. The disease-grounded retrieval and composer prompts are used for cases with a catalogued gene-disease label (Custom Simulation, MyGene2, UDN-Pathogenic); the symptom-grounded variants are used for the UDN-VUS half, where the novel causal gene lacks a usable disease label and the only available signal is the original HPO list. The HPO-RAG pipeline (chunking plus rank-and-select) is shared across all cases.

~~~
**Sonar grounded retrieval (disease-grounded)**
      Find the most relevant information about the following diseases:
      {disease}
      Answer the following three questions for each disease:
      1. What are the general / common symptoms of the disease?
      2. What are the rare / uncommon symptoms of the disease?
      3. Is there a spesific gender or age group that is more likely to get the
      ‘→ disease?
~~~

~~~
**Sonar grounded retrieval (symptom-grounded)**.
      Find the most relevant information about the following hpo terms:
      {hpo_terms}
      Answer the following three questions for each hpo term:
     1. Which of these hpo terms are more common and less informative for a
     ‘→ specific disease?
     2. Which of these hpo terms are rare and informative for a specific
     ‘→ disease?
     3. Is there a spesific gender or age group that is more likely to have
     ‘→ these hpo terms?
~~~

~~~
**Epicrisis composer (disease-grounded).**
     You are a medical expert helping professors to prepare a mock clinical
    ‘→ trial for their students.
    In this sample, there is a list of target diseases. Your job is to prepare
    ‘→ a mock clinical epicrisis for this sample.
    You’re given the following information about the target diseases:
    <disease_info>
    {disease_info}
    </disease_info>
    For this sample, you need to create a fake profile of the patient and their
    ‘→ symptoms!
    Since its an exam, while the symptoms should be as sufficient enough to
    ‘→ make a diagnosis, it shouldn’t be so obvious!
    Additionally, if these diseases are common for some age / gender group or
    ‘→ ethnicity, follow these for your fake profile!
    In your fake profile, you can add short familial history in your symptom ‘→ list especially when the inheritance is    important for the disease’s ‘→ context.
    
    For blurring the symptoms, you can use the following tricks:
    1. Using only a subset of the symptoms
    2. Using rare symptoms instead of common ones
    3. Adding irrelevant symptoms that can be caused by any other insignificant
    ‘→ condition
    Your output should be in the following format:
    {{
       “age”: [age],
       “gender”: [gender],
       “ethnicity”: [ethnicity] (only if relevant else None),
       “symptoms”: [symptoms] (just list some keywords, don’t bother writing
       ‘→ full sentences!)
    }}
~~~

~~~
   **Epicrisis composer (symptom-grounded)**.
   You are a medical expert helping professors to prepare a mock clinical
   ‘→ trial for their students.
   In this sample, there is a list of target hpo terms. Your job is to prepare
   ‘→ a mock clinical epicrisis for this sample.
   
    You’re given the following information about the target hpo terms:
   <hpo_info>
   {hpo_info}
   </hpo_info>
   If these hpo terms are common for some age / gender group or ethnicity,
   ‘→ follow these for your fake profile!
   Your output should be in the following format:
   {{
       “age”: [age],
       “gender”: [gender],
       “ethnicity”: [ethnicity] (only if relevant else None),
       “symptoms”: [symptoms] (just list some keywords, don’t bother writing
       ‘→ full sentences!)
   }}
~~~

~~~
   **HPO-RAG chunking call**.
   Split the following clinical history into separate meaningful parts or
   ‘→ observations.
   Each part should correspond to a single abnormality, sign, symptom that is
   ‘→ directly related to a HPO.
   If you think there is no any direct relation with a HPO, exclude this part
   Exclude any of the following:
   - Statements about demographics, family history, birth history, or parental
   ‘→ information.
   - Statements indicating that tests, evaluations, or findings are normal,
   ‘→ negative, or as expected.
   Do not merge unrelated concepts. If a phrase lists multiple findings (e.g., ‘→ “facial dysmorphic features such as frontal bossing and low-set ears”), split them into separate items.
   Input:
   {clinical_history}
   Output Format (no markdown formatting, no triple backticks, no explanation,
   ‘→ no extra text):
   {{“chunks”: [“clinical history chunk 1”, “clinical history chunk 2”, &]}}
~~~

~~~
**HPO-RAG rank-and-select call**.
      You are a medical expert helping professors to annotate a clinical history
      ‘→ with HPOs. Given a list of HPOs, choose the most relevant ones
      ‘→ (preferably 1 or 2) for the following clinical history:
     Clinical History:
     {clinical_history}
     HPOs:
     {hpo_info}
     Output Format (no markdown formatting, no triple backticks, no explanation,
     ‘→ no extra text):
     {{“hpo_ids”: [“HPO ID 1”, “HPO ID 2”, &]}}
~~~

**Table S2:**
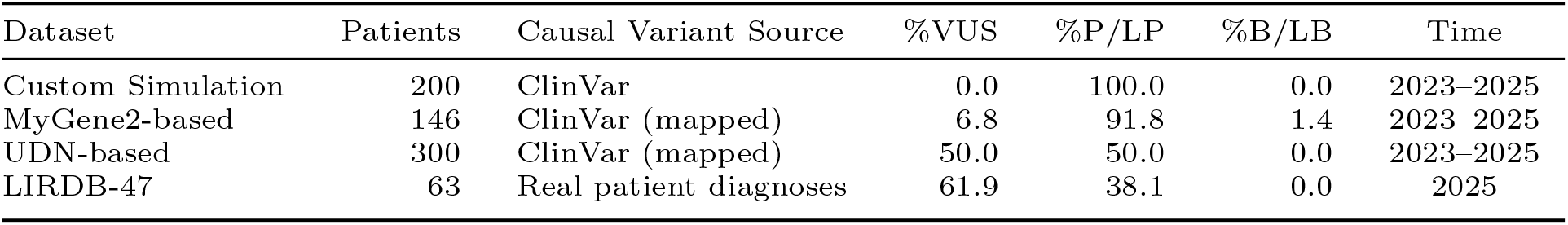
Overview of datasets and benchmarks used to evaluate DAVP. The three simulated cohorts (Custom Simulation, MyGene2-based, UDN-based) draw background genomes from the 1000 Genomes Project; LIRDB-47 uses real Turkish patient exomes. ClinVar shares are computed over the answer variants; for LIRDB-47, the 23 of 63 cases whose causal variant is not deposited in ClinVar are pooled into the VUS share, since they are functionally equivalent for a database-driven prioritiser. P/LP, pathogenic/likely pathogenic; B/LB, benign/likely benign.

**Table S3:**
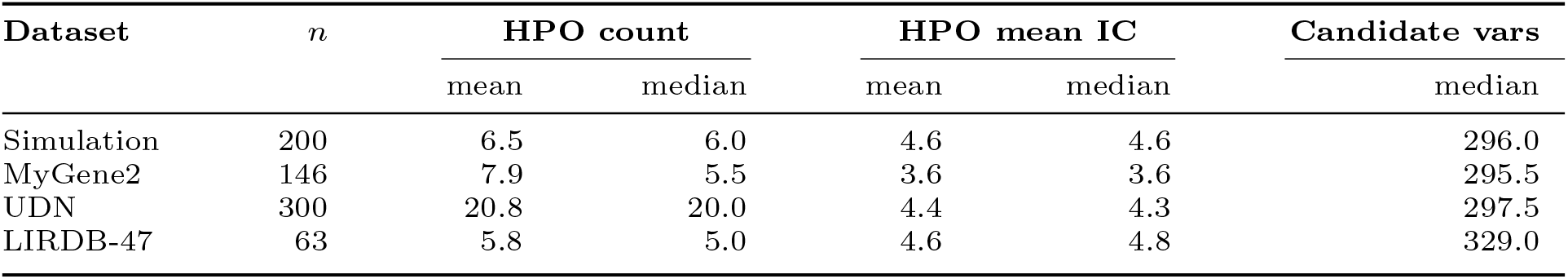
Per-dataset summary of the 709-case benchmark. HPO count is the number of Human Phenotype Ontology terms supplied at input. HPO information content (IC) is the mean over each case’s HPO list, with IC computed as − log(*f*) where *f* is the fraction of OMIM/Orphanet/DECIPHER diseases annotated with the term (or any descendant) in the 2026-02-16 HPO release. Candidate variants are the number of input variants the prioritiser sees per case (the per-sample candidate JSONL row count).

| Dataset | $n$ | HPO count | | HPO mean IC | | Candidate vars |
| --- | --- | --- | --- | --- | --- | --- |
|  |  | mean | median | mean | median | median |
| Simulation | 200 | 6.5 | 6.0 | 4.6 | 4.6 | 296.0 |
| MyGene2 | 146 | 7.9 | 5.5 | 3.6 | 3.6 | 295.5 |
| UDN | 300 | 20.8 | 20.0 | 4.4 | 4.3 | 297.5 |
| LIRDB-47 | 63 | 5.8 | 5.0 | 4.6 | 4.8 | 329.0 |

## S3. Datasets and cohort characterisation

This section provides the cohort-level overview, per-case phenotype/variant-burden statistics, ClinVar assertion distributions, organ-system breakdown, and patient-embedding analysis underlying the four benchmarks evaluated in the main text.

### Cohort overview

Supplementary Table S2 summarises the four benchmark datasets at the cohort level: number of patients, background-genome source, causal-variant source, and ClinVar significance shares.

### Per-case phenotype and variant burden

Supplementary Table S3 reports per-case HPO count, mean HPO information content (IC), and median candidate-variant count for each of the four benchmarks.

The UDN benchmark carries roughly three to four times as many HPO terms per case (median 20) as the other three cohorts (median 5–6), reflecting the detailed phenotyping captured during UDN case reviews. Despite this larger count, UDN’s mean IC per term (4.4 nats) is slightly lower than Simulation’s (4.6), indicating UDN descriptions include a broader mix of highly specific and low-IC constitutional terms. Median candidate variant counts are tightly clustered around 295–330 across cohorts, so none of the prioritisation gaps observed in the main results can be explained by differences in raw input size. The companion violin and UMAP visualisations of these per-case statistics are shown in Supplementary Figure S2 (top row), and the per-case mean HPO information-content distribution is shown in Supplementary Figure S1 below.

**Fig. S1:**
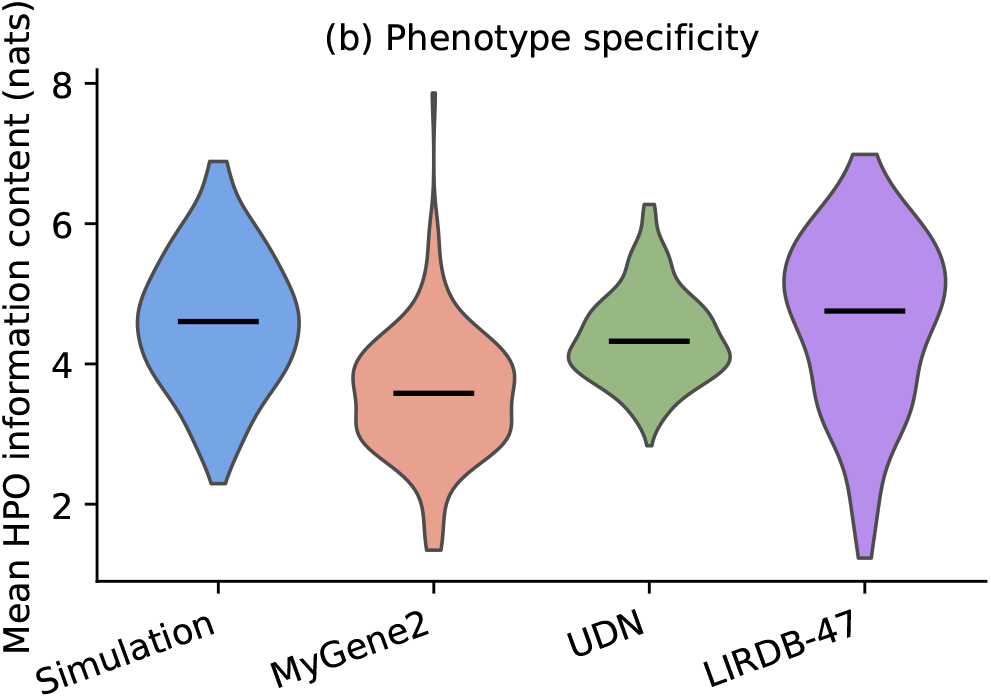
Per-case mean HPO information content (nats) across the four benchmarks. IC is − log(*f*) where *f* is the fraction of OMIM/Orphanet/DECIPHER diseases annotated with the term (or any descendant) in the 2026-02-16 HPO release; the panel reports the per-case mean over each HPO list. UDN carries the broadest IC range; Simulation and LIRDB-47 cluster around the same median.

**Table S4:** ClinVar significance class of the answer variant, tabulated across the four benchmark datasets. The LIRDB-47 column reflects the 40 of 63 cases whose causal variant is deposited in ClinVar; the remaining 23 are clinically validated Turkish variants without a public ClinVar entry and are reported in the *Not in ClinVar* row.

| Clsig | Simulation | MyGene2 | UDN | LIRDB-47 | All |
| --- | --- | --- | --- | --- | --- |
| Pathogenic | 83 | 67 | 88 | 23 | 261 |
| Likely Pathogenic | 117 | 67 | 62 | 1 | 247 |
| VUS | 0 | 10 | 111 | 5 | 126 |
| Conflicting | 0 | 0 | 39 | 11 | 50 |
| Likely Benign | 0 | 2 | 0 | 0 | 2 |
| Not in ClinVar | 0 | 0 | 0 | 23 | 23 |
| Any | 200 | 146 | 300 | 63 | 709 |

### ClinVar assertion distribution

Supplementary Tables S4 and S5 break down the ClinVar significance class and review status of each cohort’s answer variants. The UDN cohort is by construction split between 150 novel-gene VUS cases and 150 catalogued-gene pathogenic cases. MyGene2 entries are resolved through the ClinVar fuzzy-match procedure and are concentrated in the pathogenic and likely pathogenic classes.

### Disease-category breakdown

Supplementary Table S6 cross-tabulates the answer gene’s disease category (primary organ system) against dataset; Supplementary Table S7 reports the corresponding pooled top-3 and top-20 diagnosis recall per category. Brain and Nerves dominates every cohort (314/646 cases), reflecting the overrepresentation of neurological rare diseases in publicly available benchmarks; LIRDB-47 additionally contributes substantial musculoskeletal and endocrine cases. Recall is uniformly above 63% top-3 and 75% top-20 across all 14 categories.

**Fig. S2:**
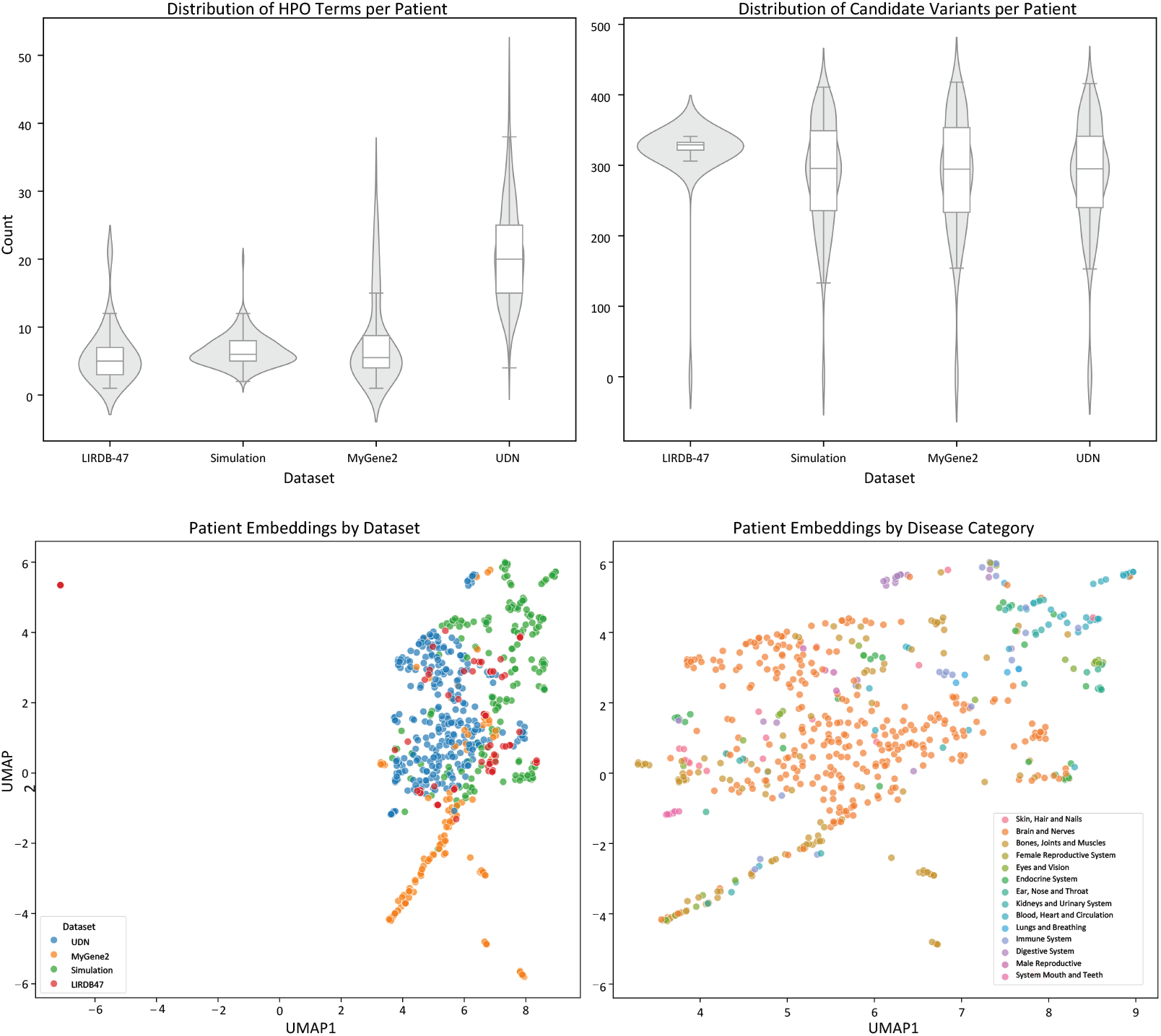
Dataset characteristics and patient embedding analysis. **Top row:** Distribution of HPO terms per patient (left) and candidate variants per patient (right) across datasets, shown as violin plots with embedded box plots. **Bottom row:** UMAP visualisation of patient embeddings derived from the clinical narratives, coloured by dataset (left) and by disease category (right). LIRDB-47 occupies the same region of embedding space as the three simulated cohorts, indicating that the synthetic clinical narratives generated by our epicrisis-RAG pipeline are not far-fetched from real-world clinician-authored descriptions; the disease-category panel further shows that cases with similar organ-system labels cluster together regardless of source dataset.

### Patient-embedding overlap with the real-world cohort

A natural concern with simulated benchmarks is whether the synthetic cases drift far from the distribution of real clinical narratives. To check this we embed each case’s epicrisis with a sentence encoder and project the resulting vectors into two dimensions with UMAP (Supplementary Figure S2, bottom row). LIRDB-47 occupies the same region of patient-embedding space as the three simulated cohorts, indicating that the synthetic clinical narratives generated by our epicrisis-RAG pipeline are not far-fetched from real-world clinician-authored descriptions; prioritisation performance on the simulated benchmarks is therefore informative about expected real-world behaviour. When coloured by disease category, cases with similar organ-system labels cluster together regardless of source dataset, reinforcing that the embedding captures clinically meaningful structure rather than dataset-of-origin artefacts.

**Table S5:** ClinVar review status (stars) of the answer variant across the four benchmark datasets. Higher stars indicate stronger curator assertion. The LIRDB-47 column reflects the 40 of 63 cases whose causal variant is deposited in ClinVar.

| Clnrevstat | Simulation | MyGene2 | UDN | LIRDB-47 | All |
| --- | --- | --- | --- | --- | --- |
| 0 stars | 0 | 0 | 37 | 11 | 48 |
| 1 star | 10 | 33 | 67 | 11 | 121 |
| 2 stars | 90 | 73 | 143 | 14 | 320 |
| 3 stars | 83 | 39 | 52 | 4 | 178 |
| 4 stars | 17 | 1 | 1 | 0 | 19 |
| Not in ClinVar | 0 | 0 | 0 | 23 | 23 |
| Any | 200 | 146 | 300 | 63 | 709 |

**Fig. S3:**
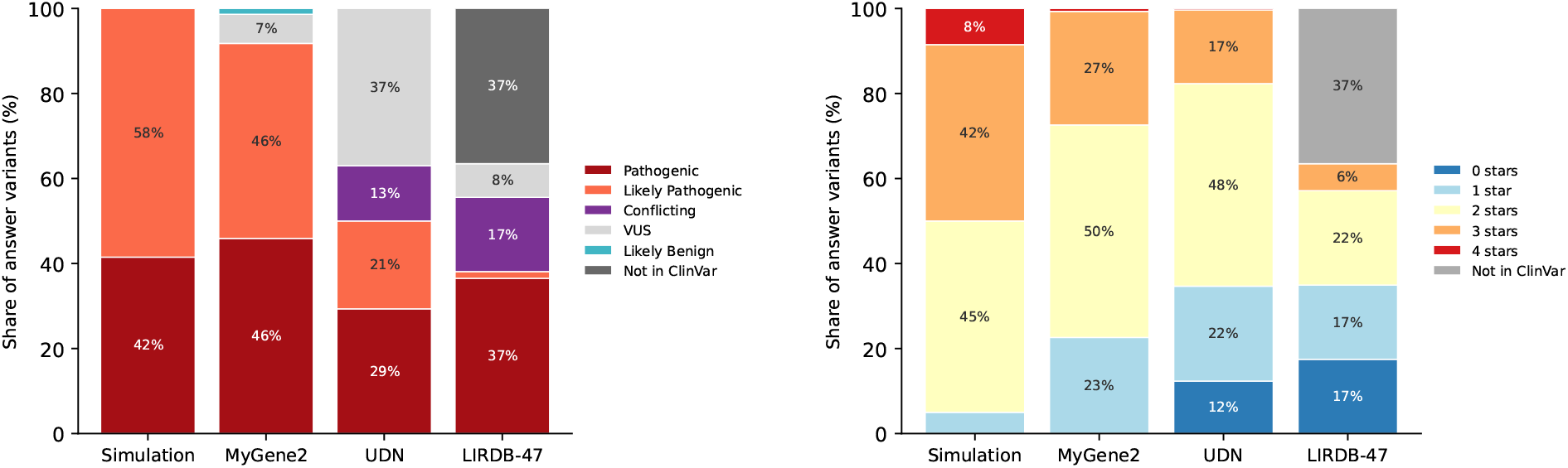
Distribution of ClinVar assertions for the answer variants in each benchmark. (a) Clinical significance class. (b) Review status (0–4 stars). LIRDB-47’s 23 of 63 cases that are not deposited in ClinVar are pooled into the dedicated *Not in ClinVar* series in both panels, so the LIRDB-47 column sums to 100% over all 63 cases.

**Table S6:**
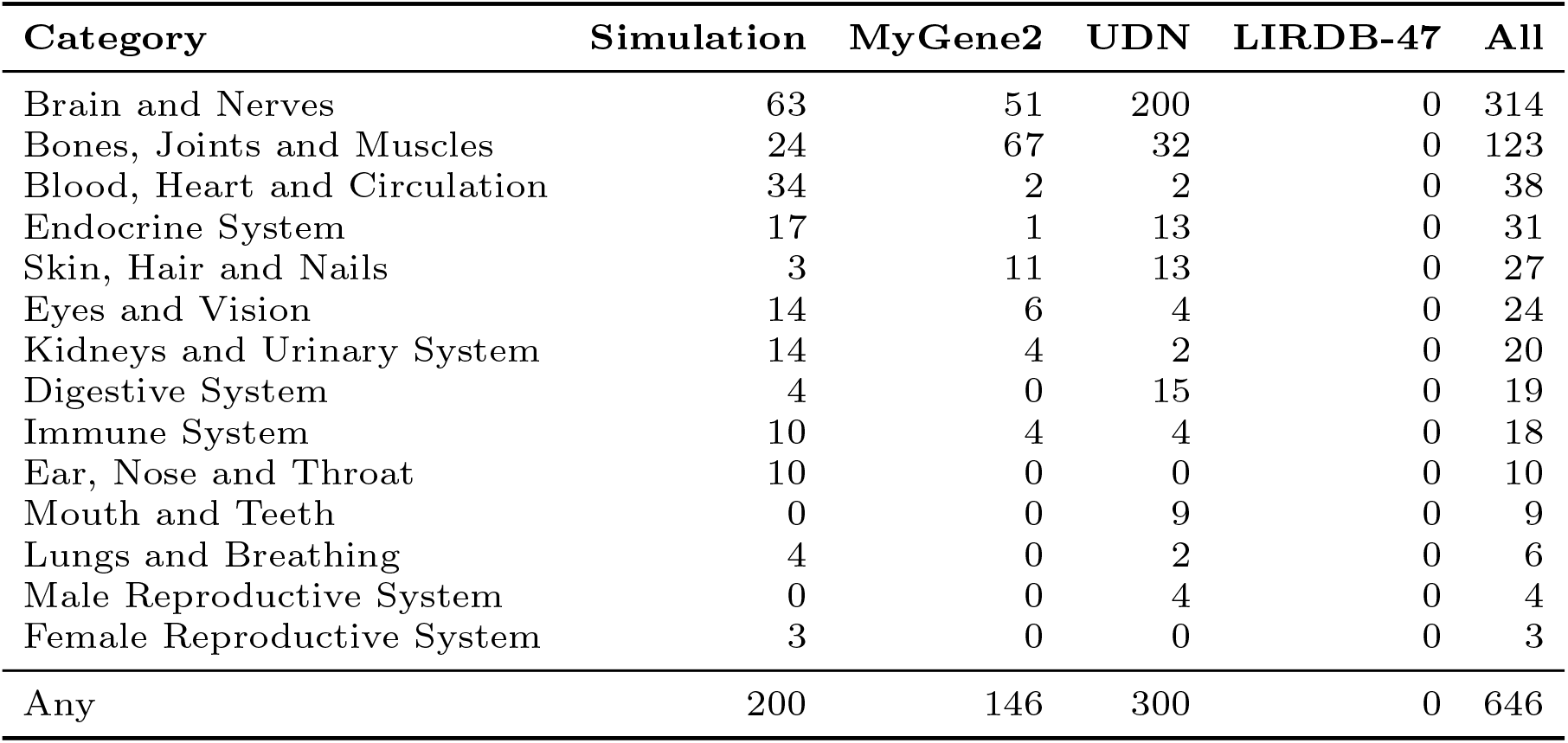
Disease category (primary organ system) of the answer gene across the four benchmark datasets. Categories are sorted by overall cohort size.

**Table S7:** Breakdown of pooled diagnostic recall (Top-3 and Top-20) by disease category (organ system).

| Category | N | Recall@3 (%) | Recall@20 (%) |
| --- | --- | --- | --- |
| Brain and Nerves | 314 | 85.99 | 92.68 |
| Bones, Joints and Muscles | 123 | 87.80 | 95.93 |
| Blood, Heart and Circulation | 38 | 89.47 | 94.74 |
| Endocrine System | 31 | 70.97 | 77.42 |
| Skin, Hair and Nails | 27 | 96.30 | 100.00 |
| Eyes and Vision | 24 | 95.83 | 95.83 |
| Kidneys and Urinary System | 20 | 100.00 | 100.00 |
| Digestive System | 19 | 63.16 | 84.21 |
| Immune System | 18 | 88.89 | 100.00 |
| Ear, Nose and Throat | 10 | 100.00 | 100.00 |
| Mouth and Teeth | 9 | 66.67 | 77.78 |
| Lungs and Breathing | 6 | 83.33 | 100.00 |
| Male Reproductive System | 4 | 75.00 | 75.00 |
| Female Reproductive System | 3 | 100.00 | 100.00 |

**Table S8:** Schema of a DAVP gene cache dossier. The presence column is the fraction of the 16,311 unique cached genes whose dossier explicitly contains that heading; sections with no source evidence are emitted with a “not available” placeholder, which we count as present.

| Section | Content | Presence (%) |
| --- | --- | --- |
| Gene Identity & Clinical Context | HGNC/OMIM identifiers, primary disease associations, clinical significance, inheritance patterns | 99.7 |
| Constraint & Variant Intolerance | gnomAD pLI, LOEUF, pRec, pNull; interpretation of constraint scores | 99.6 |
| Phenotype Spectrum & HPO Terms | Primary and secondary HPO terms, age-of-onset patterns, severity spectrum | 99.5 |
| Genotype-Phenotype Correlations | Variant class to phenotype mappings, protein-domain-specific patterns, representative examples | 99.5 |
| Clinical Variants & Phenotype Associations | Well-characterised pathogenic variants from ClinVar with established phenotype associations | 98.7 |
| Tissue Expression & Clinical Relevance | GTEx-derived high-expression tissues, tissue-specific phenotypes, developmental expression | 99.5 |
| Molecular Mechanism & Phenotype Pathways | Normal function, disease mechanism, disrupted cellular pathways, protein-protein interactions | 99.5 |
| Diagnostic Yield & Clinical Utility | Contribution to overall yield, clinical utility of testing, reclassification trends | 95.1 |
| Key Clinical Literature & Studies | Landmark studies, cohort sizes, diagnostic recommendations | 99.5 |
| HPO-Variant Matching Summary | Per-run summary of how documented phenotypes align with the patient’s HPO terms | 99.4 |

## S4. Gene cache

The gene cache is a shared, patient-agnostic resource consulted by every DAVP stage that reasons over genes. Each entry is a free-text biomedical dossier summarising clinical context, constraint metrics, phenotype spectrum, mechanism of disease, and representative pathogenic variants for a single gene. Dossiers are generated once by a grounded-search language model and reused across every patient whose candidate list intersects that gene, so the per-gene literature retrieval cost is amortised over the full benchmark.

### Schema

Every dossier follows the fixed ten-section outline in Supplementary Table S8. The first nine sections are produced from grounded web retrieval and are patient-agnostic; the tenth (*HPO-Variant Matching Summary*) is a short per-patient addendum generated at runtime and is not part of the cached resource. Sections with no retrievable evidence are filled with an explicit “not available” placeholder rather than omitted.

### Coverage and statistics

Aggregating the unique per-gene dossiers consulted across the 709-case benchmark, the cache spans 16,311 unique genes with a mean dossier length of 9,206 characters and 94.7% of dossiers containing all ten template sections (Supplementary Table S9). The dossier length distribution is approximately unimodal and centred on 9,000 characters; the long tail corresponds to genes with extensive clinical literature coverage (e.g., *COL1A1, NF1, CFTR*), while the short tail comprises uncharacterised genes with placeholders in most sections.

**Table S9:** Aggregate statistics of the DAVP gene cache, computed across the 709-case benchmark.

| **Metric** | **Value** |
| --- | --- |
| Unique genes cached | 16,311 |
| Patient runs contributing dossiers | 692 |
| Mean dossiers per sample | 227.4 |
| Median dossiers per sample | 256 |
| Mean dossier length (chars) | 9,206 |
| Median dossier length (chars) | 9,011 |
| P10 / P90 dossier length (chars) | 6,555 / 12,155 |
| Mean sections present per dossier | 9.9 / 10 |
| Fraction of dossiers with all 10 sections | 94.7% |

### Generation prompt

Each dossier is produced by a single grounded-search LLM call instantiated with the gene symbol and any pre-existing structured fields, which are passed in as {`gene_symbol}` and {`existing_data_summary}`. The prompt enforces the ten-section schema, mandates ten parallel web search queries (HGNC/OMIM, ClinVar, HPO, GTEx, gnomAD, PubMed, etc.) so that every fact is grounded against external sources rather than the model’s training corpus, and requires inline numbered citations resolved in a final **References** section. The dossier emitted by this call is stored verbatim and consumed as-is by the downstream Prelimin8, Elimin8, and inGeneTopMatch stages.

~~~
[SYSTEM]
You are a clinical geneticist and biomedical knowledge curator specializing
‘→ in genotype-phenotype correlations.
Your task is to create a comprehensive gene dossier that will be used for
‘→ matching patient phenotypes (HPO terms) with genetic variants to
‘→ determine
pathogenicity and disease causality.
Formatting & style rules
- Write in clear clinical language with focus on phenotype-genotype
‘→ relationships.
- Use **bold Markdown section headers** followed by concise bullet points
‘→ (<= 3 lines per bullet).
- Add an inline numbered citation after each fact - e.g. [1], [2].
- Prioritize information that helps correlate variants with specific
‘→ phenotypes.
- End with a “** References **” header and a numbered list of URLs/DOIs in
‘→ citation order.
- For URLs: Use original URLs when available, otherwise redirect URLs are
‘→ acceptable.
- Do not include explanatory text about your own reasoning - only the
‘→ clinical dossier.
[USER]
Create a comprehensive clinical genetics dossier for the human gene
‘→ “{gene_symbol}”.
**PRIMARY PURPOSE: This dossier will be used to match patient HPO terms ‘→ with genetic variants to determine which variants are most likely pathogenic and causative for the patient’s phenotype.**
{existing_data_summary}
**CRITICAL REQUIREMENT: YOU MUST USE WEB SEARCH FOR ALL INFORMATION. Do not
‘→ use any training data. Search for each piece of information
‘→ separately.**
**REQUIRED SEARCH QUERIES: You must perform exactly these 10 web
‘→ searches:**
1. “{gene_symbol} gene HGNC OMIM disease phenotype”
2. “{gene_symbol} ClinVar pathogenic variants phenotype HPO”
3. “{gene_symbol} HPO terms Human Phenotype Ontology”
4. “{gene_symbol} genotype phenotype correlation studies”
5. “{gene_symbol} clinical cases phenotype HPO codes”
6. “{gene_symbol} GTEx tissue expression clinical relevance”
7. “{gene_symbol} protein function variant effects phenotype”
8. “{gene_symbol} gnomAD constraint pLI LOEUF intolerance”
9. “{gene_symbol} disease mechanism phenotype pathway”
10. “{gene_symbol} publications phenotype clinical genetics”
**Each search query must be used to gather specific information. Prioritize ‘→ recent data (2020-2024) when available, but include landmark historical studies that established key genotype-phenotype correlations.**
Follow this outline exactly; omit a subsection only if no data exist after
‘→ web search.
**Gene Identity & Clinical Context**
- HGNC ID, OMIM gene ID, primary disease associations
- Clinical significance level (definitive, strong, moderate evidence)
- Inheritance patterns observed in patients
**Constraint & Variant Intolerance**
- pLI, LOEUF, pRec, pNull (report exact numeric values)
- Clinical interpretation of constraint scores
- Variant classes most likely to be pathogenic (nonsense, missense, splice,
‘→ etc.)
**Phenotype Spectrum & HPO Terms**
- **Primary HPO terms** (<= 20, ranked by frequency in literature)
- **Secondary HPO terms** (<= 15, less common but documented)
- Age of onset patterns (prenatal, infantile, childhood, adult)
- Phenotype severity spectrum (mild, moderate, severe)
**Genotype-Phenotype Correlations**
- Variant classes and their typical phenotypes
- Protein domain-specific phenotype patterns
- Genotype-phenotype correlation strength (strong, moderate, weak)
- Examples: specific variants -> specific phenotypes
**Clinical Variants & Phenotype Associations**
- Up to 15 well-characterized pathogenic variants
- Format: rsID / HGVS / ClinVar significance / Reported phenotypes (HPO
‘→ terms) / AF
- Variants with strongest phenotype evidence
- Novel variants from recent case reports
**Tissue Expression & Clinical Relevance**
- Highest expressing tissues (GTEx TPM) and clinical correlation
- Tissue-specific phenotypes expected
- Expression during development and age-related phenotypes
**Molecular Mechanism & Phenotype Pathways**
- Normal gene function in one sentence
- Disease mechanism: haploinsufficiency, dominant-negative,
‘→ gain-of-function
- Cellular/molecular pathways disrupted -> phenotype consequences
- Protein-protein interactions relevant to phenotype
**Diagnostic Yield & Clinical Utility**
- Diagnostic yield in clinical cohorts
- Most common reasons for testing this gene
- Clinical actionability and management implications
- Genetic counseling considerations
**Key Clinical Literature & Studies**
- PMID, year, key phenotype findings (up to 10 papers)
- Include both recent studies (2020-2025) and landmark historical papers
‘→ that established key genotype-phenotype correlations
- New genotype-phenotype correlations discovered
- Clinical trial outcomes if available
**HPO-Variant Matching Summary**
- **High-confidence HPO-variant associations** (which variants typically
‘→ cause which phenotypes)
- **Phenotype red flags** (HPO terms that strongly suggest pathogenic
‘→ variants in this gene)
- **Differential diagnosis considerations** (overlapping phenotypes with
‘→ other genes)
** References **
(numbered list of URLs/DOIs - use original URLs when available, redirect
‘→ URLs acceptable if originals not accessible)
Priority data sources: HGNC, OMIM, ClinVar, HPO, gnomAD, GTEx, PubMed,
‘→ ClinGen, Orphanet, DECIPHER.
~~~

### Example dossier

To illustrate the content and density of a cached entry, we reproduce the full dossier for *HNF1A*, the causal gene in several LIRDB-47 and simulated MODY cases. The dossier is shown verbatim and untruncated.

~~~
HNF1A Gene Biomedical Dossier
### **Clinical Genetics Dossier: HNF1A**
**Gene Identity & Clinical Context**
*  **HGNC ID:** 11621.
*  **OMIM Gene ID:** 142410.
*  **Primary Disease Associations:** Maturity-Onset Diabetes of the Young,
‘  → Type 3 (MODY3), Familial Hepatic Adenomas, and an increased risk for
‘  → Type 2 Diabetes.
*  **Clinical Significance Level:** The association between HNF1A and MODY3
‘  → is considered **Definitive**.
*  **Inheritance Patterns:** Primarily Autosomal Dominant for MODY3.
*  *Constraint & Variant Intolerance**
*  **pLI, LOEUF, pRec, pNull:** The gnomAD v2.1.1 pLI score (probability of
‘  → being loss-of-function intolerant) is 0.00, suggesting tolerance to
‘  → heterozygous loss-of-function (LoF) variants in the general population.
‘  → The loss-of-function observed/expected upper bound fraction (LOEUF)
‘  → score is a more continuous metric; a low LOEUF indicates intolerance to
‘  → LoF variation.
*  **Clinical Interpretation of Constraint Scores:** The low pLI score is ‘→ not unexpected for a gene associated with a dominant but later-onset ‘→ disease. This score primarily identifies genes where heterozygous LoF
‘  → variation leads to severe pediatric disease, not adult-onset conditions
‘  → like MODY.
*  **Variant Classes Most Likely to be Pathogenic:** Missense, frameshift,
‘  → nonsense, and splice site mutations are the most common types of
‘  → pathogenic variants reported.
**Phenotype Spectrum & HPO Terms**
*  **Primary HPO terms:**
   * HP:0000478 - Abnormality of the eye
   * HP:0000593 - Abnormality of the kidney
   * HP:0001518 - Small for gestational age
   * HP:0003077 - Hyperglycemia
   * HP:0002910 - Low renal threshold for glucose reabsorption
   * HP:0002645 - Hepatic adenoma
   * HP:0000819 - Diabetes mellitus
   * HP:0000833 - Gestational diabetes mellitus
   * HP:0004322 - Polyuria
   * HP:0001945 - Polydipsia
   * HP:0002901 - Fatigue
   * HP:0000545 - Myopia
   * HP:0000639 - Nystagmus
   * HP:0001639 - Hypertrophic cardiomyopathy
   * HP:0004908 - Proteinuria
   * HP:0000093 - Renal insufficiency
   *HP:0001988 - Recurrent skin infections
*  **Secondary HPO terms:**
   * HP:0002120 - Seizures
   * HP:0001250 - Seizures
   * HP:0002013 - Vomiting
   * HP:0001508 - Failure to thrive
   * HP:0003236 - Elevated circulating C-reactive protein concentration
   * HP:0001947 - Hypoglycemia
   * HP:0030680 - Hyperinsulinism
*  **Age of Onset Patterns:** Onset of diabetes is typically before 30-35
‘→  years of age, though a significant proportion of carriers develop
‘→  diabetes after young adulthood. Neonatal hyperinsulinism has also been
‘→  reported.
*  **Phenotype Severity Spectrum:** The phenotype can range from mild,
‘→  non-progressive hyperglycemia manageable by diet to progressive
‘→  hyperglycemia requiring pharmacological intervention.
**Genotype-Phenotype Correlations**
*           **Variant Classes and Their Typical Phenotypes:** Protein-truncating
‘→   variants (PTVs), such as nonsense and frameshift mutations, are
‘→   generally associated with an earlier age of diabetes diagnosis compared
‘→   to missense variants.
* **Protein Domain-Specific Phenotype Patterns:** Mutations in the 
‘→   DNA-binding domain are common and disrupt the protein’s ability to 
‘→   regulate target genes. Dimerization domain mutations prevent the 
‘→   formation of functional transcription factor units.
*  **Genotype-Phenotype Correlation Strength:** The correlation is
‘→   considsered moderate; while truncating mutations often lead to an earlier
‘→   onset, significant variability exists. For instance, the common I27L 
‘→   polymorphism can modify the age of diagnosis in individuals with a 
‘→   pathogenic ‘HNF1A’ mutation.
*  **Examples: specific variants → specific phenotypes:**
          * p.(Gly292fs): This common frameshift variant is associated with
          ‘→ classic HNF1A-MODY, characterized by hyperglycemia, insulin
          ‘→ deficiency, and markedly lower adult BMI.
          * p.Arg271Glyfs: A frameshift variant identified as a likely de novo
          ‘→ mutation, leading to MODY3.
**Clinical Variants & Phenotype Associations**
*  Up to 15 well-characterized pathogenic variants:
          * c.872dup / p.Leu291fsinsC: Pathogenic / Maturity-onset diabetes of
          ‘→ the young, type 3 (MODY3)
          * c.608G>A / p.Arg203His: Pathogenic / Monogenic diabetes
          * c.811del / p.Arg271Glyfs: Pathogenic / MODY3
          * p.G319S: Associated with a decrease in insulin secretion before
          ‘→ diabetes onset in T2D patients.
          * c.391C>T: Pathogenic / MODY3
          * p.P291fsinsC: A dominant-negative mutant associated with reduced
          ‘→ insulin secretion.
          * c.-158insGGGTTGG: Pathogenic / MODY3 (promoter mutation)
          * p.G31X: Pathogenic / MODY3
          * p.E41X: Pathogenic / MODY3
          * p.Q130X: Pathogenic / MODY3
          * p.L162P: Pathogenic / MODY3
          * p.R245I: Pathogenic / MODY3
          * p.A269P: Pathogenic / MODY3
          * p.S355X: Pathogenic / MODY3
          * p.Q398X: Pathogenic / MODY3
**Tissue Expression & Clinical Relevance**
*         **Highest Expressing Tissues (GTEx TPM):** Liver, Kidney (Cortex), and
‘→  Pancreas.
*         **Tissue-Specific Phenotypes Expected:**
                 * **Pancreas:** Impaired beta-cell function leads to defective insulin
                 ‘→ secretion and diabetes (MODY3).
                 * **Liver:** Dysregulation of gene expression can lead to hepatic
                 ‘→ adenomas and an atherogenic lipid profile.
                 * **Kidney:** Altered glucose reabsorption leads to a low renal
                 ‘→ threshold for glucosuria.
* **Expression During Development:** HNF1A is essential for the mature
‘→ pancreatic beta-cell in the adult pancreas.
**Molecular Mechanism & Phenotype Pathways**
*         **Normal Gene Function:** HNF1A is a transcription factor that forms 
‘→ homodimers to regulate the expression of genes crucial for pancreatic 
‘→ islet cell and liver function, including those involved in glucose
‘→ metabolism and transport.
* **Disease Mechanism:**
*             **Haploinsufficiency** is the primary mechanism for MODY3, where one 
‘→     non-functional copy of the gene is insufficient for normal beta-cell 
‘→     function.
*             **Dominant-negative** effects have also been described for certain
‘→     mutations.
*             Rare **gain-of-function (GOF)** variants have been identified that 
‘→     protect from type 2 diabetes but increase the hepatic secretion of 
‘→     atherogenic lipoproteins.
*             **Cellular/Molecular Pathways Disrupted → Phenotype Consequences:**
                     * Disruption of the HNF1A/HNF4A regulatory network impairs
                     ‘→ glucose-stimulated insulin secretion from pancreatic beta cells,
                     ‘→ causing hyperglycemia.
                     * In the liver, HNF1A dysfunction alters the expression of genes like
                     ‘→ ‘ANGPTL3’ and ‘PCSK9’, promoting a pro-atherogenic serum profile.
*             **Protein-Protein Interactions Relevant to Phenotype:** HNF1A interacts
‘→     with HNF4A in a cross-regulatory network essential for maintaining
‘→     glucose homeostasis.
**Diagnostic Yield & Clinical Utility**
*          **Diagnostic Yield:** Pathogenic variants in HNF1A account for a
‘→  significant portion of MODY cases, often cited as the most common cause
‘→  (MODY3).
*          **Most Common Reasons for Testing:** Clinical suspicion of MODY,
‘→  characterized by early-onset diabetes (typically <30 years), autosomal
‘→  dominant family history, and absence of pancreatic autoantibodies.
*          **Clinical Actionability and Management Implications:** Patients with 
‘→  HNF1A-MODY are often highly sensitive to low-dose sulfonylureas, which 
‘→  can be a more effective treatment than insulin, especially in the
‘→  initial stages.
*          **Genetic Counseling Considerations:** Autosomal dominant inheritance 
‘→  means a 50% risk of transmission to offspring. Predictive testing for 
‘→  at-risk family members is possible. Penetrance is high, but the age of 
‘→  onset can be variable.
**Key Clinical Literature & Studies**
                  * **PMID: 35171780 (2022):** This review highlights that HNF1A mutations
‘→  cause MODY3 by altering glucose metabolism in beta cells, leading to
‘→  reduced insulin secretion.
*          **PMID: 35075678 (2022):** A multigenerational study of the common
‘→  p.(Gly292fs) variant showed it causes not only hyperglycemia and insulin 
‘→  deficiency but also increased lipolysis and lower BMI, with a significa _⌋_ 
‘→  nt portion of carriers developing diabetes after young adulthood.
*          **PMID: 37248270 (2023):** A landmark study demonstrating that rare
‘→  gain-of-function HNF1A variants can protect against type 2 diabetes but 
‘→  paradoxically increase the secretion of atherogenic lipoproteins from 
‘→  the liver.
*   **PMID: 29895583 (2018):** Established that the common HNF1A variant
‘→  I27L acts as a genetic modifier, influencing the age of diabetes
‘→  diagnosis in individuals with protein-truncating HNF1A mutations.
*   **PMID: 21220268 (2011):** A study from a German-Austrian database that 
‘→  characterized the clinical phenotypes of HNF1A-MODY patients and found 
‘→  that individuals with missense mutations were less likely to require
‘→  insulin than those with truncating mutations.
*   **PMID: 33318991 (2020):** Reported on the incidence of HNF1A variants
‘→  in a South African population, highlighting the presence of
‘→  MODY-associated variants.
*   **PMID: 36622176 (2023):** A case report of a de novo frameshift
‘→  mutation (p.Arg271Glyfs) causing MODY3 in an Indian patient, emphasizing
‘→  the role of spontaneous mutations.
*   **PMID: 19349603 (2009):** Showed that common HNF1A polymorphisms are
‘→  associated with multiple cardiovascular risk phenotypes, including
‘→  levels of C-reactive protein and gamma-glutamyl transferase.
*   **PMID: 32363579 (2020):** This review summarizes the role of HNF1A 
‘→  polymorphisms in MODY3, T2D, and gestational diabetes, noting the 
‘→  considerable phenotypic variability.
*   **PMID: 37471891 (2024):** A study exploring the molecular mechanism of 
‘→  how variants in the HNF4A promoter, a gene regulated by HNF1A, can lead 
‘→  to a MODY phenotype, illustrating the intricate regulatory network.
**HPO-Variant Matching Summary**
*           **High-confidence HPO-variant associations:**
                   * Truncating variants (nonsense, frameshift) are strongly associated
                   ‘→ with a classic MODY3 phenotype: **Hyperglycemia (HP:0003077)**, 
                   ‘→ **Maturity-onset diabetes of the young (HP:0004904)**, and **Low 
                   ‘→ renal threshold for glucose reabsorption (HP:0002910)**.
* **Phenotype red flags:**
                   * The combination of early-onset diabetes (before age 35), a
                   ‘→ multi-generational family history of diabetes, and significant
                   ‘→ glucosuria at normal or only mildly elevated blood glucose levels
                   ‘→ strongly suggests a pathogenic ‘HNF1A’ variant.
                   * Presence of **Hepatic adenoma (HP:0002645)** in a patient with
                   ‘→ diabetes is a significant indicator.
* **Differential diagnosis considerations:**
                   * **HNF4A-MODY (MODY1):** Phenotypically very similar to HNF1A-MODY,
                   ‘→ including sensitivity to sulfonylureas.
                   * **GCK-MODY (MODY2):** Typically causes a stable, mild fasting
                   ‘→ hyperglycemia and rarely requires treatment, unlike the progressive
                   ‘→ hyperglycemia in HNF1A-MODY.
                   * **HNF1B-MODY (MODY5):** Often presents with renal abnormalities 
                   ‘→ (cysts, renal dysplasia) and pancreatic atrophy, which are less 
                   ‘→ common in HNF1A-MODY.
                   * **Type 1 Diabetes:** Distinguished by the presence of pancreatic
                   ‘→ autoantibodies.
                   * **Type 2 Diabetes:** HNF1A-MODY patients are typically younger at
                   ‘→ diagnosis, not obese, and show marked sensitivity to sulfonylureas.
~~~

**Table S10:** DAVP hyperparameter configuration used for all experiments. All stages use Gemini 2.5 Flash as the base model with temperature 0.7 and maximum token limit of 65,536.

| Stage | Model | Temp. | Thinking Budget | Score Weights |
| --- | --- | --- | --- | --- |
| Prelimin8 | Gemini 2.5 Flash | 0.7 | 8,192 | [10, 7, 5, 3, 2, 1, 0, -1] |
| inGeneTopMatch | Gemini 2.5 Flash | 0.7 | 0 | — |
| Elimin8 | Gemini 2.5 Flash | 0.7 | 8,192 | [10, 7, 5, 3, 2, 1, 0, -1] |
| Final Tournament | Gemini 2.5 Flash | 0.7 | 8,192 | — |

(B) Pipeline Flow
| Stage | Input | Output |
| --- | --- | --- |
| Exomiser | All variants from VCF | Top-256 genes |
| Prelimin8 | Top-256 genes | Top-64 genes |
| inGeneTopMatch | Top-64 genes | Detailed variant reports |
| Elimin8 | Variant reports | Top-20 variants |
| Final Tournament | Top-20 variants | Top-3 variants |

**Table S11:** Per-stage DAVP configuration and measured mean LLM call count over the 709-case benchmark. Companion to Table S10; reports the same per-stage configuration in a single horizontal row alongside the measured mean call count, for cross-reference with the per-cohort breakdown in Table S12.

| Stage | Model | Temp. | Max out. | Thinking | Call type | Mean calls |
| --- | --- | --- | --- | --- | --- | --- |
| Prelimin8 | Gemini 2.5 Flash | 0.7 | 65536 | 8192 | 8-way rank | 42.9 |
| inGeneTopMatch | Gemini 2.5 Flash | 0.7 | 65536 | 0 | 1 report | 81.3 |
| Elimin8 | Gemini 2.5 Flash | 0.7 | 65536 | 8192 | 8-way rank | 33.6 |
| Final Tournament | Gemini 2.5 Flash | 0.7 | 65536 | 8192 | 1 winner | 27.5 |
| **Total per sample** |  |  |  |  |  | **181.1** |

## S5. DAVP stage configurations and pipeline prompts

DAVP orchestrates four LLM-driven stages (Prelimin8, inGeneTopMatch, Elimin8, Final Tournament) against a fixed base model with stage-specific prompting and output structure. This section reports the configuration used for every benchmark run, the measured per-stage call cost over the full 709-case benchmark, and the verbatim prompt templates.

### Shared configuration

All four stages use Gemini 2.5 Flash with sampling temperature 0.7, a 65,536-token output ceiling, and an 8,192-token extended-reasoning budget per call (inGeneTopMatch is the sole exception, running with the thinking budget disabled because its task is summarisation rather than ranking). Stages differ only in the prompt template, the structured response schema, and the algorithmic role of each call (Supplementary Tables S10 and S11). No retrieval is performed at inference time: every stage operates over a fixed context assembled from the patient record, the shared gene cache, and the Detailed Variant Reports produced by inGeneTopMatch.

### Measured cost

The theoretical LLM call count per stage follows the formulas in Supplementary Table S1. We measure the realised cost per case over the full benchmark and report per-cohort means in Supplementary Table S12. Prelimin8 and the Final Tournament have nearly constant cost across datasets because their input sizes are fixed by the preceding stages; inGeneTopMatch and Elimin8 vary with the number of candidate variants in the top-gene set, and consequently are largest on the UDN cohort with its broader candidate lists.

**Table S12:** Measured LLM call counts per sample, per stage, under the paper pipeline configuration. Each value is mean ± s.d. over the cases in the row’s cohort.

| Cohort | N | Prelimin8 | inGeneTopMatch | Elimin8 | Final Tourn. | Total |
| --- | --- | --- | --- | --- | --- | --- |
| Simulation | 200 | 41.9 ± 8.9 | 81.2 ± 8.0 | 17.8 ± 3.5 | 27.6 ± 1.6 | 165.7 ± 23.2 |
| MyGene2 | 146 | 42.2 ± 8.8 | 81.4 ± 8.9 | 20.9 ± 13.6 | 27.5 ± 1.5 | 167.6 ± 30.1 |
| UDN | 300 | 42.9 ± 7.7 | 82.5 ± 10.3 | 44.0 ± 27.5 | 27.4 ± 1.7 | 191.1 ± 44.4 |
| LIRDB-47 | 63 | 48.0 ± 0.2 | 75.4 ± 4.3 | 63.9 ± 3.4 | 27.5 ± 1.6 | 210.9 ± 24.9 |
| All | 709 | 42.9 ± 8.1 | 81.3 ± 9.2 | 33.6 ± 24.3 | 27.5 ± 1.6 | 180.9 ± 38.1 |

### Prelimin8 prompt

Eight candidate genes are presented as their cached dossiers augmented with up to five of the patient’s variants in each gene; the model emits a total ranking constrained to a JSON object with keys `rank_1`.. `rank_8`.

~~~
You are a genetic assistant AI. You will be given genes and the patient 
‘→ information which includes phenotypes of patients as HPO terms, the 
‘→ clinical summary and additional comments.
IMPORTANT CONTEXT:
==================
The patient has genetic variants in each of the genes listed below. For
‘→ each gene, you will see:
1. A comprehensive gene report (including disease associations, phenotypes,
‘→ and clinical significance)
2. A list of the ACTUAL VARIANTS found in this patient within that gene,
‘→ with detailed information including:
                 - Genomic location and allele information
                 - Clinical annotations (ClinVar significance, ACMG classification, OMIM
                 ‘→ status)
                 - Population frequencies (gnomAD, 1000 Genomes)
Your task is to rank these eight genes based on their potential
‘→ significance to the patient’s condition.
CRITICAL EVALUATION CRITERIA:
==============================
When ranking each gene, you must consider BOTH:
1. The gene’s general disease associations and known phenotypes (from the
‘→ gene report)
2. The SPECIFIC VARIANTS found in this patient within that gene (variant
‘→ details provided)
A gene ranks higher if:
- Its known disease phenotypes align well with the patient’s symptoms
- The patient’s variants in this gene show pathogenic/likely pathogenic
‘→ ClinVar classifications
- The variants are rare in the population (low gnomAD frequencies)
- The ACMG classification supports pathogenicity
IMPORTANT: Ranking a gene as significant means you believe one or more of
‘→ the variants in that gene could be contributing to the patient’s
‘→ condition. The gene report provides context, but the actual variants in
‘→ the patient are what matter for diagnosis.
Patient Information:
{target}
Here are the 8 genes to rank (each with the patient’s variants listed):
---Gene 1 ---
Identifier: {item_1_id} 
Report: {item_1_info}
---Gene 2 ---
Identifier: {item_2_id} 
Report: {item_2_info}
---Gene 3 ---
Identifier: {item_3_id} 
Report: {item_3_info}
---Gene 4 ---
Identifier: {item_4_id} 
Report: {item_4_info}
---Gene 5 ---
Identifier: {item_5_id} 
Report: {item_5_info}
---Gene 6 ---
Identifier: {item_6_id} 
Report: {item_6_info}
---Gene 7 ---
Identifier: {item_7_id} 
Report: {item_7_info}
---Gene 8 ---
Identifier: {item_8_id} 
Report: {item_8_info}
Please rank all eight genes from 1 (most significant) to 8 (least
‘→ significant).
Return JSON with keys ‘rank_1’..’rank_8’, each containing only ‘gene_id’
‘→ (no explanation).
Return your response as pure JSON only. Do not wrap it in markdown code
‘→ blocks or use backticks.
~~~

### inGeneTopMatch prompt

For each variant surviving the Prelimin8 cut, the LLM composes a Detailed Variant Report by summarising the knowledge-graph evidence (overlapping genomic entities, ontology annotations, ClinVar and GWAS hits in the same entities). The placeholder {`variant_info_section}` is replaced with the inlined evidence at runtime.

~~~
Background: You are a genomic data analyst AI. Overall goal is to spot
‘→ pathogenic variants in a patient.
In this step, your job is to create a detailed report about the variant and
‘→ the genetic entities that are most likely to be affected by the
‘→ variant.
0a) Previously an LLM has provided us a relevant genetic entities (genes,
‘→ enhancers, promoters, transcription factors, etc.) that might be
‘→ affected by a variant in a pathogenic way.
0b) Then we have retrieved other variants on that entity which has known
‘→ GWAS associations and ClinVar entries.
-Now you will be given the variant and a list of genetic entities, each ‘→ entity will be provided with a list of GWAS associations and ClinVar ‘→ data for the variants on that entity.
1) Your job is to summarize the information, about the variant, genetic
‘→ entities and GWAS associations and ClinVar data of the variants on that
‘→ entity.
2a) Disease and phenotype information coming from GWAS and ClinVar data wi _⌋_ ‘→ ll be very important and should be included in the report. Any pattern ‘→ of phenotypes or diseases about a genetic entity is very important.
2b) Mention all the phenotypes and diseases that are associated.
3) Please mention any clue about the pathogenicity of the variant and its
‘→ relations to the disease and phenotypes based on GWAS and ClinVar data.
The report should include all the important information and should not be
‘→ too long.
{variant_info_section} Now create the report:
~~~

### Elimin8 prompt

Structurally identical to the Prelimin8 prompt (eight items, total ranking, JSON output) but ranks variants rather than genes and reasons over the Detailed Variant Reports.

~~~
You are a genetic assistant AI. You will be given genes and the patient 
‘→ information which includes phenotypes of patients as HPO terms, the 
‘→ clinical summary and additional comments.
IMPORTANT CONTEXT:
==================
The patient has genetic variants in each of the genes listed below. For
‘→ each gene, you will see:
1. A comprehensive gene report (including disease associations, phenotypes,
‘→ and clinical significance)
2. A list of the ACTUAL VARIANTS found in this patient within that gene,
‘→ with detailed information including:
                 - Genomic location and allele information
                 - Clinical annotations (ClinVar significance, ACMG classification, OMIM
                 ‘→ status)
                 - Population frequencies (gnomAD, 1000 Genomes)
Your task is to rank these eight genes based on their potential
‘→ significance to the patient’s condition.
CRITICAL EVALUATION CRITERIA:
==============================
When ranking each gene, you must consider BOTH:
1. The gene’s general disease associations and known phenotypes (from the
‘→ gene report)
2. The SPECIFIC VARIANTS found in this patient within that gene (variant
‘→ details provided)
A gene ranks higher if:
- Its known disease phenotypes align well with the patient’s symptoms
- The patient’s variants in this gene show pathogenic/likely pathogenic
‘→ ClinVar classifications
- The variants are rare in the population (low gnomAD frequencies)
- The ACMG classification supports pathogenicity
IMPORTANT: Ranking a gene as significant means you believe one or more of
‘→ the variants in that gene could be contributing to the patient’s
‘→ condition. The gene report provides context, but the actual variants in
‘→ the patient are what matter for diagnosis.
Patient Information:
{target}
Here are the 8 genes to rank (each with the patient’s variants listed):
---Gene 1 ---
Identifier: {item_1_id} 
Report: {item_1_info}
---Gene 2 ---
Identifier: {item_2_id} 
Report: {item_2_info}
---Gene 3 ---
Identifier: {item_3_id} 
Report: {item_3_info}
---Gene 4 ---
Identifier: {item_4_id} 
Report: {item_4_info}
---Gene 5 ---
Identifier: {item_5_id} 
Report: {item_5_info}
---Gene 6 ---
Identifier: {item_6_id} 
Report: {item_6_info}
---Gene 7 ---
Identifier: {item_7_id} 
Report: {item_7_info}
---Gene 8 ---
Identifier: {item_8_id} 
Report: {item_8_info}
Please rank all eight genes from 1 (most significant) to 8 (least
‘→ significant).
Return JSON with keys ‘rank_1’..’rank_8’, each containing only ‘gene_id’
‘→ (no explanation).
Return your response as pure JSON only. Do not wrap it in markdown code
‘→ blocks or use backticks.
~~~

### Final Tournament prompt

Pairwise comparison: given two variants and their Detailed Variant Reports the LLM returns a single winner identifier. Ties are disallowed so every call contributes a strict order.

~~~
You are a genetic assistant AI. You will be given two genetic variants
‘→ (mutations) and the patient information which includes phenotypes of
‘→ patients as HPO terms, the clinical summary and additional comments.
‘→ Variants are analyzed and annotated by our systems previous steps and a
#x2018;→ report was created for them which includes important genetic entities 
‘→ such as genes, transcripts, TF-binding sites, enhancers, ChromHMM
‘→ states, etc that the variant is located in. Also, other variants’ GWAS
‘→ associations and ClinVar annotations that are on the same entity are 
‘→ included.
Additionally, you will be provided with detailed biomedical reports for the
‘→ genes associated with each variant. These gene reports contain
‘→ comprehensive information about gene function, disease associations,
‘→ and clinical significance.
Your task is to compare these two genetic variants and determine which one
‘→ is more significant for the given patient.
When making your decision, it is crucial to consider the patient’s listed
‘→ phenotypes, epicrisis and comments. A variant is more significant if
‘→ its known clinical associations align well with the patient’s symptoms.
‘→ Conversely, it is less likely to be the cause if it is strongly
‘→ associated with phenotypes that the patient does not have.
Patient Information:
{target}
---
{item_1_id}:
Report: {item_1_info}
---
{item_2_id}:
Report: {item_2_info}
---
Please compare these two variants and declare a winner. You must always
‘→ choose a single winner; ties are not allowed.
Return your response as pure JSON only with a single key “winner_id”
‘→ containing the identifier of the winning variant. Do not wrap it in
‘→ markdown code blocks or use backticks.
~~~

### Item serialization

Within each prompt, individual variants and genes are rendered through fixed serialisation templates. Each variant occupies a single line containing its genomic coordinates, genotype, database identifiers, allele frequencies, ACMG and ClinVar assertions, and pathogenicity predictors. Each gene is rendered as its cached dossier followed by up to five of the patient’s variants within that gene.

~~~
**Variant template**.
{rank}. Variant: {chr}:{pos} {ref}>{alt} | Gene: {gene} | GT: {gt} | IDs:
‘→ {ids} | AF: 1KG={kg}, gnomAD={gad}, gnomADg={gadg} | ACMG: {acmg} |
‘→ ClinVar: {clinvar} | Scores: SIFT={sift}, Polyphen={polyphen},
‘→ CADD={cadd}, REVEL={revel}, SpliceAI={spliceai}, DANN={dann},
‘→ MetalR={metalr} | Alpha Missense: {am_score} ({am_pred}) | OMIM: {omim}
~~~

~~~
**Gene template**.
{gene_cache} \nVariants on this gene with their ranks: \n{variants}
~~~

## S6. Filtering cost-recall Pareto

DAVP’s per-sample LLM-call budget scales linearly with the size of the pre-filtered candidate gene set *G*^*′′*^ (Supplementary Table S1); the choice of pre-filter and of the top- *K* cut-off therefore directly trades downstream reasoning cost against the probability of retaining the causal gene. We characterise this trade-off across nine candidate pre-filter strategies and the grid *K* ∈ {8, 16, 32, 64, 128, 256, 512, 1024}.

### Candidate pre-filter families

We compare three families of gene-level pre-filters. The first is phenotype-aware: Exomiser with HPO terms supplied at input, LIRICAL, and Xrare. The second is the same Exomiser version run on the same VCFs with an empty HPO list, isolating the contribution of the phenotype prioritiser from the rest of Exomiser’s pipeline. The third is variant-level pathogenicity, aggregated to the gene by taking each gene’s maximum score: ACMG severity, AlphaMissense, REVEL, CADD, and DANN. The variant-level scores are computed on the full unfiltered per-sample annotated variant list (median ∼3.0 × 10^4^ variants per case from raw exome calls), *not* on the upstream Exomiser top-256 slice that DAVP itself consumes; this isolates each phenotype-agnostic baseline’s intrinsic gene-prioritisation ability from any benefit it would inherit from a phenotype-aware pre-sort. For each strategy we compute the rank of the answer gene in its full output list and report cumulative recall at *K* ∈ {8, 16, 32, 64, 128, 256, 512, 1024}.

### Cost axis

We translate each cut-off *K* into a per-sample LLM-call budget using the stage complexity formulas of Supplementary Table S1. With Prelimin8 reducing its input to *K*_p8_ = 64 when *K >* 64, inGeneTopMatch emitting one report per surviving gene, Elimin8 reducing its input to *K*_e8_ = 20, and the final tournament extracting the top-3, the total cost follows

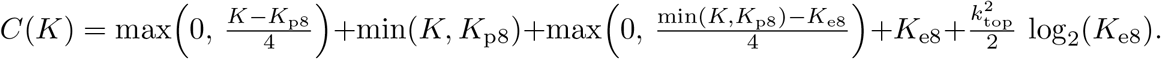

Evaluated at the paper’s *K* = 256 and calibrated to the measured per-stage means in Supplementary Table S12, this yields the per-cutoff cost reported in Supplementary Table S13 (a measured mean of 181 calls per sample at *K* = 256).

### Per-call token usage and dollar cost

Supplementary Table S14 resolves the per-call counts of Supplementary Table S12 into per-call token volumes and dollar cost by replaying twenty randomly sampled calls from each DAVP stage, reconstructed verbatim from the paper-run logs (8-way ranking groups for Prelimin8 and Elimin8, single-variant evidence sections for inGeneTopMatch, pairwise variant pairs for the Final Tournament). Prelimin8 dominates the per-call input budget at ∼24.7k tokens because each call carries eight full gene-cache dossiers plus up to five patient variants per gene. Elimin8 is roughly 1.8× smaller because the long gene dossiers are replaced by the more compact Detailed Variant Reports; inGeneTopMatch summarises a single variant’s evidence section (∼ 5.6k tokens) with reasoning disabled, and the Final Tournament compares only two variant reports (∼ 4.7k tokens). Reasoning consumes the bulk of output-side tokens for the three ranking stages (the residual is the JSON ranking) and is exactly zero for inGeneTopMatch by configuration. Translated through the public Gemini 2.5 Flash list price (input $0.30 / 1M tokens, output $2.50 / 1M tokens), this yields a mean per-call cost of $0.026 (Prelimin8), $0.005 (inGeneTopMatch), $0.020 (Elimin8) and $0.008 (Final Tournament); the per-cutoff total per-sample cost is reported in Supplementary Table S13.

**Table S13:** Per-sample LLM call count and dollar cost as a function of the gene-level pre-filter cutoff *N*_g_, calibrated to the measured per-stage means at *N*_g_ = 256 reported in Supplementary Table S12. Per-stage costs scale with the input sizes set by the upstream stages: Prelimin8 with 0.89 · max(0, (*N*_g_ − *K*_p8_)*/*4) calls; inGeneTopMatch with 1.27 calls per top-min(*N*_g_, *K*_p8_) gene (1.27 variants per gene observed); Elimin8 with 2.20 · max(0, (iGTM calls − *K*_e8_)*/*4) calls; the Final Tournament with 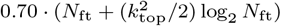 calls where *N*_ft_ = min(*K*_e8_, iGTM calls) and *k*_top_ = 3. Stages whose input is at or below their target size are skipped (entries shown as 0). The total cost column applies the per-call mean from Supplementary Table S14 (Prelimin8 $0.02642, inGeneTopMatch $0.00505, Elimin8 $0.01983, Final Tournament $0.00846) at the public Gemini 2.5 Flash list price. The paper configuration is *N*_g_ = 256 (bold row); the bold-row stage entries match the measured means in Table S12 (the row total of 186 reflects the sum of rounded stage means; the cohort-pooled total mean of 181 differs by ≈5 calls because per-case totals exclude cases that fail before the last stage).

| Pre-filter cutoff $N_g$ | Prelimin8 | inGeneTopMatch | Elim8 | Final Tournament | Total calls | Total cost (USD) |
| --- | --- | --- | --- | --- | --- | --- |
| 8 | 0 | 10 | 0 | 17 | 27 | 0.194 |
| 16 | 0 | 20 | 0 | 28 | 48 | 0.338 |
| 20 | 0 | 25 | 3 | 28 | 56 | 0.423 |
| 32 | 0 | 41 | 12 | 28 | 81 | 0.682 |
| 64 | 0 | 81 | 34 | 28 | 143 | 1.320 |
| 128 | 14 | 81 | 34 | 28 | 157 | 1.690 |
| **256** | **43** | **81** | **34** | **28** | **186** | **2.456** |
| 512 | 100 | 81 | 34 | 28 | 243 | 3.963 |
| 1024 | 214 | 81 | 34 | 28 | 357 | 6.975 |

**Table S14:**
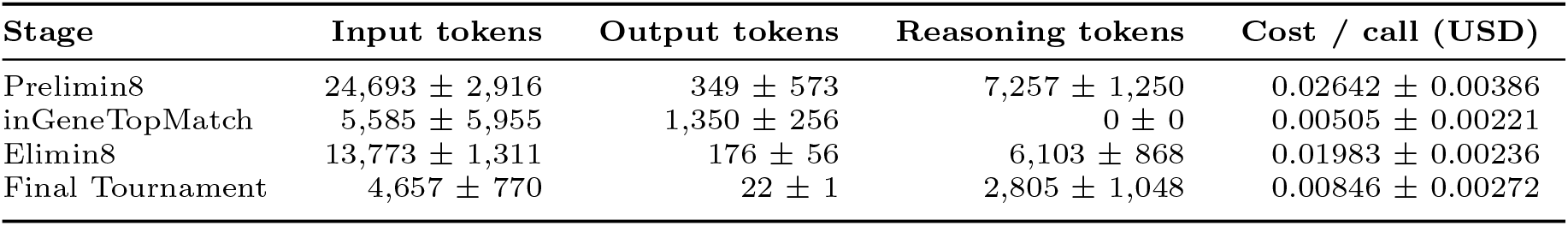
Per-call token usage and dollar cost by DAVP stage, given as mean ±s.d. over twenty calls drawn at random per stage from the paper-run logs (8-way ranking groups for Prelimin8 and Elimin8, single-variant evidence sections for inGeneTopMatch, pairwise variant pairs for the Final Tournament) and replayed against Gemini 2.5 Flash with an 8,192-token thinking budget for the three ranking stages and thinking disabled for inGeneTopMatch (thinking_config.thinking_budget is set explicitly per call to match the paper configuration). The output-token column reports the visible response text only; reasoning tokens are reported separately. Dollar cost is computed at the public Gemini 2.5 Flash list price (input $0.30 / 1M tokens, both visible output and reasoning tokens billed as output at $2.50 / 1M). Per-cutoff total per-sample cost is reported in Supplementary Table S13.

### Pareto frontier

Supplementary Figure S4 lays out recall against *K* (panel a), recall against the theoretical downstream LLM budget (panel b), and the dataset-level breakdown of the Exomiser curve (panel c). Three findings are consistent across all three views. First, no phenotype-agnostic pathogenicity score approaches the phenotype-aware ceiling at the operationally relevant small-*K* regime when scored on the raw exome output: ACMG severity tops the agnostic family at 54.6% recall at *K* = 256, CADD reaches 47.4%, REVEL 36.7%, AlphaMissense 34.1%, and DANN only 17.1%, all far below the phenotype-aware band of 91–97.7%. The two missense-only predictors (AlphaMissense, REVEL) cap below 50% by construction because non-missense causal variants are unscored and rank at the tail. Second, the phenotype-aware alternatives LIRICAL and Xrare are the best pre-filters at very small *K* but plateau early; Exomiser continues to accumulate recall out to *K* ≈ 256 and reaches the highest plateau (97.7% vs. 91–92%). Third, a *K* = 64 Exomiser cut-off would cost only ∼114 calls per sample but throws away 5 percentage points of filter recall; a *K* = 1024 cut-off would buy nothing beyond what *K* = 256 already delivers. The paper’s choice of *K* = 256 sits on the knee of the Exomiser-with-HPO curve.

**Table S15:** Gene-level filter recall across the 709-case benchmark for each candidate pre-filter strategy, evaluated at four representative cut-offs. Phenotype-aware tools (Exomiser, LIRICAL, Xrare) score every gene against the patient’s HPO terms; phenotype-agnostic tools (ACMG severity, AlphaMissense, REVEL, CADD, DANN) rank by the per-gene maximum of the listed variant-level pathogenicity score, computed on the full unfiltered per-sample variant list. **Retained** = number of cases for which the answer gene appears anywhere in the filter’s ranked output; the recall denominator is the full 709-case benchmark, so cases where the score is undefined for the causal variant type (e.g. AlphaMissense and REVEL are missense-only) appear as zeros at every *K*. The phenotype-aware tools cluster in the high-90% region; the single-score predictors never approach that ceiling even at *K* = 512. The paper’s configuration is *K* = 256 under Exomiser (with HPO). Bold marks the best strategy in each column.

| Filter strategy | Retained | $K=64$ | $K=128$ | $K=256$ | $K=512$ |
| --- | --- | --- | --- | --- | --- |
| Exomiser (with HPO) | 693/709 | **92.5%** | **96.9%** | **97.7%** | **97.7%** |
| Exomiser (no HPO) | **698/709** | 83.5% | 88.3% | 93.8% | 96.3% |
| LIRICAL | 646/709 | 90.8% | 91.1% | 91.1% | 91.1% |
| Xrare | 653/709 | 91.8% | 92.1% | 92.1% | 92.1% |
| ACMG Severity | 651/709 | 52.6% | 54.2% | 54.6% | 57.0% |
| AlphaMissense | 448/709 | 27.5% | 29.6% | 34.1% | 38.5% |
| REVEL | 447/709 | 24.8% | 31.5% | 36.7% | 42.5% |
| CADD | 540/709 | 20.9% | 35.1% | 47.4% | 56.4% |
| DANN | 539/709 | 6.1% | 11.0% | 17.1% | 30.9% |

**Fig. S4:**
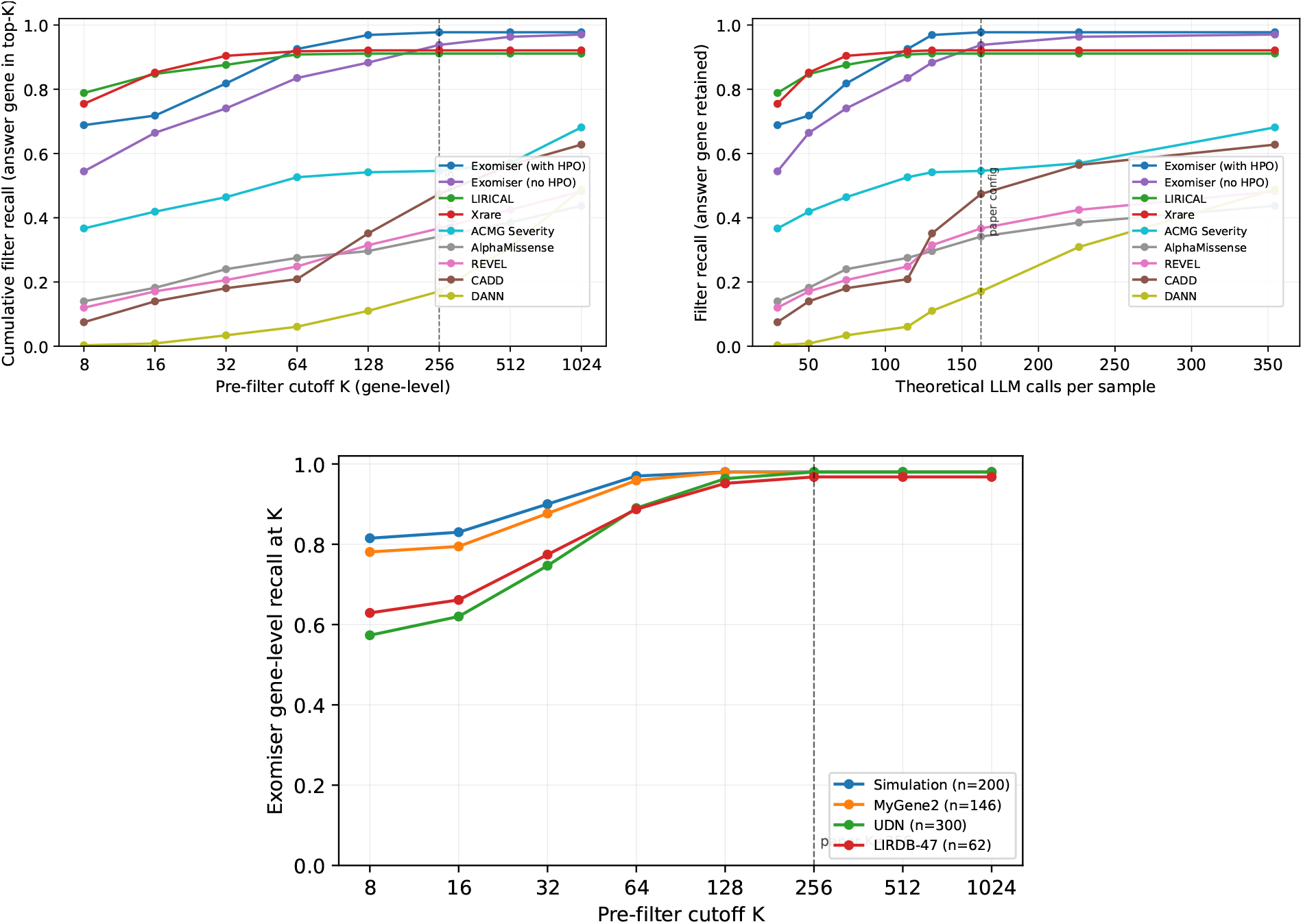
Filtering cost-recall Pareto across the 709-case benchmark. (a) Gene-level recall as a function of the pre-filter cut-off *K* for each candidate strategy; phenotype-aware tools dominate pathogenicity-only baselines across the plotted range. Dashed vertical line marks the paper’s *K* = 256. (b) Same curves re-plotted against the theoretical downstream LLM-call budget per sample. (c) Exomiser recall-vs-*K* broken out by dataset.

**Table S16:** Variant-level top-*k* recall for purely phenotype-agnostic baselines, compared to DAVP’s phenotype-aware full pipeline. Single-score rankers (CADD, REVEL, AlphaMissense, DANN, ACMG severity) sort each case’s full unfiltered annotated variant list (median ∼3.0 × 10^4^ variants per case from raw exome calls) by the listed score alone; variants missing a score are treated as worst-possible and within-score ties are broken by the original variant ordering. **Exomiser (no HPO)** runs the full Exomiser exome filter with an empty hpoIds list (frequency, inheritance, OMIM-prioritiser, and variant pathogenicity machinery all retained, only the phenotype prioritiser removed); its variant ranking is obtained by concatenating per-gene contributing variants in Exomiser’s gene-rank order, then ordering within each gene by variantScore. DAVP outperforms every phenotype-agnostic baseline at every *k* by a wide margin, confirming that even a sophisticated exome filter without phenotype input cannot substitute for patient-specific reasoning.

| Method | $n$ | Top-1 | Top-3 | Top-5 | Top-10 | Top-20 |
| --- | --- | --- | --- | --- | --- | --- |
| DAVP | 687 | **71.9** | **88.6** | **91.4** | **94.5** | **96.1** |
| Exomiser (no HPO) | 707 | 13.4 | 27.9 | 35.2 | 47.5 | 60.1 |
| CADD | 708 | 1.3 | 3.1 | 5.2 | 10.6 | 18.2 |
| REVEL | 708 | 4.8 | 7.2 | 9.0 | 14.0 | 18.9 |
| AlphaMissense | 708 | 6.4 | 10.7 | 12.7 | 15.7 | 20.1 |
| DANN | 708 | 0.1 | 0.1 | 0.1 | 0.6 | 1.1 |
| ACMG severity | 708 | 22.5 | 37.6 | 42.4 | 48.2 | 52.4 |

## S7. Phenotype-agnostic baselines

To isolate the contribution of phenotype-aware reasoning, we compare DAVP against a family of purely phenotype-agnostic baselines. The single-score rankers (CADD, REVEL, AlphaMissense, DANN, ACMG severity) sort each case’s full unfiltered annotated variant list (median ∼3.0 × 10^4^ variants per case from raw exome calls, prior to any phenotype-aware pre-filter) by one scalar variant score and report the rank of the answer variant. We additionally include an Exomiser-no-HPO row: the full Exomiser exome filter rerun with an empty hpoIds list, retaining the frequency, inheritance, OMIM-prioritiser, and variant pathogenicity machinery while removing only the phenotype prioritiser. No HPO terms, epicrisis, or patient-specific reasoning are used by any of these baselines.

Supplementary Table S16 reports the resulting top-*k* recall. DAVP outperforms every phenotype-agnostic baseline at every *k* by a wide margin: DAVP recovers 71.9% of answer variants at rank 1 and 88.6% at top-3, versus 22.5% and 37.6% for the strongest single-score ranker (ACMG severity), 13.4% and 27.9% for Exomiser-no-HPO, and below 10% top-1 for CADD, REVEL, AlphaMissense, and DANN. Notably, Exomiser-no-HPO – which retains every Exomiser stage except phenotype matching – still trails DAVP by roughly 60 percentage points at top-3, confirming that even a sophisticated exome-filter pipeline cannot substitute for patient-specific phenotype reasoning when the candidate pool is the full exome.

**Fig. S5:**
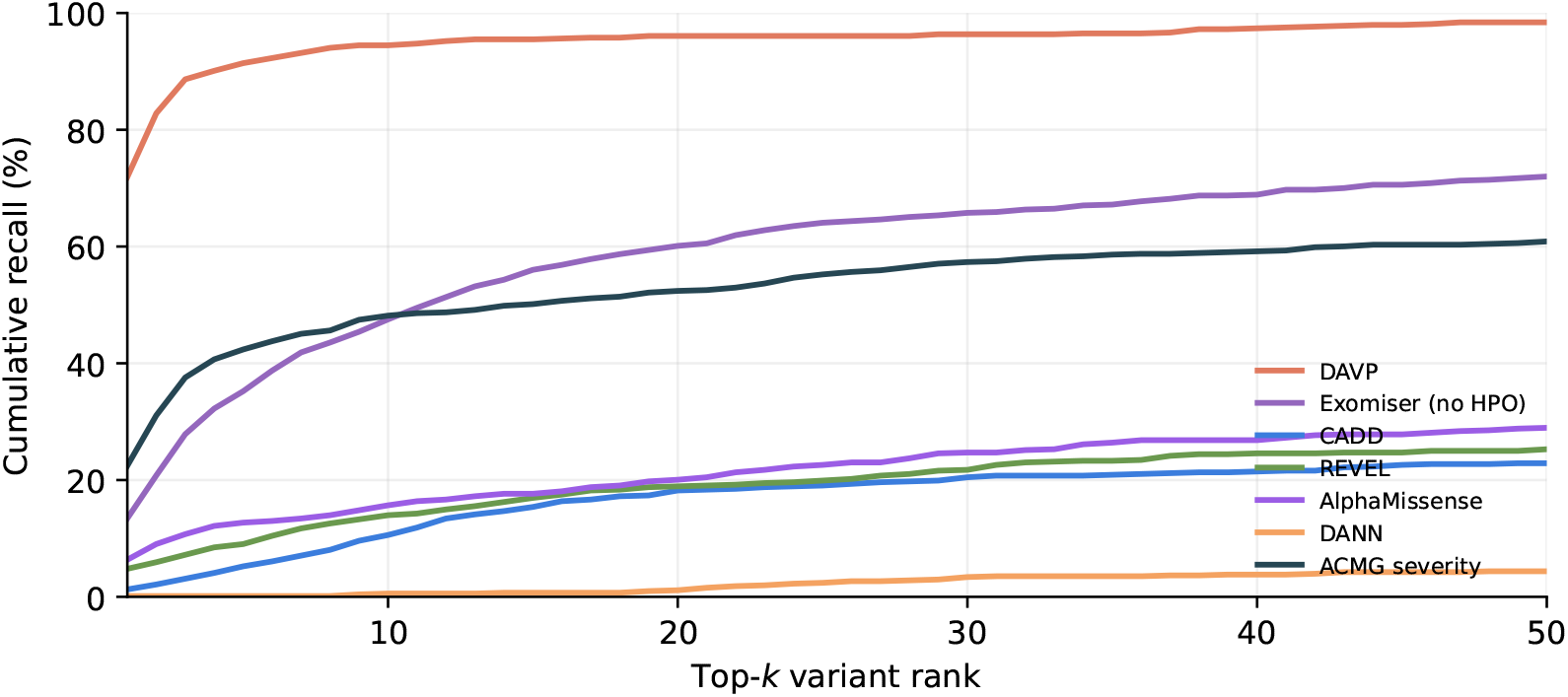
Cumulative variant-level recall as a function of rank for DAVP versus the phenotype-agnostic baselines (Exomiser-no-HPO, CADD, REVEL, AlphaMissense, DANN, ACMG severity). DAVP’s curve sits roughly an order of magnitude above the next-best baseline at every rank cutoff. Numerical top-*k* values for *k* ∈ {1, 3, 5, 10, 20} are tabulated in Supplementary Table S16.

**Table S17:** Top-3 recall (%) of three ablation variants across the four benchmark datasets, stratified by ClinVar significance class of the answer variant. **DAVP (Prelimin8)** reports *gene-level* top-3 recall, i.e. the fraction of cases in which the answer gene appears in the top 3 of the Prelimin8 gene ranking; the remaining two variants report *variant-level* top-3 recall (answer variant in the top 3 of the variant ranking). **DAVP (Prelimin8 + Elimin8)** adds inGeneTopMatch and the Elimin8 variant-ranking stage but skips the Final Tournament; **DAVP (full)** is the four-stage paper pipeline. *Pathogenic* aggregates ClinVar `pathogenic` and `likely pathogenic`; *VUS* aggregates `uncertain` and `conflicting`. LIRDB-47 variants lack ClinVar labels and are reported only in the All row.

| Cohort | Class | DAVP (Prelimin8) | DAVP (Prelimin8 + Elimin8) | DAVP (full) |
| --- | --- | --- | --- | --- |
| Simulation | Pathogenic | 34.7 | 91.8 | **98.0** |
| MyGene2 | Pathogenic | 33.1 | 95.5 | **99.2** |
|  | VUS | 25.0 | 85.7 | **100.0** |
|  | All | 32.9 | 93.6 | **97.9** |
| UDN | Pathogenic | 32.4 | 74.1 | **84.4** |
|  | VUS | 24.7 | 60.6 | **73.2** |
|  | All | 28.6 | 67.5 | **78.9** |
| LIRDB-47 | All | 24.2 | 58.1 | **77.4** |
| All | Pathogenic | 33.5 | 87.4 | **94.1** |
|  | VUS | 24.7 | 61.7 | **74.5** |
|  | All | 30.8 | 78.9 | **88.1** |

## S8. Stage-level ablation

To quantify the contribution of each LLM-driven stage, we evaluate three pipeline variants on every case in the 709-case benchmark, each adding one stage on top of the previous one to form a monotone lattice: DAVP (Prelimin8) stops after gene-level Prelimin8 ranking; DAVP (Prelimin8 + Elimin8) runs Prelimin8, builds detailed variant reports via inGeneTopMatch, and ranks variants with Elimin8; DAVP (full) runs the full four-stage pipeline.

Gene-level Prelimin8 alone recovers 30.8% of answer genes at top-3 across the full benchmark. Adding variant-level reasoning via Elimin8 brings top-3 recall to 78.9%, a 48.1-point absolute gain and the largest single-stage contribution. The Final Tournament adds a further 9.2 percentage points overall (full pipeline 88.1% top-3), with the largest effect concentrated in the two hardest cohorts: LIRDB-47 (58.1% to 77.4%) and UDN (67.5% to 78.9%). On Simulation and MyGene2 the Prelimin8 + Elimin8 checkpoint is already close to ceiling and the Final Tournament mostly reorders cases that Elimin8 already placed in the top few. The full per-dataset rank distributions are shown in Supplementary Figure S6.

### Hyperparameter sweep

Beyond the three-stage truncation lattice above, we additionally swept the per-stage base model, rounds-before-elimination (RBE), and extended-reasoning thinking budget (TB) on the held-out UDN-VUS subset (*n* = 108) to select the paper’s final configuration. Supplementary Table S18 reports gene-level recall after Prelimin8 and variant-level recall after Elimin8 for each combination. The chosen configuration (Gemini 2.5 Flash, RBE=1, TB=8192 at both stages) trades a modest top-5 gene-recall plateau for the largest variant-recall gain (top-5 jumping from 28.7–50.0% to 67.6%), confirming that an extended-reasoning budget at Elimin8 is the operative driver of variant-level discrimination.

**Fig. S6:**
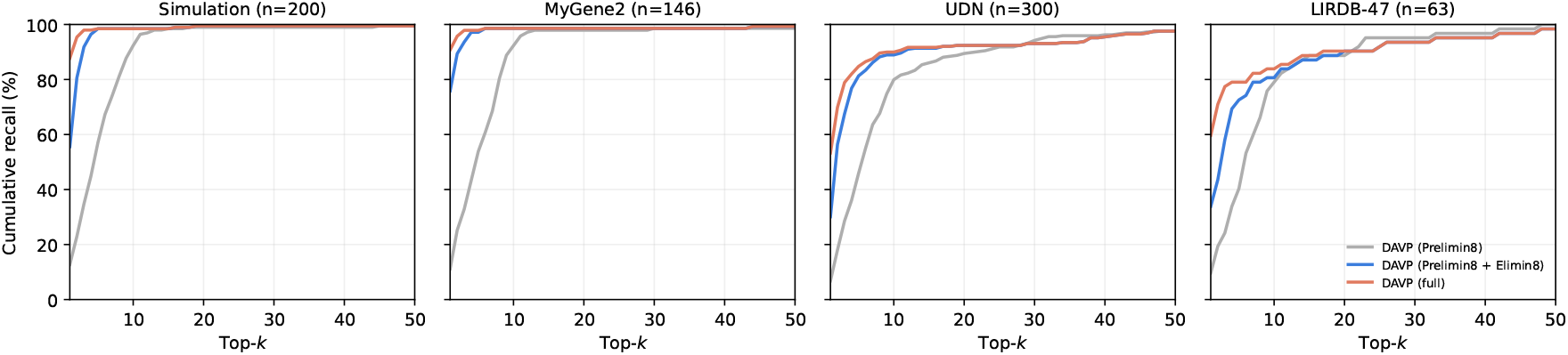
Cumulative distribution of the answer-variant rank (or answer-gene rank, for the Prelimin8-only variant) for the three ablation variants, shown separately for each of the four benchmarks. Higher curves are better. Variant-level reasoning via Elimin8 provides the largest recall jump over gene-level Prelimin8 alone on every dataset. The Final Tournament contributes an additional gain of a few percentage points at top-3, with the largest contribution on the two harder cohorts (LIRDB-47: 58.1% to 77.4%; UDN: 67.5% to 78.9%). Curves correspond to the same three variants as Supplementary Table S17: DAVP (Prelimin8), DAVP (Prelimin8 + Elimin8), DAVP (full).

**Table S18:**
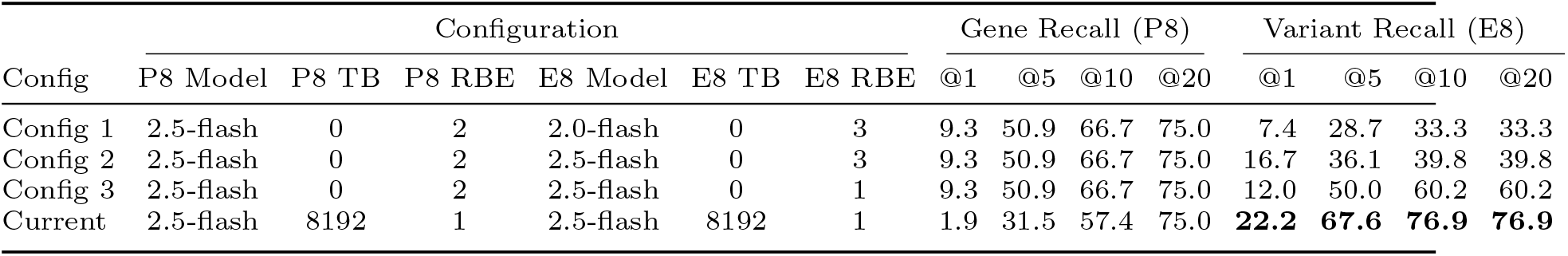
Hyperparameter sweep across DAVP configurations on the held-out UDN VUS subset (*n* = 108). Best Variant Recall column bolded. P8=Prelimin8, E8=Elimin8, RBE=rounds before elimination, TB=thinking budget.

**Table S19:** Gene-Level Top-k recall performance of DAVP compared to baseline methods.

| Dataset | Method | Top-1 | **Top-3** | Top-5 | Top-10 | **Top-20** | Top-50 |
| --- | --- | --- | --- | --- | --- | --- | --- |
| Overall | DAVP | **69.39** | **85.61** | **88.58** | **91.11** | **93.09** | **96.19** |
|  | Xrare | 25.25 | 49.51 | 64.60 | 80.54 | 87.17 | 91.96 |
|  | Lirical | 59.24 | 72.36 | 76.45 | 80.54 | 85.33 | 90.27 |
|  | Exomiser | 55.29 | 65.44 | 68.27 | 69.53 | 74.05 | 90.55 |
| LIRDB-47 | DAVP | **60.32** | **77.78** | **79.37** | **84.13** | **92.06** | **96.83** |
|  | Xrare | 17.46 | 33.33 | 41.27 | 66.67 | 76.19 | 84.13 |
|  | Lirical | 26.98 | 44.44 | 53.97 | 68.25 | 82.54 | 93.65 |
|  | Exomiser | 38.10 | 53.97 | 63.49 | 63.49 | 66.67 | 88.89 |
| Simulation | DAVP | **86.00** | **96.00** | **96.50** | **96.50** | **97.50** | **97.50** |
|  | Xrare | 45.50 | 77.50 | 89.50 | 95.50 | 96.00 | 96.00 |
|  | Lirical | 62.50 | 82.50 | 87.00 | 90.50 | 94.50 | 97.00 |
|  | Exomiser | 69.50 | 80.00 | 81.50 | 81.50 | 83.50 | 94.50 |
| MyGene2 | DAVP | **87.67** | **94.52** | **94.52** | **95.89** | **95.89** | **97.26** |
|  | Xrare | 37.67 | 73.29 | 92.47 | 94.52 | **95.89** | 96.58 |
|  | Lirical | 85.62 | 91.10 | 93.84 | **95.89** | **95.89** | **97.26** |
|  | Exomiser | 58.90 | 73.29 | 77.40 | 78.08 | 80.82 | 95.21 |
| UDN | DAVP | **51.33** | **76.00** | **82.33** | **86.67** | **89.00** | **94.67** |
|  | Xrare | 7.33 | 22.67 | 39.33 | 66.67 | 79.33 | 88.67 |
|  | Lirical | 51.00 | 62.33 | 65.67 | 69.00 | 74.67 | 81.67 |
|  | Exomiser | 47.67 | 54.33 | 56.00 | 58.67 | 66.00 | 88.00 |

## S9. Top-*k* recall by dataset and method

This section reports the headline gene-level and variant-level top-*k* recall numbers underlying the main-text Figure 2 CDFs and the per-dataset claims, along with the per-case head-to-head win/tie/loss breakdown.

Supplementary Table S19 reports top-*k* gene-level recall (*k* ∈ {1, 3, 5, 10, 20, 50}) for DAVP against three phenotype-driven baselines (Exomiser, LIRICAL, Xrare), pooled across the 709-case benchmark and broken out by dataset. DAVP is best at every k on every cohort except for two ties at large k on MyGene2 (Top-10/20 against LIRICAL).

Supplementary Table S20 reports the analogous variant-level recall and Supplementary Table S21 the per-case head-to-head win/tie/loss percentages versus each baseline.

**Table S20:** Variant-Level Top-k recall performance of DAVP compared to baseline methods.

| Dataset | Method | Top-1 | **Top-3** | Top-5 | Top-10 | **Top-20** | Top-50 |
| --- | --- | --- | --- | --- | --- | --- | --- |
| Overall | DAVP | **69.25** | **85.47** | **88.15** | **90.83** | **92.67** | **95.63** |
|  | Xrare | 25.25 | 49.22 | 64.32 | 80.11 | 87.02 | 91.54 |
|  | Lirical | 58.39 | 63.61 | 65.87 | 70.10 | 74.47 | 79.27 |
|  | Exomiser | 54.87 | 64.32 | 66.85 | 68.55 | 71.51 | 83.64 |
| LIRDB-47 | DAVP | **58.73** | **76.19** | **77.78** | **82.54** | **88.89** | **96.83** |
|  | Xrare | 17.46 | 31.75 | 41.27 | 61.90 | 74.60 | 80.95 |
|  | Lirical | 23.81 | 31.75 | 34.92 | 44.44 | 55.56 | 68.25 |
|  | Exomiser | 34.92 | 47.62 | 53.97 | 58.73 | 63.49 | 65.08 |
| Simulation | DAVP | **86.00** | **96.00** | **96.50** | **96.50** | **97.50** | **97.50** |
|  | Xrare | 45.50 | 77.00 | 89.00 | 95.50 | 96.00 | 96.00 |
|  | Lirical | 61.50 | 68.00 | 71.50 | 76.50 | 82.00 | 88.50 |
|  | Exomiser | 69.00 | 79.00 | 81.00 | 81.50 | 83.00 | 92.50 |
| MyGene2 | DAVP | **87.67** | **94.52** | **94.52** | **95.21** | 95.21 | 95.89 |
|  | Xrare | 37.67 | 73.29 | 92.47 | 94.52 | **95.89** | **96.58** |
|  | Lirical | 85.62 | 88.36 | 89.04 | 91.10 | 93.15 | 95.21 |
|  | Exomiser | 58.90 | 73.29 | 76.03 | 77.40 | 79.45 | 89.73 |
| UDN | DAVP | **51.33** | **76.00** | **81.67** | **86.67** | **89.00** | **94.00** |
|  | Xrare | 7.33 | 22.67 | 39.00 | 66.67 | 79.33 | 88.33 |
|  | Lirical | 50.33 | 55.33 | 57.33 | 61.00 | 64.33 | 67.67 |
|  | Exomiser | 47.67 | 53.67 | 55.67 | 57.67 | 61.67 | 78.67 |

**Table S21:**
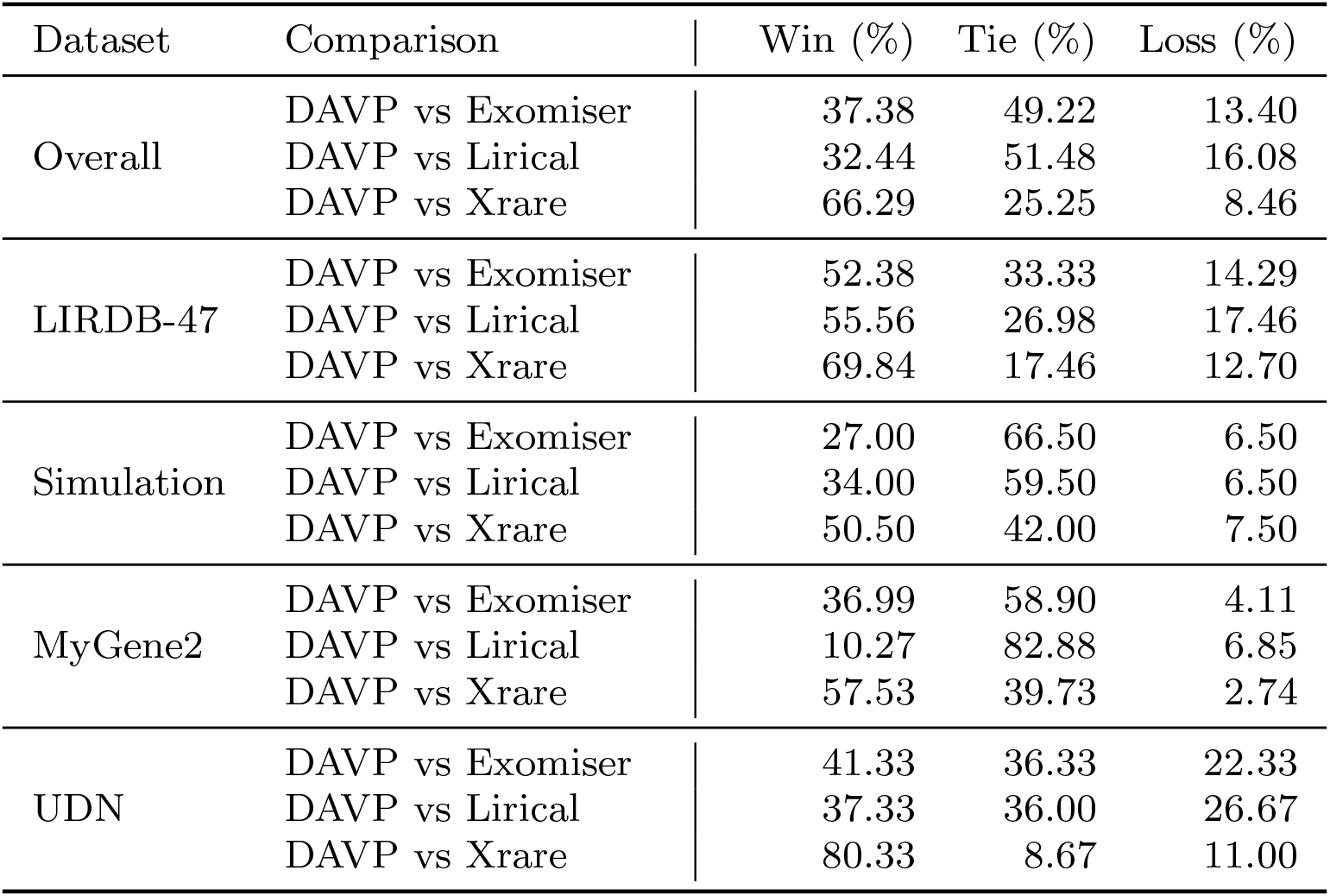
Head-to-Head performance comparison of DAVP versus baseline methods.

**Table S22:**
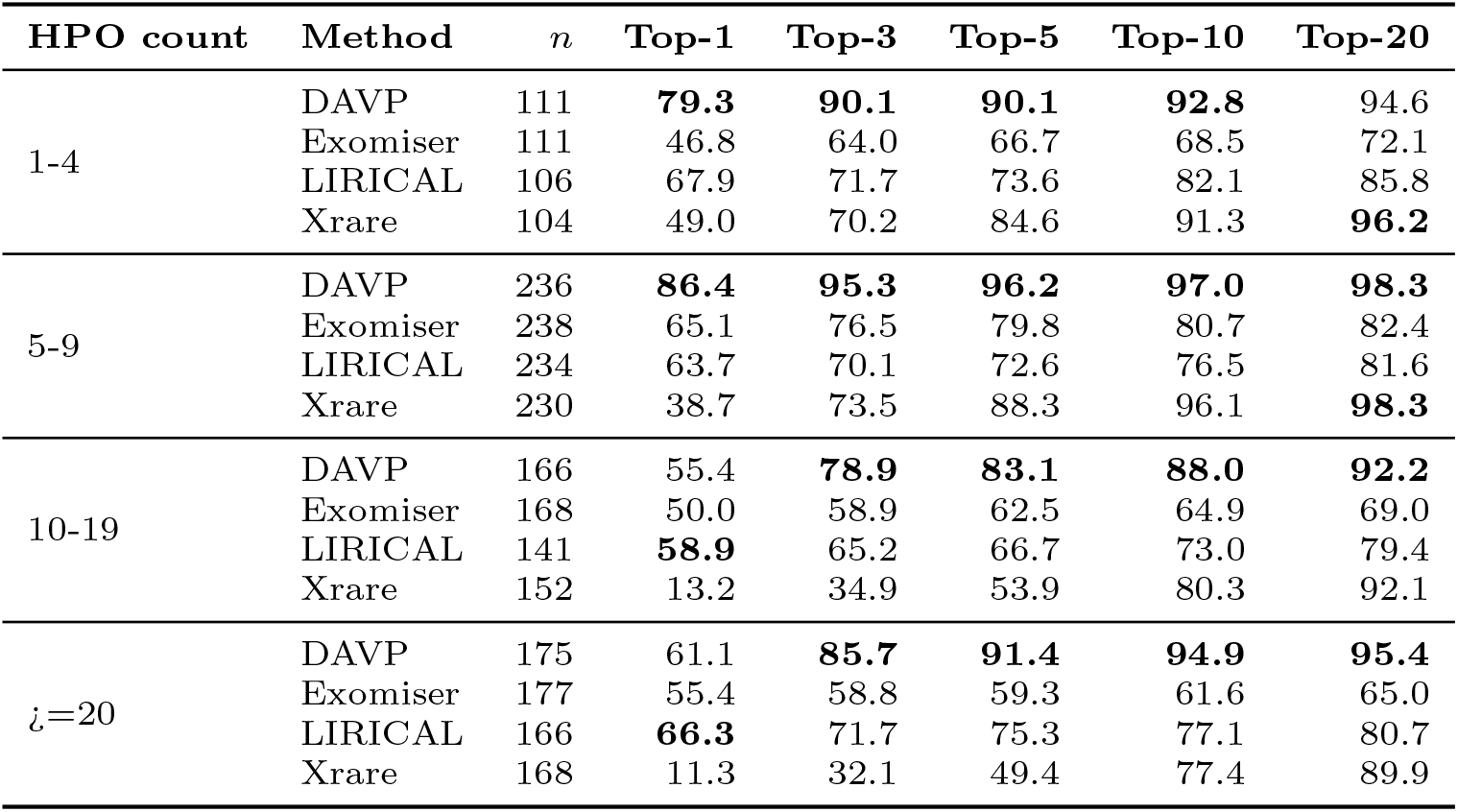
Top-*k* variant recall stratified by the number of HPO terms supplied at input. Each HPO count bucket lists all four compared methods; bold marks the best value in each column (ties are bolded jointly).

## S10. Phenotype and variant class stratification

This section stratifies top-*k* recall on five axes: the number of HPO terms supplied at input, the information content of those terms, the molecular consequence class of the answer variant, the ClinVar significance class (Pathogenic/Likely Pathogenic vs. VUS), and the ClinVar review-status star rating. Each axis is reported for all four methods (DAVP, Exomiser, LIRICAL, Xrare).

### Phenotype annotation breadth

Supplementary Table S22 reports top-*k* recall as a function of the number of HPO terms supplied per case (bins 1–4, 5–9, 10–19, ≥20). DAVP is the only method that maintains near-constant top-5 recall (≥ 85%) from the sparsest to the richest phenotype input. At the sparsest end (1–4 HPO terms), DAVP’s top-5 recall exceeds Exomiser by over ≥20 percentage points, suggesting that free-text epicrisis reasoning compensates for thin HPO input in a way that structured phenotype matchers cannot.

### HPO-count companion: gene-level recall

Supplementary Table S23 and Supplementary Figure S7 report the gene-level recall companion to the variant-level table above, switching the metric to top-*k gene* recall under the same per-case HPO-count stratification.

### Phenotype specificity

HPO terms vary by orders of magnitude in how specifically they identify a disease. We quantify specificity by computing per-term information content as − ln(*f*), where *f* is the fraction of OMIM, Orphanet, and DECIPHER diseases annotated with the term or any descendant in the HPO DAG (units: nats). Each case’s mean HPO-IC is then computed and the full 709-case benchmark partitioned into three equal-size terciles. The empirical tercile cut-points are: Low IC ≤ 3.91 nats (mostly broad constitutional terms such as “intellectual disability” or “seizure”), Mid IC (3.91, 4.64] nats, and High IC > 4.64 nats (highly specific terms typical of named syndromic findings); per-case mean IC across the full benchmark spans 1.23 to 7.86 nats with median 4.25.

**Table S23:** Gene-level top-*k* recall stratified by the number of HPO terms supplied per case. Companion to Supplementary Figure S7, which visualises the top-10 column. Recall denominator per (bin, method) is the count of cases in that bin for which the method returned a finite gene rank. Bold marks the best method in each (bin, top-*k*) cell.

| HPO count | Method | $n$ | Top-1 | Top-3 | Top-5 | Top-10 | Top-20 |
| --- | --- | --- | --- | --- | --- | --- | --- |
| 1-5 | DAVP | 182 | **84.1** | **92.9** | **93.4** | 95.1 | 96.7 |
|  | Exomiser | 181 | 55.8 | 71.8 | 75.1 | 75.7 | 78.5 |
|  | LIRICAL | 178 | 70.8 | 83.7 | 88.2 | 92.1 | 95.5 |
|  | Xrare | 173 | 46.2 | 72.3 | 86.7 | **96.5** | **97.7** |
| 6-10 | DAVP | 183 | **82.5** | **94.0** | **94.5** | **96.2** | **97.8** |
|  | Exomiser | 183 | 63.9 | 74.3 | 78.1 | 78.7 | 82.0 |
|  | LIRICAL | 175 | 61.1 | 78.3 | 82.9 | 89.7 | 96.6 |
|  | Xrare | 176 | 35.8 | 72.7 | 86.4 | 93.8 | 97.7 |
| 11-20 | DAVP | 177 | 54.8 | **77.4** | **82.5** | **87.0** | 91.0 |
|  | Exomiser | 177 | 49.7 | 59.3 | 62.7 | 64.4 | 71.2 |
|  | LIRICAL | 148 | **58.8** | 73.6 | 81.1 | 85.1 | 91.2 |
|  | Xrare | 157 | 12.7 | 31.2 | 52.2 | 80.9 | **92.4** |
| >20 | DAVP | 153 | 59.5 | **84.3** | **90.8** | **93.5** | **94.1** |
|  | Exomiser | 153 | 56.2 | 60.8 | 61.4 | 64.1 | 69.9 |
|  | LIRICAL | 147 | **68.0** | 80.3 | 81.6 | 84.4 | 89.1 |
|  | Xrare | 148 | 10.8 | 33.1 | 50.0 | 75.7 | 89.2 |

**Table S24:** Top-*k* variant recall stratified by mean disease information content (IC, in nats) of the case’s HPO list. Cases are split into three equal-size terciles: lower ≤ 3.91, middle (3.91, 4.64], upper *>* 4.64 nats.

| HPO IC tercile (nats) | Method | $n$ | Top-1 | Top-3 | Top-5 | Top-10 | Top-20 |
| --- | --- | --- | --- | --- | --- | --- | --- |
| $IC \leq 3.91$ | DAVP | 231 | **73.2** | **91.8** | **94.4** | **96.1** | **98.3** |
|  | Exomiser | 231 | 59.3 | 74.5 | 77.1 | 78.8 | 80.5 |
|  | LIRICAL | 225 | 73.8 | 77.3 | 79.6 | 84.0 | 88.0 |
|  | Xrare | 219 | 28.3 | 58.4 | 74.4 | 90.4 | 95.9 |
| $IC 3.91-4.64$ | DAVP | 227 | **70.5** | **85.0** | **89.0** | **92.5** | **93.8** |
|  | Exomiser | 232 | 50.0 | 55.6 | 56.9 | 58.2 | 63.4 |
|  | LIRICAL | 211 | 66.4 | 71.6 | 73.5 | 76.3 | 80.6 |
|  | Xrare | 218 | 22.5 | 48.6 | 64.7 | 81.7 | 92.2 |
| $IC > 4.64$ | DAVP | 230 | **70.4** | **87.4** | **89.1** | **92.2** | **94.3** |
|  | Exomiser | 231 | 58.9 | 67.1 | 71.0 | 73.2 | 75.3 |
|  | LIRICAL | 211 | 51.2 | 59.7 | 63.0 | 69.7 | 75.8 |
|  | Xrare | 217 | 31.3 | 53.0 | 70.0 | 88.5 | 94.9 |

Supplementary Table S24 shows that DAVP’s advantage over the baselines widens as phenotype specificity increases, particularly on the high-IC tercile where small differences in interpretation can shift discrimination between similar diseases. The effect is most pronounced at top-1, where DAVP exceeds the best baseline by 15–20 percentage points on the high-IC tercile.

### Molecular consequence class

We resolve each answer variant’s molecular consequence by looking it up in the ClinVar *MC* field (Sequence Ontology) and assigning a coarse class: missense, stop gained, frameshift, splice, start lost, inframe indel, synonymous, UTR, intronic, non-coding transcript, or other. All 200 Simulation, 300 UDN, and 145 of 146 MyGene2 answer variants resolve cleanly. For LIRDB-47, 39 of 63 cases resolve to a ClinVar entry; the remaining 24 are clinically validated Turkish variants not yet deposited in ClinVar and are excluded from this stratification.

**Fig. S7:**
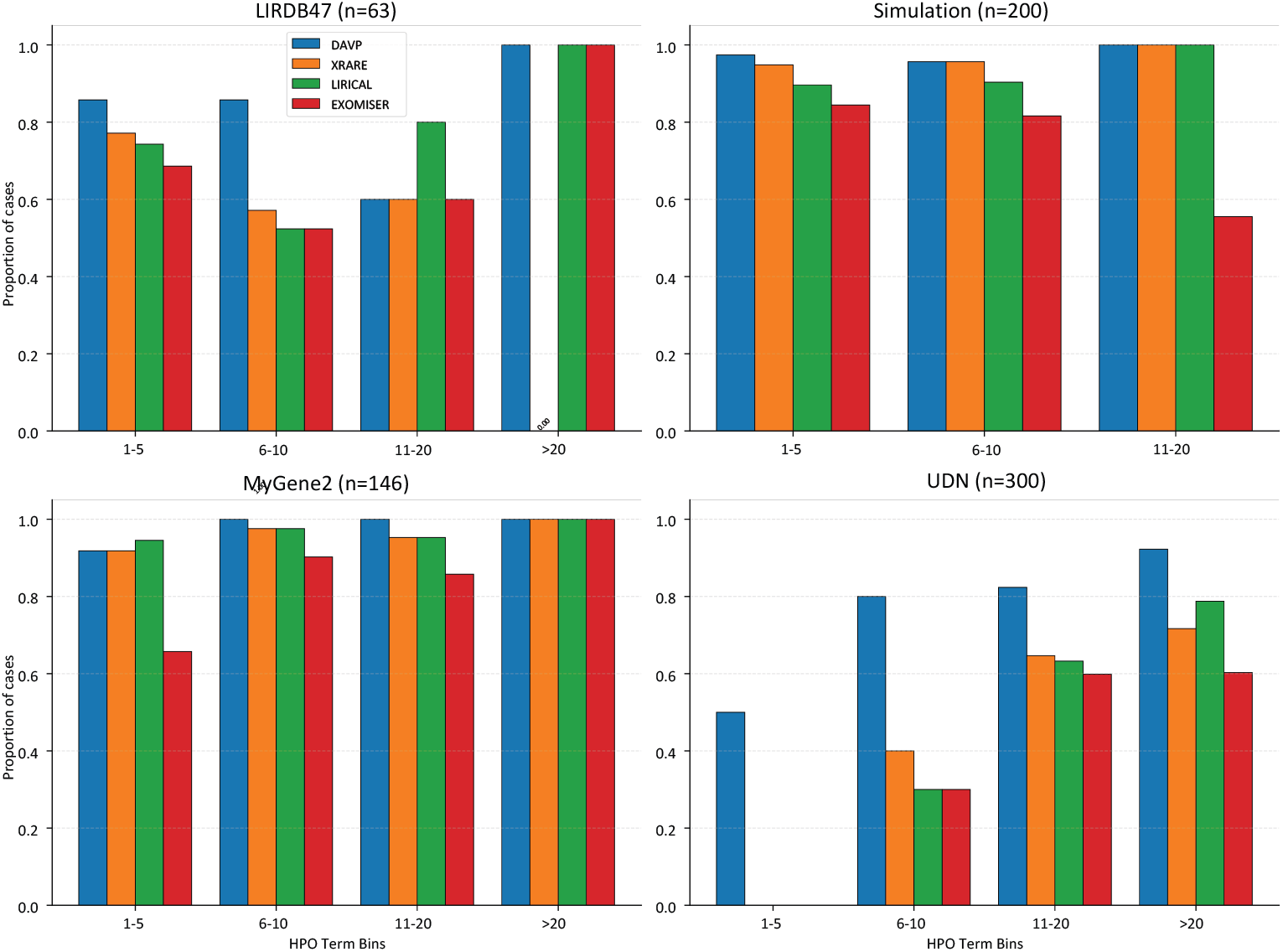
Top-10 gene recall stratified by the number of HPO terms per patient (bins: 1–5, 6–10, 11–20, *>*20). Bar colours encode method (DAVP, Xrare, LIRICAL, Exomiser). Performance generally improves with more phenotype information, though DAVP maintains robust performance even with limited HPO terms; numerical values for top-1, 3, 5, 10, and 20 gene recall are in Supplementary Table S23.

Supplementary Table S25 reports top-*k* recall within each class. DAVP maintains its lead across every class with at least ten cases, with the advantage particularly visible on the loss-of-function classes (stop gained, splice, start lost), where prior clinical knowledge about the mechanism is informative and can be surfaced through the gene cache and knowledge-graph reasoning. Intronic and non-coding classes remain the hardest for every method.

### ClinVar significance class

Supplementary Table S26 pools the simulated cohorts (Simulation, MyGene2, UDN; *n* = 644) and reports top-*k* recall for the Pathogenic/Likely Pathogenic (*n* = 484) and VUS (*n* = 160) subsets. DAVP’s lead is largest on the VUS subset, where Xrare drops to 1.88% top-1 and 14.37% top-3 while DAVP maintains 41.25% and 69.38%, respectively.

**Table S25:**
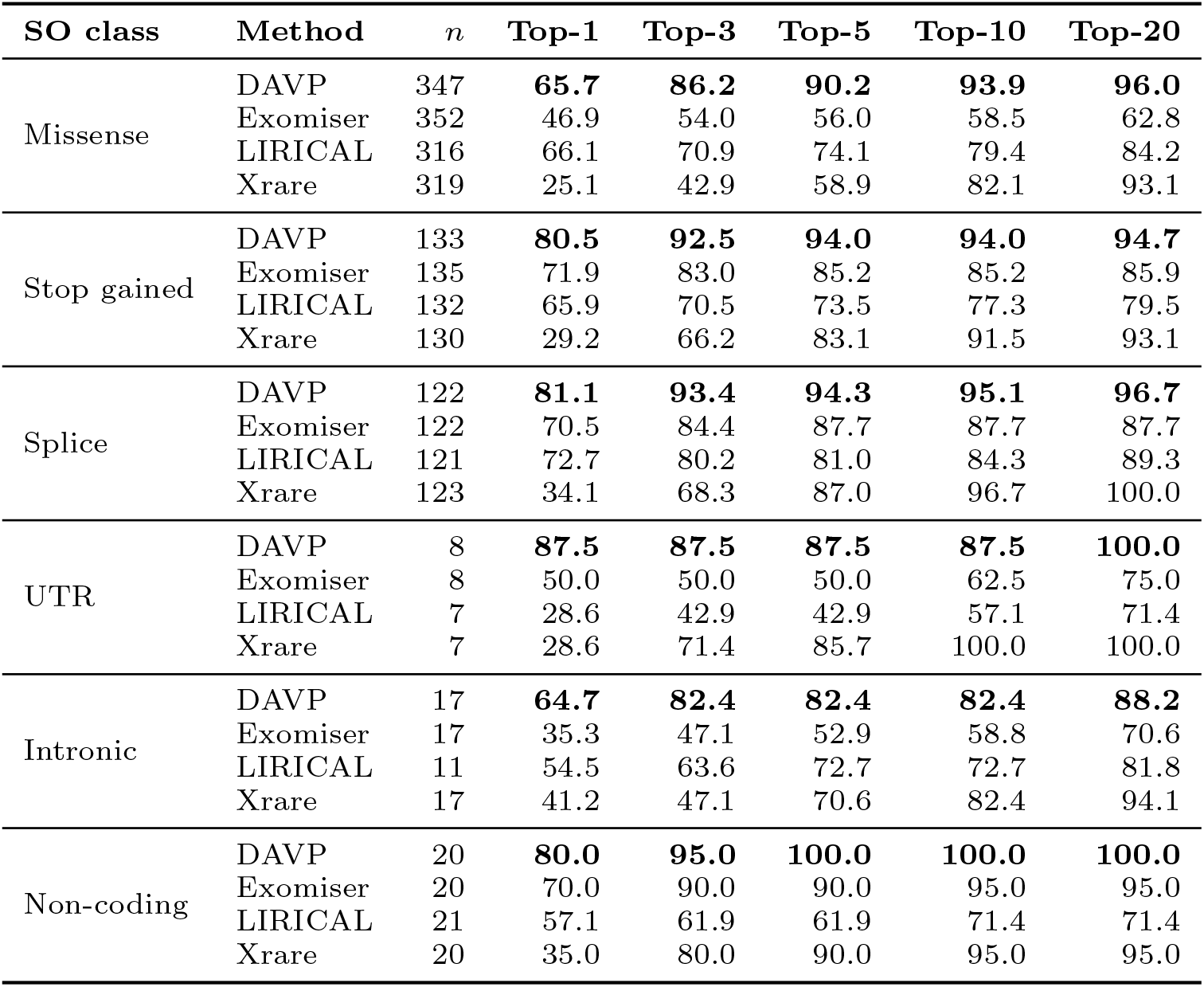
Top-*k* variant recall stratified by the ClinVar-reported Sequence Ontology molecular consequence of the answer variant. Classes with fewer than 10 cases are omitted (Frameshift (*n*=5), Start lost (*n*=4), Inframe indel (*n*=1), Synonymous (*n*=8)); the companion heatmap in Supplementary Figure S8 reports all classes.

**Fig. S8:**
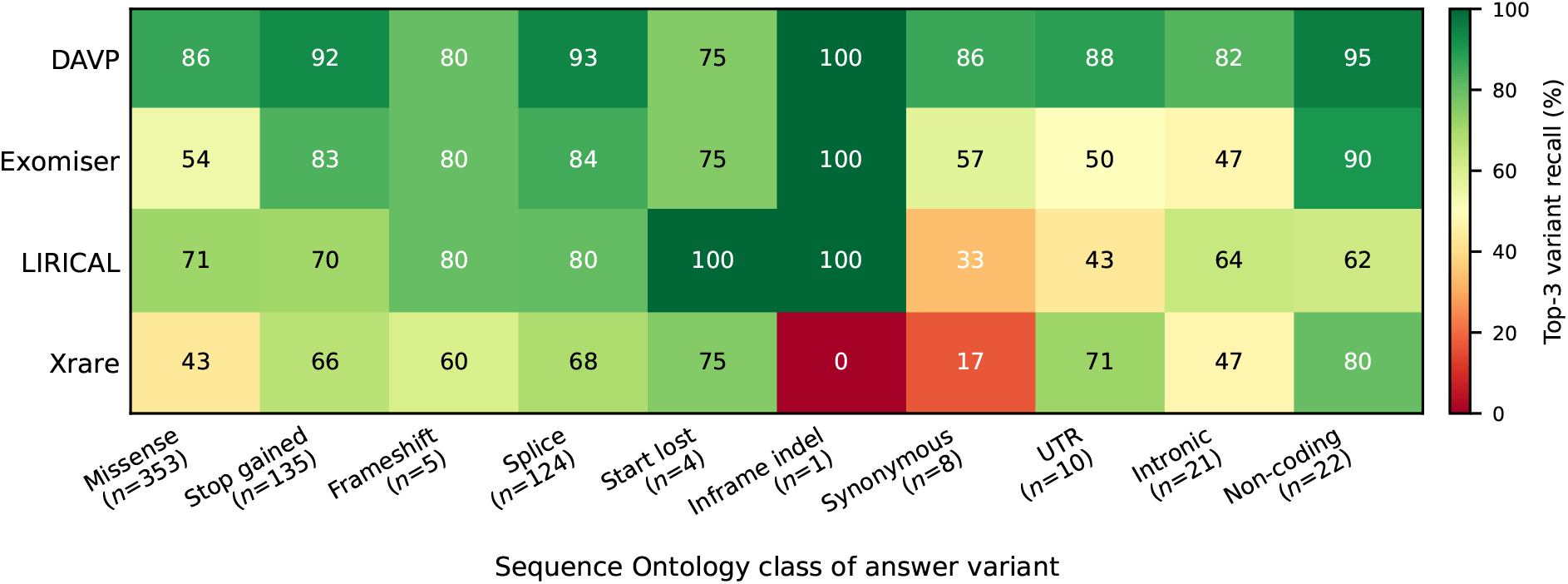
Top-3 variant recall (%) by method (rows) and answer-variant Sequence Ontology class (columns), colour-encoded from red (low) to green (high). Per-class case counts are shown beneath the column labels; classes with single-digit *n* carry wide sampling noise. The heatmap visualises the same axis as Supplementary Table S25 (which lists every top-*k* for *k* ∈ {1, 3, 5, 10, 20}): DAVP dominates every class with double-digit *n*, with the largest absolute lead on the loss-of-function classes (stop gained, splice, start lost) where mechanism-aware reasoning surfaced through the gene cache and knowledge graph contributes most.

**Table S26:**
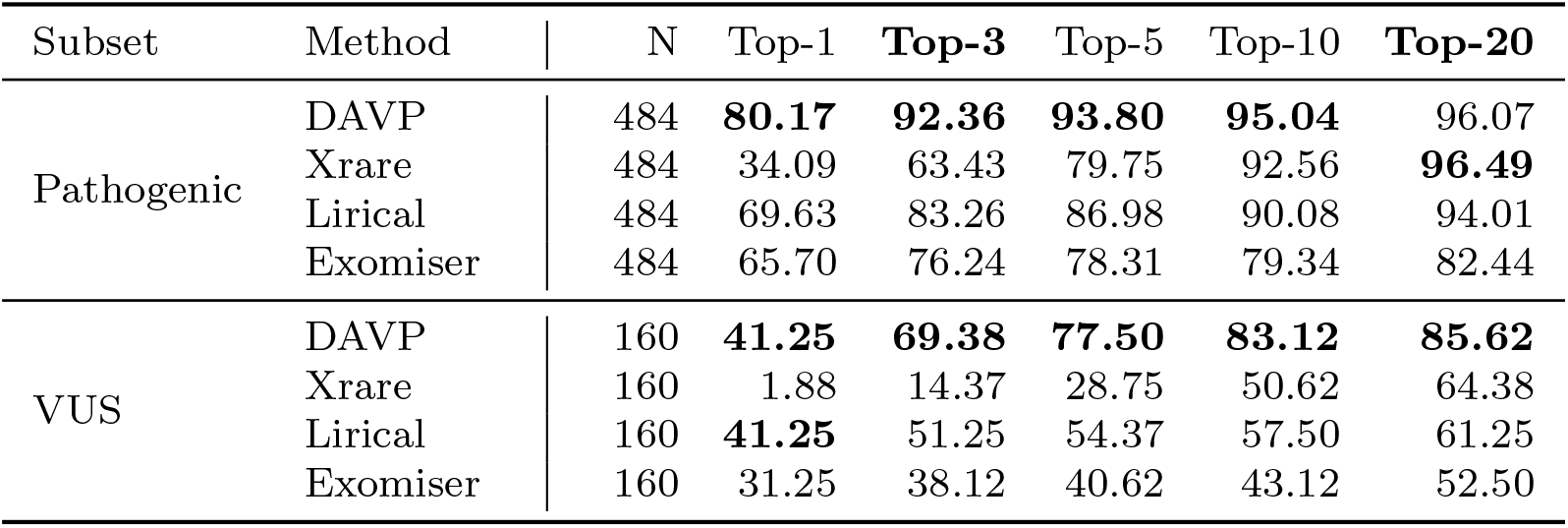
Recall Performance stratified by clinical subset: Pathogenic/Likely Pathogenic (Pathogenic) and Variants of Uncertain Significance (VUS).

**Table S27:** DAVP gene-level ranking performance stratified by ClinVar classification (Panel A) and review status (Panel B).

| Stratification | Group | N | Top-1 | **Top-3** | Top-5 | Top-10 | **Top-20** |
| --- | --- | --- | --- | --- | --- | --- | --- |
| ClinVar Class | Uncertain Significance | 121 | 38.0 | 67.8 | 76.0 | 81.0 | 83.5 |
|  | Conflicting | 39 | 51.3 | 74.4 | 82.1 | 89.7 | 92.3 |
|  | Pathogenic | 238 | 80.7 | 93.3 | 94.1 | 95.8 | 96.6 |
|  | Likely Pathogenic | 246 | 79.7 | 91.5 | 93.5 | 94.3 | 95.5 |
| Review Status | 0-1 | 147 | 68.7 | 86.4 | 88.4 | 93.2 | 93.9 |
|  | 2 | 306 | 66.3 | 82.7 | 86.9 | 88.6 | 90.8 |
|  | 3-5 | 193 | 77.7 | 92.2 | 94.3 | 95.9 | 96.4 |

**Table S28:**
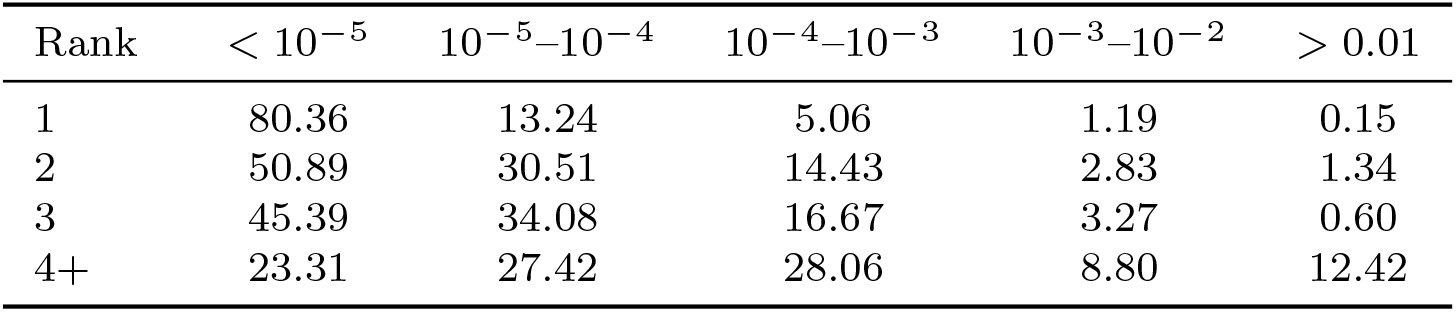
Allele frequency (AF) distribution of DAVP’s top-ranked candidate variants, by output rank position. Each row is a percentage breakdown across five AF bins (rows sum to 100%). Frequencies are gnomAD population AFs.

### ClinVar review status

Supplementary Table S27 additionally splits the same pool by ClinVar review-status star rating (Panel B) and provides a finer four-way significance class breakdown (Panel A) that distinguishes `uncertain` from `conflicting` interpretations and pathogenic from likely pathogenic.

### Allele frequency by output rank

Supplementary Table S28 reports the gnomAD allele-frequency distribution of DAVP’s top-ranked candidate variants, stratified by output rank position (1, 2, 3, 4+). Top-ranked variants are heavily concentrated at very low population frequencies (< 10^−5^ for 80.4% of rank-1 variants), with the AF distribution shifting toward more common alleles as rank position increases. This confirms that the top of DAVP’s output list is dominated by truly rare alleles, in line with the prior expectation for monogenic rare-disease causal variants.

**Table S29:** UDN sub-cohort top-*k* variant recall. The 300-case UDN cohort is split by causal-variant pathogenicity into a 150-case *Pathogenic* half (catalogued causal genes with prior ClinVar pathogenic annotation) and a 150-case *VUS* half (novel causal genes whose ground-truth variants are ClinVar VUS or conflicting interpretations). Per-method *n* values differ where a baseline could not score every case.

| Sub-cohort | Method | $n$ | Top-1 | Top-3 | Top-5 | Top-10 | Top-20 |
| --- | --- | --- | --- | --- | --- | --- | --- |
| UDN-Pathogenic | DAVP | 147 | 64.7 | **84.4** | **87.7** | **91.8** | 93.9 |
|  | Exomiser | 148 | **65.5** | 72.3 | 74.3 | 77.1 | 79.8 |
|  | LIRICAL | 144 | 65.3 | 70.1 | 72.2 | 75.7 | 79.9 |
|  | Xrare | 146 | 13.0 | 32.2 | 53.4 | 86.3 | **97.3** |
| UDN-VUS | DAVP | 142 | 41.6 | **73.3** | **81.7** | **88.1** | **90.8** |
|  | Exomiser | 146 | 31.5 | 37.0 | 39.1 | 40.4 | 45.9 |
|  | LIRICAL | 108 | **52.7** | 60.2 | 63.0 | 68.6 | 72.2 |
|  | Xrare | 120 | 2.5 | 17.5 | 32.6 | 61.7 | 80.0 |

## S11. UDN cohort deep-dive

This section provides a deeper characterisation of the 300-case UDN cohort introduced in the Methods, breaking out the Pathogenic and VUS sub-cohorts and analysing DAVP’s ranking behaviour against the per-case distractor variants spiked in by the benchmark.

Supplementary Table S29 reports per-method top-*k* recall on the two sub-cohorts separately. On the Pathogenic half DAVP, Exomiser, and LIRICAL converge near the top of the list at small *k*, with Exomiser narrowly leading at top-1 (65.5% vs. 64.7%) and DAVP taking over from top-3 onward. On the VUS half the gap widens: DAVP reaches 73.3% top-3 and 90.8% top-20, against 60.2% and 72.2% for LIRICAL and 17.5% and 80.0% for Xrare. DAVP’s advantage is largest on the VUS sub-cohort, where database lookups provide the least discriminative signal and the gene-cache plus knowledge-graph evidence is the dominant ranking input.

### Distractor burden

Each UDN case carries, alongside its causal variant, a small set of additional candidate variants that the benchmark spikes in as plausible-but-wrong distractors; the spiked set is recorded in the truth file’s `candidate_allele_ids_`field and contains 3–19 variants per case (mean 11.0 ± 3.1). Because the distractors are by construction biologically plausible (rare, coding, often in OMIM disease genes), they are the operative source of difficulty for the prioritiser, and stratifying recall by distractor count rather than by raw exome size is the meaningful axis. Supplementary Table S30 reports per-method top-*k* recall jointly stratified by causal-variant pathogenicity (Pathogenic vs. VUS) and by per-case distractor-count bin (3–7, 8–12, 13+). The cohort-level signal carries through every bin: on the VUS half DAVP clears 70% top-3 in every bin, while Exomiser stays below 43%, LIRICAL between 46 and 71%, and Xrare below 21%. Supplementary Figure S10 visualises the same data at top-5 and shows that DAVP’s recall is approximately constant across distractor-count bins (76–86% on VUS, 82–100% on Pathogenic) whereas the three baselines decline monotonically with distractor count on both halves.

### Robustness regression

To put a single number on this distractor-robustness story we regress log_10_(rank) on the per-case distractor count for each method. On the harder VUS half DAVP’s slope is +0.003 (*R*^2^ = 0.0, i.e. rank is statistically independent of distractor count over the 3–19 range), against +0.031 for Exomiser, +0.020 for LIRICAL and +0.018 for Xrare; equivalently, each additional spiked distractor multiplies DAVP’s expected rank by 1.006× but multiplies Exomiser’s by 1.075×. On the Pathogenic half all four methods carry small positive slopes (DAVP +0.024, Exomiser +0.017, LIRICAL +0.052, Xrare +0.042), consistent with the cohort being uniformly easier and dominated by gene-level signal rather than variant-level distractor competition. The per-method robustness gap is therefore concentrated where it matters: on novel-gene VUS cases, gene-cache and knowledge-graph evidence carry the ranking decision while pathogenicity-score baselines degrade with the size of the distractor set.

**Fig. S9:**
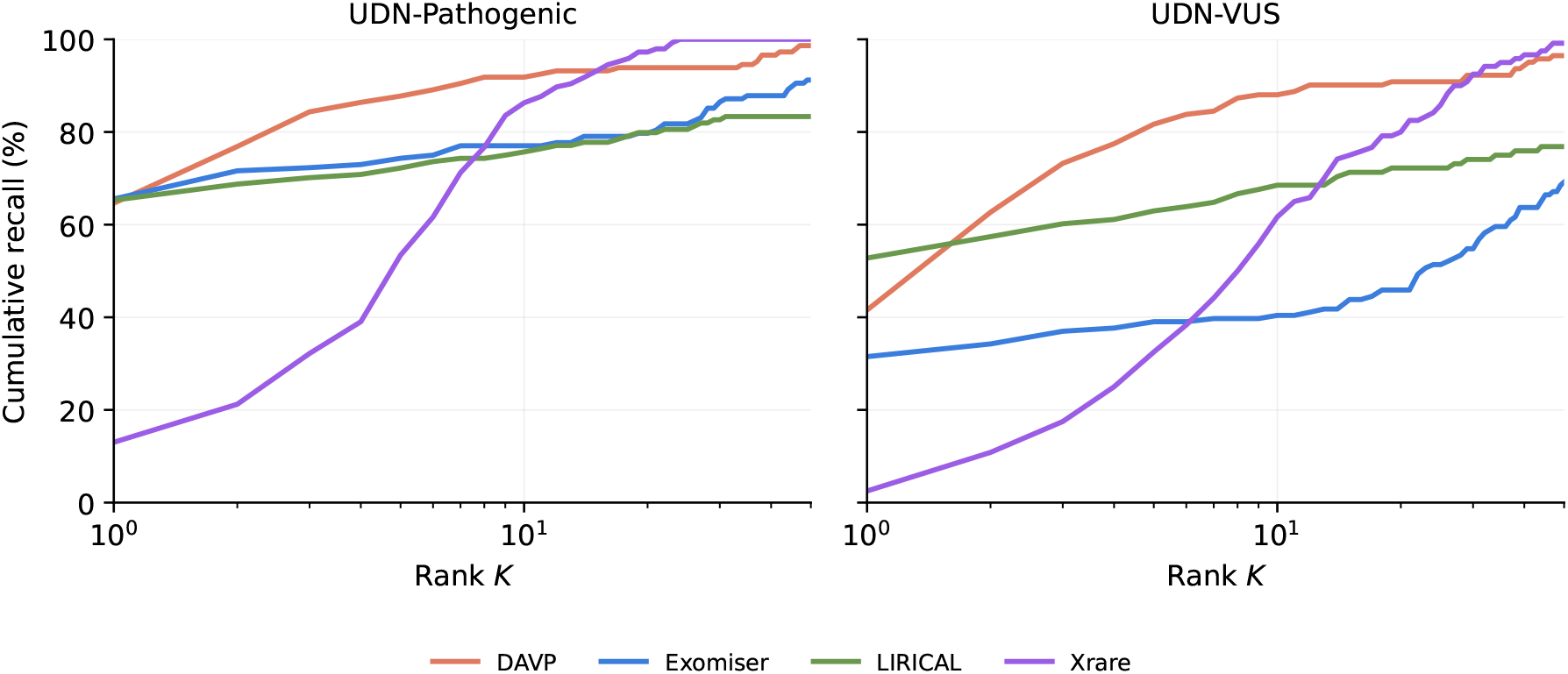
Per-method cumulative distribution of the answer-variant rank on the two halves of the UDN cohort. Each panel plots the empirical recall curve Pr[rank ≤ *K*] for DAVP, Exomiser, LIRICAL and Xrare; the rank axis is log-scaled to resolve the small-*K* behaviour where clinical decision support operates. On the Pathogenic half the four methods converge near *K*=1 and DAVP pulls ahead from rank 3 onward. On the VUS half DAVP retains a 25–40 percentage-point lead over the strongest baseline through *K*=20, while Exomiser and LIRICAL flatten well below their Pathogenic curves and Xrare requires *K*≥10 to recover.

**Table S30:** Per-method top-*k* variant recall on the UDN cohort, stratified by causal-variant pathogenicity (Pathogenic vs. VUS) and by per-case spiked-distractor count. Each UDN case carries the causal variant plus a benchmark-supplied set of plausible-but-wrong distractor variants (3–19 per case, mean 11.0 ± 3.1); the bin label indicates how many distractors were spiked into the cases pooled in that row. Sample sizes (*n*_cases_) are at the cohort-bin level; per-method totals can be slightly smaller where a baseline could not score every case (counted in the rightmost column). DAVP’s lead is largest on the VUS half, where database lookups carry the least signal, and is approximately constant across distractor-count bins. Bold marks the best method in each (cohort, bin, top-*k*) cell.

| Cohort | Bin | $n_{\text{cases}}$ | Method | Top-1 | Top-3 | Top-5 | Top-10 | Top-20 | $n_{\text{m}}$ |
| --- | --- | --- | --- | --- | --- | --- | --- | --- | --- |
| Pathogenic | 3-7 | 16 | DAVP | 80.0 | **100.0** | **100.0** | **100.0** | **100.0** | 15 |
|  |  |  | Exomiser | 80.0 | 86.7 | 86.7 | 86.7 | 86.7 | 15 |
|  |  |  | LIRICAL | **93.8** | 93.8 | 93.8 | 93.8 | 93.8 | 16 |
|  |  |  | Xrare | 33.3 | 66.7 | 73.3 | 93.3 | **100.0** | 15 |
|  | 8-12 | 78 | DAVP | **69.7** | **88.2** | **89.5** | 93.4 | 93.4 | 76 |
|  |  |  | Exomiser | 66.2 | 74.0 | 75.3 | 77.9 | 80.5 | 77 |
|  |  |  | LIRICAL | 68.0 | 74.7 | 77.3 | 80.0 | 81.3 | 75 |
|  |  |  | Xrare | 13.2 | 31.6 | 55.3 | **94.7** | **98.7** | 76 |
|  | 13+ | 56 | DAVP | 53.6 | **75.0** | **82.1** | **87.5** | 92.9 | 56 |
|  |  |  | Exomiser | **60.7** | 66.1 | 69.6 | 73.2 | 76.8 | 56 |
|  |  |  | LIRICAL | 52.8 | 56.6 | 58.5 | 64.2 | 73.6 | 53 |
|  |  |  | Xrare | 7.3 | 23.6 | 45.5 | 72.7 | **94.5** | 55 |
| VUS | 3-7 | 27 | DAVP | 28.0 | **60.0** | **76.0** | **80.0** | **84.0** | 25 |
|  |  |  | Exomiser | 38.5 | 42.3 | 46.2 | 46.2 | 50.0 | 26 |
|  |  |  | LIRICAL | **40.9** | 50.0 | 54.5 | 54.5 | 59.1 | 22 |
|  |  |  | Xrare | 4.8 | 19.0 | 42.9 | 66.7 | 81.0 | 21 |
|  | 8-12 | 83 | DAVP | 47.5 | **78.8** | **86.2** | **92.5** | **93.8** | 80 |
|  |  |  | Exomiser | 32.1 | 39.5 | 42.0 | 43.2 | 48.1 | 81 |
|  |  |  | LIRICAL | **65.5** | 70.7 | 72.4 | 79.3 | 81.0 | 58 |
|  |  |  | Xrare | 2.9 | 20.6 | 35.3 | 66.2 | 83.8 | 68 |
|  | 13+ | 40 | DAVP | **37.8** | **70.3** | **75.7** | **83.8** | **89.2** | 37 |
|  |  |  | Exomiser | 25.6 | 28.2 | 28.2 | 30.8 | 38.5 | 39 |
|  |  |  | LIRICAL | 35.7 | 46.4 | 50.0 | 57.1 | 64.3 | 28 |
|  |  |  | Xrare | 0.0 | 9.7 | 19.4 | 48.4 | 71.0 | 31 |

**Fig. S10:**
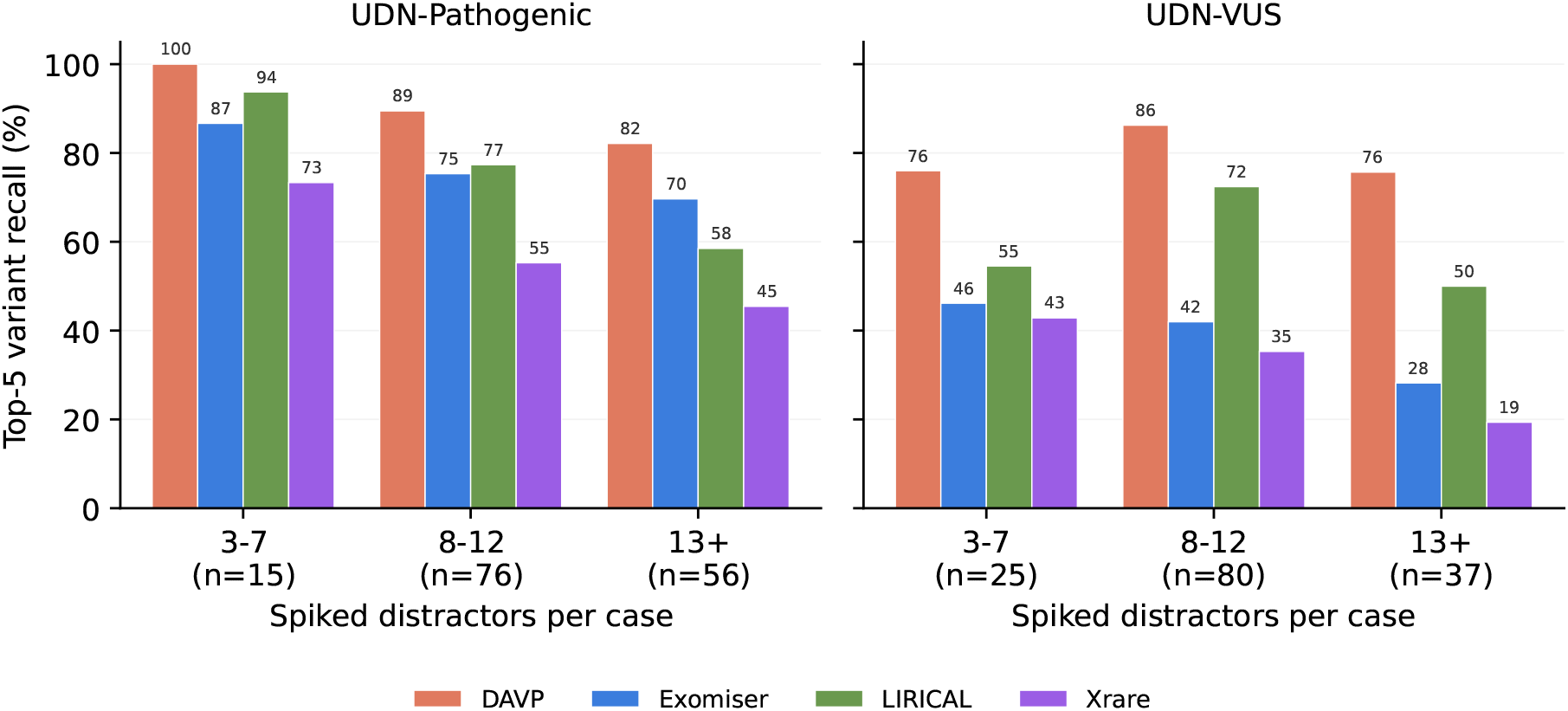
Per-method top-5 variant recall on the UDN cohort, stratified by causal-variant pathogenicity (UDN-Pathogenic, UDN-VUS) and by spiked-distractor count per case (3–7, 8–12, 13+). Tick labels report the per-bin number of cases. DAVP’s lead is largest on the VUS half and is approximately constant across distractor-count bins, consistent with the per-method log-rank slope reported in the prose. Numerical values for additional top-*k* thresholds are in Supplementary Table S30.

